# Depletion of effector regulatory T cells drives major response to induction dual immune checkpoint blockade

**DOI:** 10.1101/2024.01.04.23300616

**Authors:** Xianli Jiang, Nils-Petter Rudqvist, Bo Jiang, Shengbin Ye, Shan He, Qingnan Liang, Jinzhuang Dou, Michelle Williams, Joe Dan Dunn, Jason M. Johnson, Keiko Akagi, Weihong Xiao, Shaoheng Liang, Satvik Elayavalli, Baohua Sun, Edwin Roger Parra Cuentas, Renata Ferrarotto, Adam Garden, Clifton Fuller, Jay Reddy, Neil Gross, Miriam Lango, Cheuk Hong Leung, Suyu Liu, Diane Liu, J Jack Lee, Michael A. Curran, Jack Phan, Ken Chen, Maura L. Gillison

## Abstract

In a phase 2 trial, local-regionally advanced HPV-positive oropharyngeal carcinoma (OPC) patients received ipilimumab (anti-CTLA-4) and nivolumab (anti-PD-1) as induction immunotherapy and concurrently with radiotherapy (NCT03799445). Co-primary endpoints achieved included 6-month complete metabolic response rate (94%) and 2-year progression-free survival (84%). Induction yielded a 46% major histological response rate. Single-cell profiling revealed responders had higher baseline intratumoral tissue-resident memory (TRM) CD8^+^ T cells and NK cells expressing Fc Gamma Receptor IIIa (*FCGR3A*). Decreases in effector regulatory T (eTreg) cells, which highly expressed *CTLA4*, occurred only in responders, suggesting ipilimumab-dependent depletion by *FCGR3A^+^* NK cells. eTreg depletion correlated with increased Interferon Gamma (*IFNG*)^+^ effector CD8^+^ T cells. CD8^+^ T-cell clonotypes transitioned from TRM to effector memory and *IFNG^+^* effector cells in responders, whereas clonotypes transitioned to exhausted TRM and proliferating cells in nonresponders. We conclude that eTreg depletion is critical for major response to induction dual immune checkpoint blockade.

## INTRODUCTION

Human papillomavirus-positive oropharyngeal squamous cell carcinoma (HPV-positive OPC) has an improved prognosis relative to HPV-negative head and neck SCC (HNC)^1^, due in part to enhanced immune surveillance by greater numbers of tumor-infiltrating lymphocytes^2–6^. Most HPV16-positive tumors contain E6- and E7-specific CD8^+^ and CD4^+^ T cells with a type 1 cytokine profile (e.g., IFNγ, TNFα, and IL-2)^7^, and circulating T cells that recognize E1, E2, E4, E5 and L1 epitopes have been detected^8^. Single-cell RNA sequencing (scRNA-seq) of intratumoral HPV16-specific PD-1^+^CD8^+^ T cells revealed stem-like phenotypes with capacity to proliferate and differentiate into effector T cells^9^. However, HPV-positive OPC also have higher regulatory T cell (Treg) to CD8^+^ T cell ratios than several other solid tumors^4^. Immunofluorescence studies have confirmed a high density of CD8^+^ T cells near Tregs in HPV-positive tumors^10,11^. Thus, immunotherapies capable of depleting Tregs and restoring functionality of a pre-existing HPV-reactive T cell population would have tremendous therapeutic potential in newly diagnosed HPV-positive OPC.

Immune checkpoint blockade (ICB) targeting PD-1 improves survival of recurrent or metastatic (R/M) HNSCC relative to platinum-based chemotherapy in tumors that express its ligand PD-L1^12^ and in platinum-refractory disease^13^. However, the addition of CTLA-4 to PD-1 ICB did not improve survival relative to single-agent PD-1 ICB^14,15^. PD-(L)1 ICB concurrent with and adjuvant to chemoradiotherapy also did not improve cancer control in patients with newly diagnosed HNC^16,17^. Negative effects of high-dose radiotherapy on both T-cell priming in tumor-draining lymph nodes and on systemic immunity are hypothesized to contribute to these negative results. In contrast, neoadjuvant (i.e., systemic therapy prior to resection) ICB of intact tumors may increase antigen presentation, T-cell priming, T-cell receptor (TCR) clonality and systemic immunity vs adjuvant ICB, in part because the patient’s immune system has not been suppressed by prior therapy^18,19^. In support of this, the addition of neoadjuvant PD-1 ICB improved event-free survival vs adjuvant ICB alone in advanced, resectable melanoma^20^. Given pathological response to neoadjuvant therapy is a surrogate for long-term disease control in several solid tumor types, including breast^21^, lung^22^, melanoma^23^, and HPV-negative HNC^24^, neoadjuvant trials can also provide an early signal of clinical benefit. Phase 2 trials of neoadjuvant PD-(L)1 +/− CTLA-4 ICB in resectable HPV-negative HNC have observed major pathological response (MPR; <10% viable tumor) rates of 7–35%^24–27^. Only two trials included small subsets of HPV-positive OPSCC^28,29^; therefore, response to neoadjuvant ICB in this patient population is largely unexplored.

Given pre-existing HPV-specific T-cell responses are abundant in the tumor immune microenvironment (TIME) of HPV-positive OPC, we performed a phase 2 clinical trial in which newly diagnosed patients received CTLA-4 and PD-1 ICB as induction (i.e., systemic therapy prior to radiotherapy) followed by concurrent ICB with intensity modulated radiotherapy (hereafter, XRT). We profiled tumors by scRNA-seq and single-cell TCR sequencing (scTCR-seq) at baseline and on induction to investigate mechanisms of response and resistance.

## RESULTS

From July 2019 to November 2021, 37 patients with newly diagnosed PD-L1 combined positive score (CPS) ≥1, AJCC 8^th^ edition Stage I-II (T1-T3N1-N2M0, T3N0M0), HPV-positive OPC (regardless of smoking history) were treated with a six-week cycle of ipilimumab (anti-CTLA-4) and <u>n</u>ivolumab (anti-PD-1) ICB followed by a second cycle concurrent with dose and <u>v</u>olume response-<u>a</u>dapted <u>X</u>RT (INVAX trial; **Fig. 1a**). To minimize effects on tumor draining lymph nodes, elective nodal XRT was restricted to one adjacent echelon. Co-primary endpoints included estimates of 6-month complete metabolic response (CMR) rate by fluorodeoxyglucose positron emission tomography (FDG-PET) and 2-year progression-free survival (PFS).

**Fig. 1.**
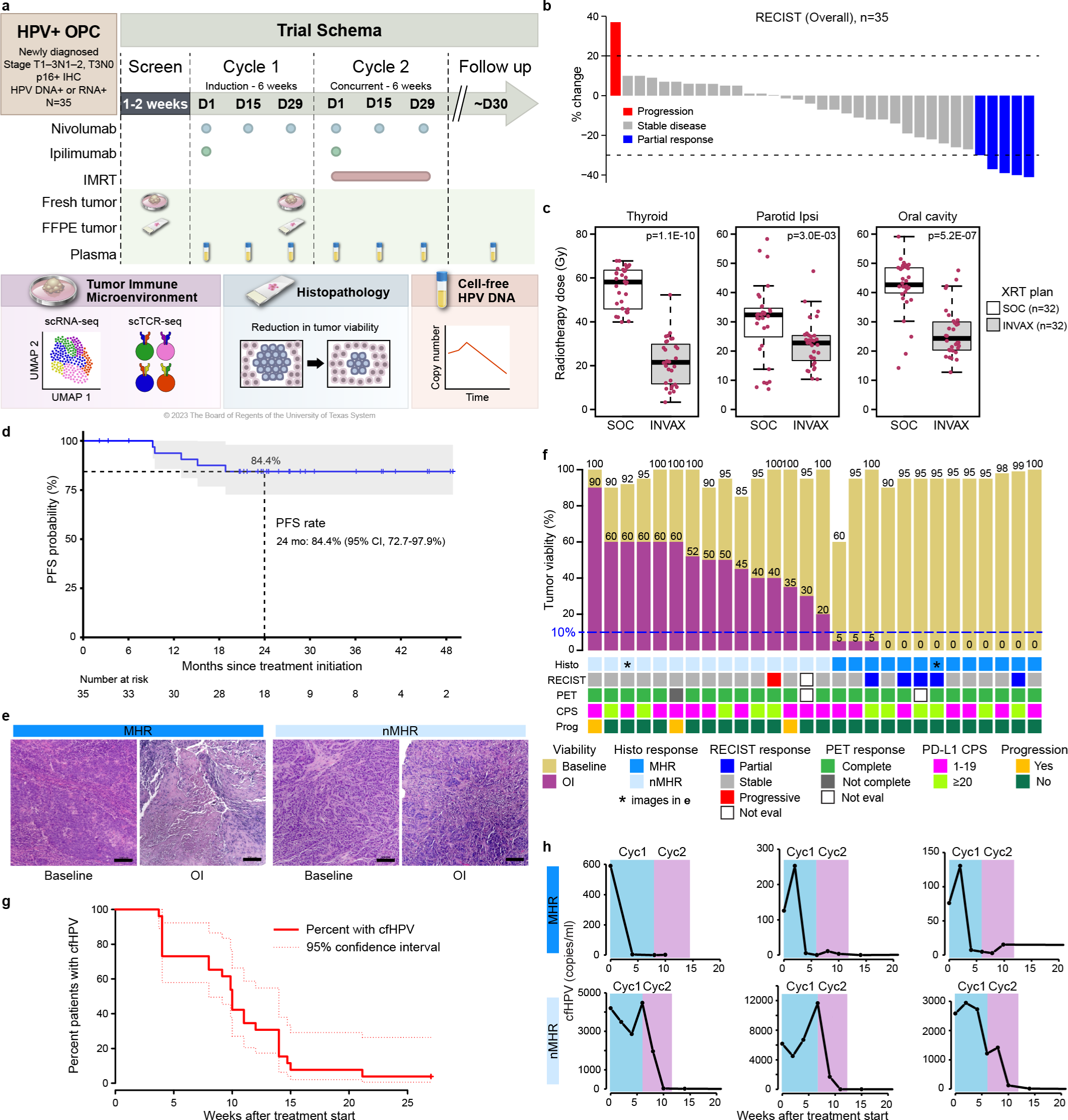
Induction dual immune checkpoint blockade results in major histological responses in patients with HPV-positive oropharyngeal cancer. **a,** Schema of phase 2 clinical trial of a six-week cycle of anti-CTLA-4 (ipi) and anti-PD-1 (nivo) given as induction immune checkpoint blockade (ICB) followed by a six-week cycle of concurrent ICB and intensity modulated radiotherapy (IMRT) in patients with HPV-positive oropharyngeal squamous cell carcinoma (OPC). Biospecimens collected and analyzed as indicated. **b,** Waterfall plot of radiological response to induction ICB, determined per RECIST 1.1 by a neuroradiologist, masked to clinical data, by independent review of baseline and post-induction computed tomography scans. y-axis, percent change in measurable disease volume. **c,** Radiotherapy (XRT) dose delivered per standard-of-care (SOC) vs per trial protocol (INVAX) plans to thyroid (mean dose, 55.7 vs 21.9 Gy), ipsilateral parotid (mean dose, 30.1 vs 22.3 Gy), and oral cavity (mean dose, 40.2 vs 25.9 Gy). Dots, individual patient values; midline, median; box limits, interquartile range; whiskers, 1.5x interquartile range. Wilcoxon rank sum test used for comparison. *P*-values were adjusted using the false discovery rate method. **d,** Kaplan-Meier curve of progression-free survival (PFS; n=35). Shaded area, 95% confidence interval. **e,** Representative images of FFPE slides stained with hematoxylin and eosin from a major histological responder (MHR; <10% viable tumor) and a non-MHR (nMHR) at baseline and on induction (OI) ICB. Percent viable tumor was calculated as the viable tumor area/total tumor bed area (in mm^2^). The total tumor bed was the sum of the regression bed (characterized by lymphoid infiltrate, proliferative fibrosis, and neovascularization), residual viable tumor, and necrosis. **f,** Baseline and OI tumor viability (y-axis) determined for 28 patients (bars) with paired baseline-OI FFPE slides. Grid indicates histological (histo), overall RECIST (RECIST), and metabolic/fluorodeoxyglucose positron emission tomography (PET) response for the 28 patients, in addition to PD-L1 combined positive score (CPS) and whether disease progression (Prog) occurred during follow up. Metabolic/PET response was determined 6 months after completion of Cycle 2 per Hopkin’s criteria. *, panel **e** includes images of FFPE slides from patient; Not eval, not evaluable. **g,** Kaplan-Meier curve of time to plasma cell-free HPV DNA (cfHPV) clearance measured by digital-droplet PCR for 26 patients with quantifiable cfHPV. **h,** Representative graphs of cfHPV copy number per ml plasma (y-axis) over time (x-axis) for 3 MHRs (top) and 3 nMHRs (bottom). HPV, human papillomavirus; FFPE, formalin fixed and paraffin-embedded tumor sample; UMAP, uniform manifold approximation and projection; scRNA-seq, single-cell RNA sequencing; scTCR-seq, single-cell T-cell receptor sequencing; Cyc1, first treatment cycle; Cyc2, second treatment cycle.

Two patients were excluded from clinical trial endpoint analysis due to ineligibility or a protocol deviation (**Extended Data Fig. 1**). Patient characteristics are shown in **Supplementary Tables 1 and 2**. PD-L1 CPS score was 1–19 in 63% and ≥20 in 37%. HPV types in tumors by digital-droplet (dd)PCR included HPV16 (83%), HPV33 (9%) and other (1 each of HPV18, HPV35, and HPV45). Five cases were intermediate risk by RTOG0129 criteria^1^. Fifteen (43%) were never, 9 (26%) current, and 11 (31%) former tobacco users. For 13 cigarette smokers, median pack-years was 10 (interquartile range [IQR] 5–30).

### Treatment-related adverse events

One dose-limiting toxicity of grade 3 mucositis occurred among 8 patients during the safety lead, so the study proceeded to phase 2. Maximum treatment-related adverse events (TRAEs) were grade 1-2 in 34% and grade≥3 in 66% (e.g., odynophagia and mucositis, **Supplementary Table 3**). Any-grade TRAE attributable to ICB (**Supplementary Table 4**) included hypothyroidism (n=16), adrenal insufficiency (2), colitis (2), hyperglycemia (1), pancreatitis (1), and myositis (1). Thirteen (37%) patients experienced a grade≥3 TRAE attributable to ICB (**Supplementary Table 5**). The only grade 4 TRAE was asymptomatic lipase increase. ICB was discontinued for five patients due to toxicity during induction (grade 2 myositis, grade 3 pancreatitis or hyperglycemia), RECIST progression after cycle 1, or physician concern for progression. These patients received XRT alone per INVAX plan (n=1) or standard of care (SOC) XRT (n=1) concurrent with weekly platinum chemotherapy (n=3).

### RECIST response and dose and volume-adapted radiotherapy

Modified RECIST 1.1 overall response after induction ICB (iICB) was determined per independent review. Five (14%) patients had a partial response, 29 (83%) stable disease, and one, disease progression (**Fig. 1b**). INVAX XRT plans were evaluable for 31 patients. Four patients with RECIST progression/physician concern (n=2) or toxicity (n=2) during iICB received SOC XRT plans. Eleven patients received ipsilateral neck XRT for a well-lateralized tonsil primary. Dose to primary clinical target volume (CTV) was 66 Gy in 18, 60 Gy in 12, and 50 Gy in 1 patient. Dose to nodal CTV was 66 Gy in 16, 60 Gy in 13, and 50 Gy in 2 patients. Compared to plans for each patient per SOC dose/fractionation and planning parameters, INVAX plans showed significantly lower doses to the thyroid, ipsilateral parotid, and oral cavity (**Fig.1c**), in addition to the contralateral submandibular gland and parotid and the larynx (**Supplementary Fig. 1**).

### Disease control endpoints

All 35 evaluable patients had an FDG-PET score of 5 per Hopkins Criteria at enrollment. At 3 and 6 months after XRT, 33 patients were evaluable for FDG-PET response (1 withdrawn consent, 1 not done). CMR rate by 6 months was 94.3% (95% CI: 80-99%). Two patients with a positive FDG-PET at 6 months had biopsy-confirmed disease (1 local, 1 regional) and were treated with stereotactic body XRT alone or after neck dissection.

After a median follow-up of 24.5 months (data cut-off October 13, 2023), five progression events had occurred. Patterns of failure included local (2), regional (2), or distant (1). Local failures were in the subclinical field (i.e., 36 Gy) or outside, and regional failures were in-field (60 Gy) or subclinical field. The patient with distant metastasis received induction and SOC cisplatin-XRT. 2-year PFS and overall survival (OS) were 84.4% (95% CI: 72.7-97.9%; **Fig.1d**) and 100%, respectively. Two patients were alive with cancer at the data cutoff. Cancer progression was more common in patients with a history of tobacco use (5/20 vs 0/14, Fisher’s exact p=0.06).

### Histopathological response

Paired baseline and on induction tumor biopsies with available hematoxylin and eosin-stained slides were collected from 28 patients (missing data due to COVID shutdown). Of these, 96% had ≥20% reduction in tumor viability, i.e., pathological treatment effect^24^ (**Fig. 1e**). Median tumor viability declined from baseline (95% vs 25%, p=4.5 e-13). Thirteen (46%) patients were major histological responders (MHRs; <10% viable tumor in biopsy) to iICB^30^. MHR status was associated with RECIST response (Fisher’s exact p=0.01).

### Dynamics of cfHPV during therapy

Because tumor biopsies may not accurately reflect overall response, we evaluated quantitative changes in plasma cell-free HPV DNA (cfHPV) as a measure of minimal residual disease (MRD). Of 35 patients, 91% were positive for cfHPV (≥5 copies/ml plasma) at baseline and 26 were quantifiable (≥16 copies/ml plasma; median 255, IQR 107–589), with a median time to clearance of 10 weeks, corresponding to Cycle 2 Day 29 (**Fig. 1g**). In the 20 patients with quantifiable cfHPV evaluable for histological response, cfHPV clearance occurred on iICB in 71.4% of MHRs vs 0% of non-MHRs (Fisher’s exact p=0.001; representative plots in **Fig. 1h**). Thus, most MHRs were MRD negative prior to the start of XRT.

### TIME composition and response to iICB

We conducted scRNA-seq and scTCR-seq on 64 fresh tumor biopsies, including 29 baseline and on-induction pairs. After quality filtering (**Supplementary Fig. 2**), we profiled 367,451 cells, which distributed by unsupervised clustering into 11 cell types (**Fig. 2a, Supplementary Fig. 3, and Supplementary Table 6**). Here we focus on the lymphocyte population, which included 78,622 CD4^+^ T cells (of which 20,006 were Tregs), 76,319 CD8^+^ T cells, and 8,000 natural killer (NK) cells.

**Fig. 2.**
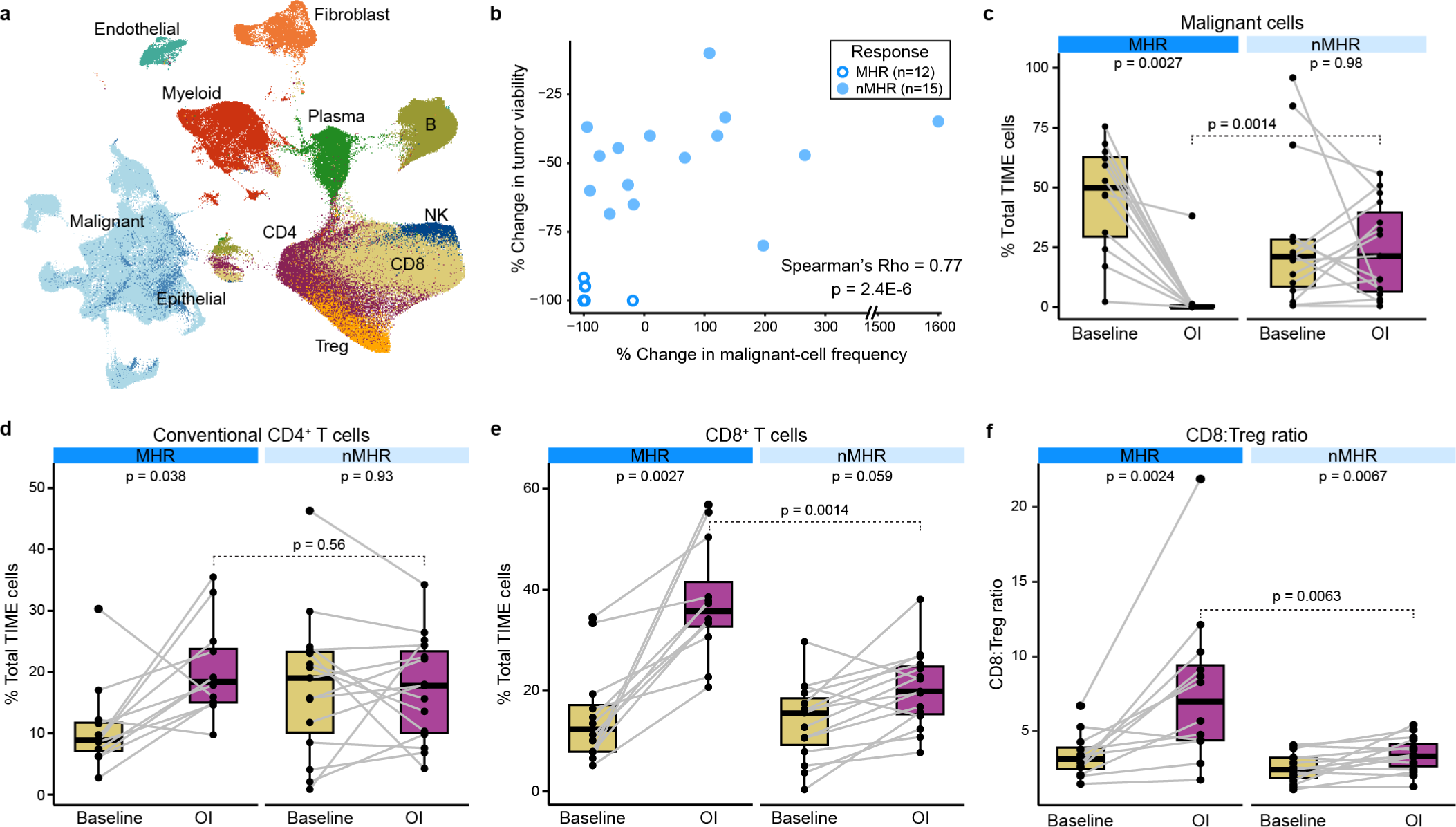
Increased total CD8^+^ T cells and CD8:Treg ratio in the TIME associated with response to induction dual immune checkpoint blockade. scRNA-seq was performed on fresh tumor biopsies collected at baseline and on induction (OI) CTLA-4 and PD-1 immune checkpoint blockade from 35 patients (64 samples in total, including 27 matched baseline-OI pairs with histological response information). Patients were classified as major histological responders (MHRs; n=12) or non-MHRs (nMHRs; n=15). **a,** UMAP of 367,451 tumor immune microenvironment (TIME) cells color-coded by cell type. B, B cell; Plasma, plasma cell; CD4, conventional CD4^+^ T cell; Treg, regulatory CD4^+^ T cell; CD8, CD8^+^ T cell; NK, natural killer cell. **b,** Scatterplot of percent change in tumor viability OI compared to baseline per histological review (y-axis) vs percent change in malignant cell frequency OI compared to baseline per single-cell data analysis (x-axis). **c-f,** Proportion of malignant cells (**c**), conventional CD4^+^ T cells (**d**), or CD8^+^ T cells (**e**) or the CD8:Treg ratio (**f**) at baseline and OI in MHRs and nMHRs. Dots, individual patient values; midline, median; box limits, interquartile range; whiskers, 1.5x interquartile range; grey lines, matched baseline-OI sample pairs. Wilcoxon signed-rank test and Wilcoxon rank-sum test used for comparisons within (paired) and between response groups, respectively. c-e, *P*-values were false discovery rate adjusted.

To gain insight into mechanisms of iICB response, we focused on differences and changes in the TIME in MHRs vs nMHRs (**Supplementary Table 2**). Malignant cells were the most abundant cell type at baseline (**Extended Data Fig. 2**). A strong correlation was observed between reduction from baseline in malignant cell proportion in scRNA-seq data and in tumor viability in histopathology (**Fig. 2b**). Notably, malignant cell proportion reduced significantly on iICB in MHRs but not nMHRs (**Fig. 2c**). High rates of cfHPV clearance and malignant cell proportion in scRNA-seq data support the stratification of our analysis by MHR status. MHRs had a significant increase in the proportions of CD4^+^ and CD8^+^ T cells in response to iICB whereas nMHRs did not (**Fig. 2d-e**). MHRs also had significantly higher CD8:Treg ratios on iICB than nMHRs (**Fig. 2f**). Given this ratio is associated with both treatment response and survival in a variety of solid tumors^31,32^, we sought to identify its underlying contributing factors in the TIME.

### Induction ICB decreases effector Treg frequency in MHRs

We defined 14 CD4^+^ T cell clusters (**Extended Data Fig. 3a,b, Supplementary Fig. 4a-d, Supplementary Table 7,8**) and observed no differences in their baseline frequency in MHRs vs nMHRs (**Extended Data Fig. 3c**). On iICB, however, MHRs had a significant reduction in frequency of the highly immunosuppressive effector Treg (eTreg) cluster (**Fig. 3a, Extended Data Fig. 3d,e**) characterized by high expression of *FOXP3, CTLA4, IL2RA/CD25, TNFRSF9/41BB*, and *CCR8* (**Fig. 3b, Supplementary Fig. 4e,f**). Mean frequency declined by 60.5% in MHRs vs a 4.8% increase in nMHRs. The eTreg cluster is analogous phenotypically to that reported in lung, liver, and melanoma^33^.

**Fig. 3.**
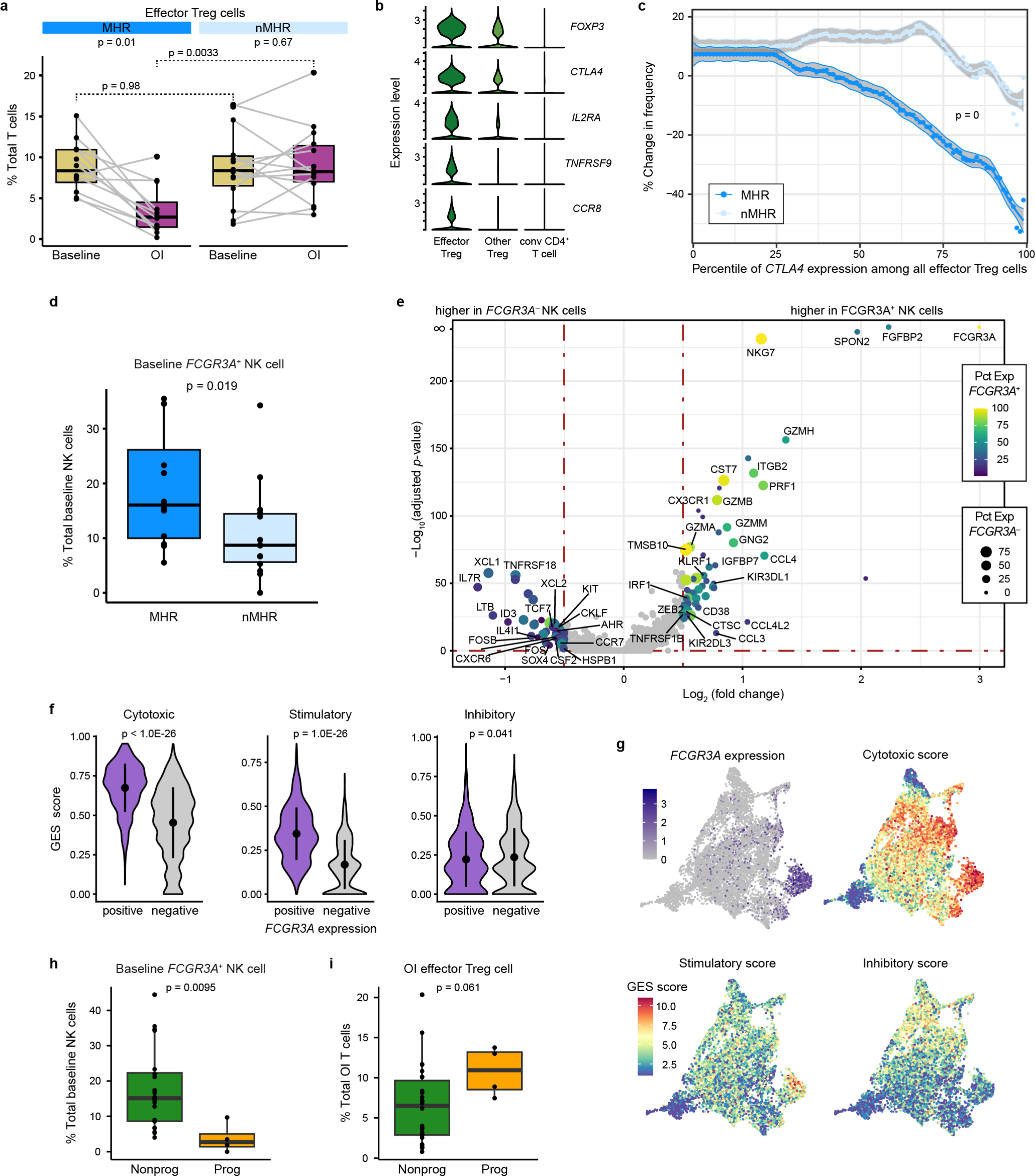
Depletion of effector Treg cells in the TIME associated with response. **a,** Proportion of effector regulatory CD4^+^ T (Treg) cells at baseline and on induction (OI) CTLA-4 and PD-1 immune checkpoint blockade in major histological responders (MHRs; n=12) and in non-MHRs (nMHRs; n=14). Wilcoxon signed-rank test and Wilcoxon rank-sum test used for comparisons within (paired) and between response groups, respectively. *P*-values were false discovery rate adjusted. **b,** Expression of effector Treg markers in effector Treg, other Treg, and conventional CD4^+^ T cells. **c,** Percent change in effector Treg frequency from baseline to OI (y-axis) vs percentile of *CTLA4* expression among effector Treg cells (x-axis) determined for effector Treg populations from MHRs and nMHRs. Shaded area, 95% confidence interval (see **Methods** for *p*-value calculation). **d,** Baseline proportion of *FCGR3A^+^* NK cells in MHRs (n=12) and nMHRs (n=14). **e,** Volcano plot of genes differentially expressed between *FCGR3A^+^* and *FCGR3A^-^* NK cells. Genes related to NK cell function are labeled. y-axis, −log_10_ Bonferroni-adjusted *p*-value from Wilcoxon rank-sum test; x-axis, log_2_ fold change in expression; color scale, percent of *FCGR3A^+^* NK cells expressing gene; dot size, percent of *FCGR3A^-^* NK cells expressing gene. **f,** Violin plots comparing cytotoxic, stimulatory, and inhibitory gene expression signature (GES) scores for *FCGR3A^+^* and *FCGR3A^-^* NK cells (see **Methods** for gene sets). Dot, mean value; vertical line, +/− one standard deviation from the mean. Wilcoxon rank-sum test used. **g,** UMAP of total NK cells (n=8,000) showing *FCGR3A* expression or indicated GES scores. **h,i,** Of 32 patients evaluable for disease progression with scRNA-seq data from either paired baseline-OI (n=26), only baseline (n=2), or only OI samples (n=4), 4 had progression during follow up. **h,** Proportion of *FCGR3A^+^* NK cells at baseline in patients that did not progress (nonprog; n=24) and those that did progress (prog; n=4). **i,** Proportion of effector Treg cells OI in nonprog (n=26) and prog (n=4) patients. **d,h,i,** Wilcoxon rank-sum test used. **a,d,h,i,** Dots, individual patient values; midline, median; box limits, interquartile range; whiskers, 1.5x interquartile range; grey lines, matched baseline-OI sample pairs.

To evaluate the relative contributions of eTreg depletion vs reprogramming to the observed decline, we integrated scRNA-seq data with scTCR-seq data for the CD4^+^ T-cell population. The overwhelming majority of TCR clonotypes in the eTreg cluster at baseline maintained that phenotype on induction, regardless of MHR status: only 3–6% of baseline eTregs shared TCR clonotypes with other CD4 clusters on induction (**Extended Data Fig. 4a,b**). Arguing against a treatment effect, a similar small proportion of TCR clonotypes (<4%) where shared between on-induction eTregs and baseline CD4 clusters (**Extended Data Fig. 4c,d**). Moreover, no changes in eTreg expression of *FOXP3*, *PRDM1*, *HELIOS*, or *EZH2* were observed on induction in MHRs or nMHRs (**Extended Data Fig. 4e-g**). Reprogramming did not appear to explain the marked decline in eTregs in MHRs but not nMHRs. However, eTreg frequency declined on induction with increased *CTLA4* expression, and this was significantly greater in MHRs than nMHRs (**Fig. 4c, Extended Data Fig. 4h**; see **Methods**). Treg depletion *in vitro* is reported to increase as surface CTLA-4 expression increases^34^. Therefore, our data support a dominant role for eTreg depletion in response to iICB in MHRs.

**Fig. 4.**
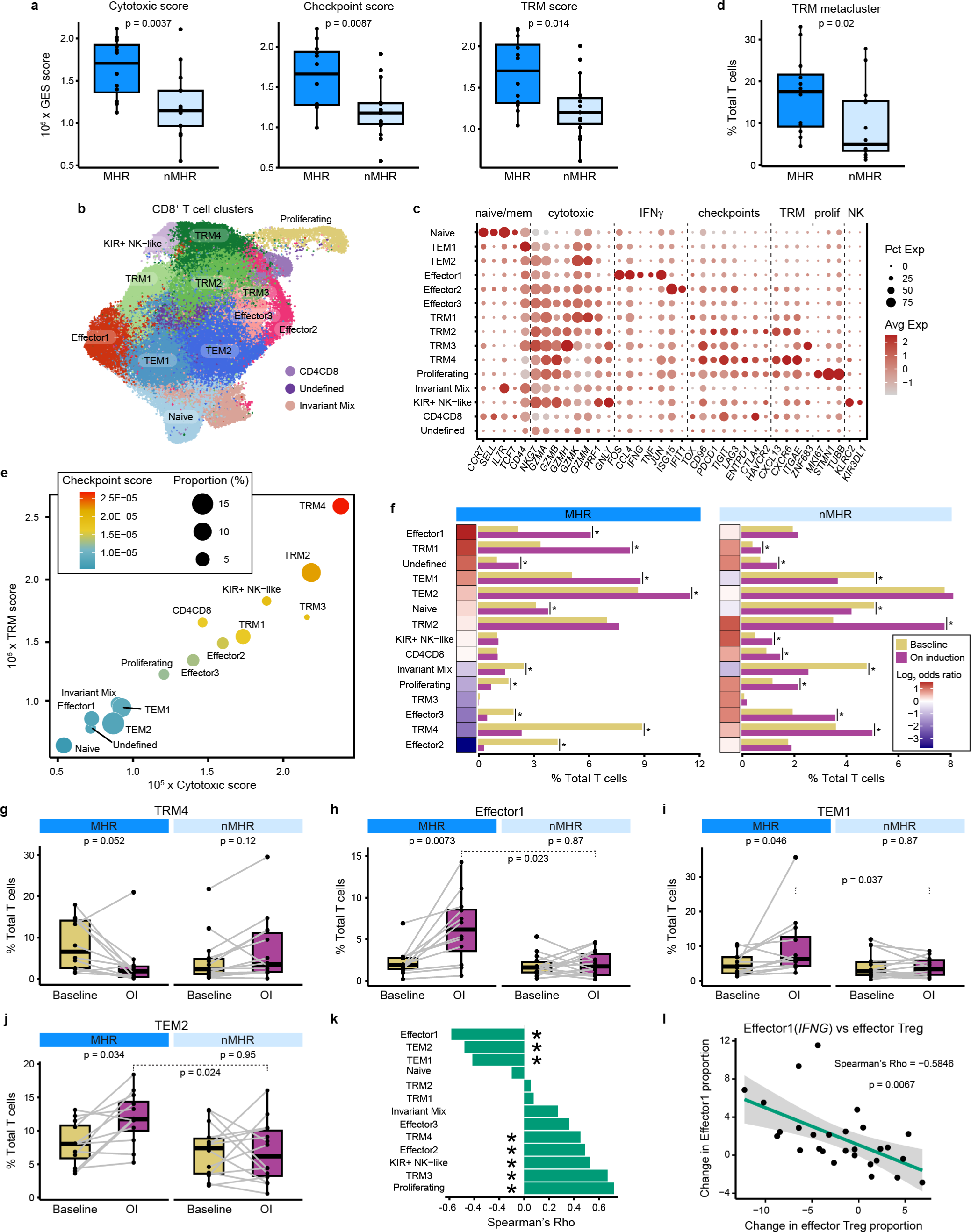
Higher baseline TRM CD8^+^ T-cell proportion and increased *IFNG*-expressing effector CD8^+^ T-cell proportion associated with response. **a,** Cytotoxic (*PRF1, GZMA, GZMB, GZMH,* and *GNLY*), immune checkpoint (*PDCD1, CTLA4, TIM3* [*HAVCR2*]*, LAG3*, and *TOX*), and tissue-resident memory (TRM; *ITGAE* [*CD103*]*, ZNF683* [*HOBIT*]*, ITGA1* [*CD49a*]*, CD69, CXCR6*, and *CXCL13*) gene expression signature (GES) scores for CD8^+^ T cells from major histological responders (MHRs; n=12) or non-MHRs (nMHRs; n=15) at baseline. Wilcoxon rank-sum test used. **b,** UMAP of total CD8^+^ T cells (n=76,319) color-coded by cluster. TEM, effector memory T cell; KIR, killer Ig-like receptor. **c,** Bubble plot showing cluster-level expression of canonical immune genes. Prolif, proliferation; dot size, percent of cells in cluster expressing gene (Pct Exp); color scale, average gene expression level (Avg Exp). **d,** Boxplot comparing proportion of total CD8^+^ T cells in the TRM metacluster (TRM1–4) in MHRs (n=12) vs nMHRs (n=14) at baseline. Wilcoxon rank-sum test used. **e,** Scatter plot showing the average cytotoxic (x-axis) and TRM GES score (y-axis) for each cluster. Color scale, immune checkpoint GES score; circle size, cluster proportion (percent of total CD8^+^ T cells). **f,** Proportion of total CD8^+^ T cells in each CD8^+^ T-cell cluster at baseline and on induction (OI) CTLA-4 and PD-1 immune checkpoint blockade in MHR and nMHR groups. Fisher’s exact test was used to test the change in abundance OI. *, *p*-value < 0.001, false discovery rate (FDR) adjusted. Heatmap, log_2_ of odds ratio of OI proportion to baseline proportion. **g-j,** Proportion of total CD8^+^ T cells in TRM4 (**g**), Effector1 (**h**), TEM1 (**i**), or TEM2 (**j**) at baseline and OI in MHRs (n=12) and nMHRs (n=14). Wilcoxon signed-rank test and Wilcoxon rank-sum test used for comparisons within (paired) and between response groups, respectively. *P*-values were FDR adjusted. **k**, Spearman’s Rho for correlation between change in proportion of each CD8^+^ T-cell cluster and change in proportion of effector Tregs (OI vs baseline) calculated using matched baseline-OI pairs (n=28). *, FDR-adjusted p-value < 0.05. **l**, Scatter plot of change in Effector1 proportion (y-axis) vs change in effector Treg proportion (x-axis; n=28). Shaded area, 95% confidence interval. *P*-value was FDR adjusted. **a,d,g-j,** Dots, individual patient values; midline, median; box limits, interquartile range; whiskers, 1.5x interquartile range; grey lines, matched baseline-OI sample pairs.

### *FCGR3A^+^* NK cells mediate effector Treg depletion

In preclinical murine models, CTLA-4 ICB has been shown to selectively deplete CTLA-4^+^ Tregs via antibody-dependent cellular cytotoxicity (ADCC)^35,34^, but whether CTLA-4 ICB mediates Treg depletion in human subjects remains controversial^36–38^. Given NK cells can mediate ADCC, we profiled 8,000 NK cells in our dataset (**Extended Data Fig. 5a, Supplementary Fig. 5a-c and Supplementary Table 9**). At baseline, MHRs had a considerably higher proportion of NK cells expressing Fc gamma receptor IIIa (*FCGR3A,* encoding CD16) transcripts than nMHRs (**Fig. 3d**). Compared to *FCGR3A*^-^ NK cells, *FCGR3A*^+^ NK cells had higher expression of several cytotoxic mediators, including *PRF1*, *GZMH*, *NKG7*^39^, and *CST7*^40^ (**Fig. 3e, Supplementary Table 10**). To assess *FCGR3A*^+^ NK-cell cytotoxic potential, we examined gene expression signature (GES) scores for stimulatory and inhibitory NK-cell receptors and for cell-mediated cytotoxicity^41^ (**Extended Data Fig. 5b-e**; see **Methods**). *FCGR3A*^+^ NK cells also had higher cytotoxic and stimulatory GES scores vs *FCGR3A*^-^ NK cells (**Fig. 3f,g**).

Importantly, when we explored TIME features in patients stratified by disease progression during follow up, patients who progressed had a significantly lower baseline frequency of *FCGR3A*^+^ NK cells (**Fig. 3h, Supplementary Fig. 6a**). Moreover, patients who progressed maintained a higher frequency of eTregs on iICB (**Fig. 3i, Supplementary Fig. 6b**). Our data therefore suggest that a higher frequency of cytotoxic *FCGR3A*^+^ NK cells in MHRs led to more effective ADCC and eTreg depletion, resulting in improved response to CTLA-4 and PD-1 iICB and reduced risk of cancer progression.

### Baseline CD8^+^ T cells have higher cytotoxicity and frequency of tissue-resident memory cells in MHRs

We compared characteristics of CD8^+^ T cells in MHRs and nMHRs to evaluate potential associations with response and/or eTreg depletion. Comparison of total CD8^+^ T-cell gene expression profiles at baseline revealed significant differentially expressed genes (DEGs; **Extended Data Fig. 6a, Supplementary Table 11**). For example, expression of *CCL5*, a proinflammatory chemokine for NK and other immune cells, was higher in MHRs, whereas expression of Ferritin Heavy Chain 1 (*FTH1*), associated with Treg recruitment and poor OS in HNSCC^42^, was higher in nMHRs. Gene set enrichment analysis (GSEA) revealed that several hallmark immune response pathways (e.g., interferon alpha and gamma response) were enriched in MHRs relative to nMHRs at baseline (**Extended Data Fig. 6b**). Reactome CD28, TCR, and PD-1 signaling pathways were also more enriched in MHRs (**Supplementary Fig. 7a,b**). Moreover, mean baseline CD8^+^ T-cell cytotoxic and immune checkpoint GES scores were notably higher in MHRs (**Fig. 4a, Supplementary Fig. 8a**).

To gain greater insight into CD8^+^ T-cell phenotypes, we performed a clustering analysis that identified 15 clusters (**Fig. 4b,c, Supplementary Fig. 8b-e, and Supplementary Tables 7 and 12**). These included: a naïve or stem-like cluster; two effector memory (TEM) clusters, three effector clusters, and four tissue-resident memory (TRM) clusters. The TEM metacluster comprised TEM1 (*CD44* high) and TEM2 (*GZMK* high), which has been reported to have high potential for inflammatory cytokine secretion^43^. The effector metacluster contained clusters defined by high expression of *TNF* and *IFNG* (encoding interferon gamma, Effector1) or by interferon-stimulating genes (Effector2).

MHRs had a higher baseline proportion of cells within the TRM metacluster than nMHRs (**Fig. 4d, Supplementary Fig. 8f**), consistent with their higher TRM GES score in the total CD8^+^ T-cell population (**Fig. 4a, Supplementary Fig. 8a**). Immunofluorescence imaging demonstrated a trend of higher infiltration by CD45RO^+^PD-L1^+^ CD8^+^ T cells in subsets of MHRs vs nMHRs (**Supplementary Fig. 9a-c**). Immune checkpoint GES score, also an indicator of exhaustion, increased from TRM1 to TRM3 to TRM2 to TRM4, which had the highest score of any CD8 cluster (**Fig 4e, Supplementary Fig. 8g**). TRM4 had very high *CTLA4* and *HAVCR2* expression, consistent with terminally exhausted T cells^43^. TRM clusters had high frequency and abundance of cytotoxic genes (**Fig. 4c**). We note that mean TRM and cytotoxic GES scores for all CD8^+^ T-cell clusters were highly correlated (**Fig. 4e, Extended Data Fig. 6c, Supplementary Fig. 8g,h**). Notably, *CTLA4* expression was much lower in TRM cells than eTregs (**Supplementary Fig. 8i**), decreasing the probability of depletion via ADCC.

Several prior studies have linked TRM CD8^+^ T cells, enriched for tumor antigen-specific TCRs, to ICB response^44–47^. We conclude that MHRs had a higher baseline population of CD8^+^ T cells with an antigen-experienced phenotype in the TIME than nMHRs.

### Change in proportion of CD8^+^ T-cell clusters correlated with Treg depletion

We next investigated changes from baseline in the proportion of total T cells in each CD8^+^ T-cell cluster on iICB. MHRs had more dynamic changes in cluster proportions than did nMHRs (**Fig. 4f, Extended Data Fig. 6d,e**). In MHRs, Effector1(*IFNG*), which has the potential to upregulate MHC class I molecules to enhance antigen presentation in the TIME^48^, had the highest increase in odds on induction. Both TEM clusters also increased significantly, whereas the most exhausted TRM cluster, TRM4, decreased significantly (**Fig. 4f-j**), consistent with contraction after tumor clearance. In contrast, TRM2, TRM4, and the proliferating cluster increased in nMHRs. Notably, the increased frequency from baseline in Effector1(*IFNG*), as well as in TEM1(*CD44*) and TEM2(*GZMK*), correlated significantly with decreased frequency of the eTreg cluster (**Fig. 4k,l**). In preclinical models, expansion of the effector CD8^+^ compartment was observed upon IL2RA^+^ eTreg depletion, due in part to increased IL-2 availability in the TIME^49^. Our data therefore support expansion of effector CD8^+^ T cells in response to *FCGR3A^+^* NK-cell-mediated ADCC of eTregs in the MHR TIME.

### MHRs have higher CD8^+^ T-cell clonality at baseline and clonal dynamics on iICB

Activation of tumor-reactive T cells upon recognition of tumor antigens presented by MHC molecules leads to clonal expansion. To evaluate the TIME TCR repertoire, we integrated scRNA-seq data with scTCR-seq data. CD4^+^ T cells overall had high diversity and low clonality (**Supplementary Fig. 10a-c**). Therefore, we focused on the CD8^+^ TCR repertoire. CD8^+^ T cells in MHRs had a higher degree of baseline clonality than nMHRs, as evaluated by diversity indices and clonotype abundances (**Fig. 5a**, **Extended Data Fig. 7a-d**). TCR clonotypes with high frequency (≥ 1%) mapped predominantly within the TRM metacluster at baseline and on iICB (**Fig. 5b, Extended Data Fig. 7e**), in agreement with the higher baseline antigen-experienced phenotype in the TIME of MHRs than nMHRs. Compared to nMHRs, the proportion of TCR clonotypes that expanded or contracted from baseline was significantly higher in MHRs (**Fig. 5c,d**). Novel clonotypes were also more frequent in MHRs (**Extended Data Fig. 7f**). In both patient groups, TCR clonotypes present at baseline had significantly higher frequency on iICB than did novel clones (**Extended Data Fig. 7g**), favoring a greater role for clonal revival^50^ than replacement^51^.

**Fig. 5.**
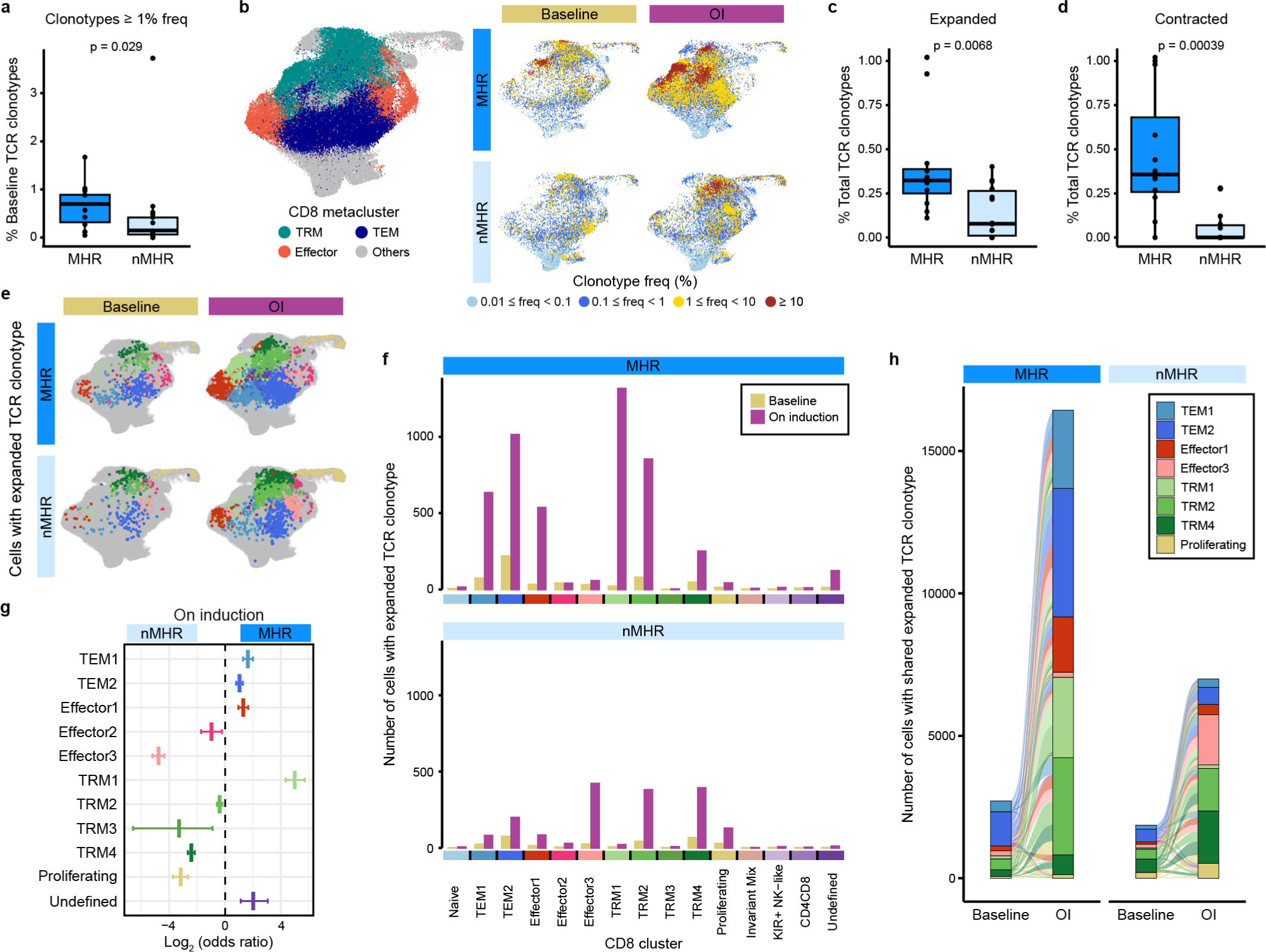
Higher baseline CD8^+^ T-cell clonality and increased clonotype dynamics associated with response. **a,** Fraction of total TCR clonotypes detected at a frequency ≥ 1% at baseline in major histological responders (MHRs; n=12) and in non-MHRs (nMHRs; n=14). **b,** Total CD8^+^ T-cell UMAP (see **Fig. 4b**) showing TEM, effector, and TRM CD8 clusters grouped as metaclusters (left) and the distribution of TCR clonotypes with the indicated frequency ranges at baseline and on induction (OI) dual immune checkpoint blockade (right). **c,d,** Fraction of expanded (**c**) or of contracted (**d**) clonotypes in MHRs and nMHRs, calculated as the number of significantly expanded or contracted clonotypes divided by the total number of unique baseline clonotypes per patient. **a,c,d,** Dots, individual patient values; midline, median; box limits, interquartile range; whiskers, 1.5x interquartile range. Wilcoxon rank-sum test used for comparisons. **e,f,** Distribution of cells with expanded TCR clonotypes among CD8 clusters at baseline and OI in MHRs and nMHRs visualized as projections on the UMAP of total CD8^+^ T cells (**e**) and bar plots (**f**). **g,** MHR:nMHR odds ratio of clonotype expansion in each cluster. y-axis, CD8 cluster; x-axis, log_2_ odds ratio; midline, odds ratio; horizontal line, 95% confidence interval. **h,** Alluvial plots showing the number (bar height) and phenotypic distribution (bar color) of baseline and OI cells that share an expanded TCR clonotype in MHRs or in nMHRs. Links, sharing between clusters; link color, baseline phenotype of shared clonotypes; link thickness, number of cells with shared clonotypes in each connected cluster. To improve clarity, only TEM1/2, effector1/3, TRM1/2/4, and proliferating clusters are shown.

### Phenotypic difference in expanded or contracted clones between MHRs and nMHRs

A total of 78 CD8^+^ TCR clonotypes (47 in MHRs, 31 in nMHRs) expanded significantly in response to iICB and distributed predominantly in the TEM and TRM metaclusters (**Fig. 5e,f and Supplementary Table 13**). In MHRs, cells with an expanded TCR clonotype distributed mostly among TEM1(*CD44*), TEM2(*GZMK*), Effector1(*IFNG*), and the less exhausted TRM clusters (TRM1/3) in contrast to the more exhausted TRM2/4 and proliferating clusters in nMHRs (**Fig. 5f, Supplementary Fig. 11**). Consistent with these observations, the odds of clonotype expansion in MHRs vs nMHRs on induction differed significantly across clusters (**Fig. 5g, Extended Data Fig. 8a**). Novel TCR clonotypes exhibited a preference for Effector1(*IFNG*) and TRM1(*GZMK*) phenotypes in MHRs and exhausted TRM 2/4 clusters in nMHRs (**Extended Data Fig. 8b-d, Supplementary Fig. 11**).

A majority of significantly contracted TCR clonotypes had TRM or Effector1(*IFNG*) phenotypes in MHRs (44 clonotypes) and TRM 4 or TEM 2 phenotypes in nMHRs (13 clonotypes; **Extended Data Fig. 8e-g, Supplementary Fig. 11, and Supplementary Table 13**). The co-occurrence of clonotype contraction and tumor clearance in MHRs is consistent with the T cells being primarily tumor-reactive. In nMHRs, where tumor cells remained detectable in the TIME, fewer instances of clonotype contraction were observed (**Fig. 5d and Extended Data Fig. 8e-h**).

### Greater cell-state transitioning among CD8^+^ TCR clonotypes in MHRs

The distribution of individual TCR clonotypes across several CD8^+^ T-cell clusters indicated possible cell-state transitions (**Fig. 5h, Extended Data Figs. 8h and 9a-c**). To evaluate the effects of iICB on cell-state transitions, we developed a transition index as a quantitative measure for each patient of TCR clonotype sharing between CD8^+^ T-cell clusters before and after iICB (see **Methods**).

MHRs had significantly higher transition indices than nMHRs (**Fig. 6a**). Focusing specifically on clonotypes that expanded or contracted, we noted that expanded clones transitioned predominantly from TEM2(*GZMK*) or TRM2 to Effector1(*IFNG*) and TEM1(*CD44*) in MHRs (**Fig. 6b**). In contrast, transitions from TRM2 and TRM4 to the proliferating cluster predominated among the expanded TCR clonotypes in nMHRs (**Fig. 6c**). Contracted clonotypes in MHRs tended to transition from exhausted to less exhausted TRM clusters and to the Effector1(*IFNG*) cluster (**Fig. 6d**), while in nMHRs, the Effector3 cluster was involved (**Fig. 6e**). Among expanded clonotypes, 36% and 32% underwent significant cell-state transitions (Fisher’s exact test, adjusted p-value<0.05) in MHRs and nMHRs, respectively. For contracted clonotypes, 23% and 0% had significant cell-state transitions (**Supplementary Table 13**).

**Fig. 6.**
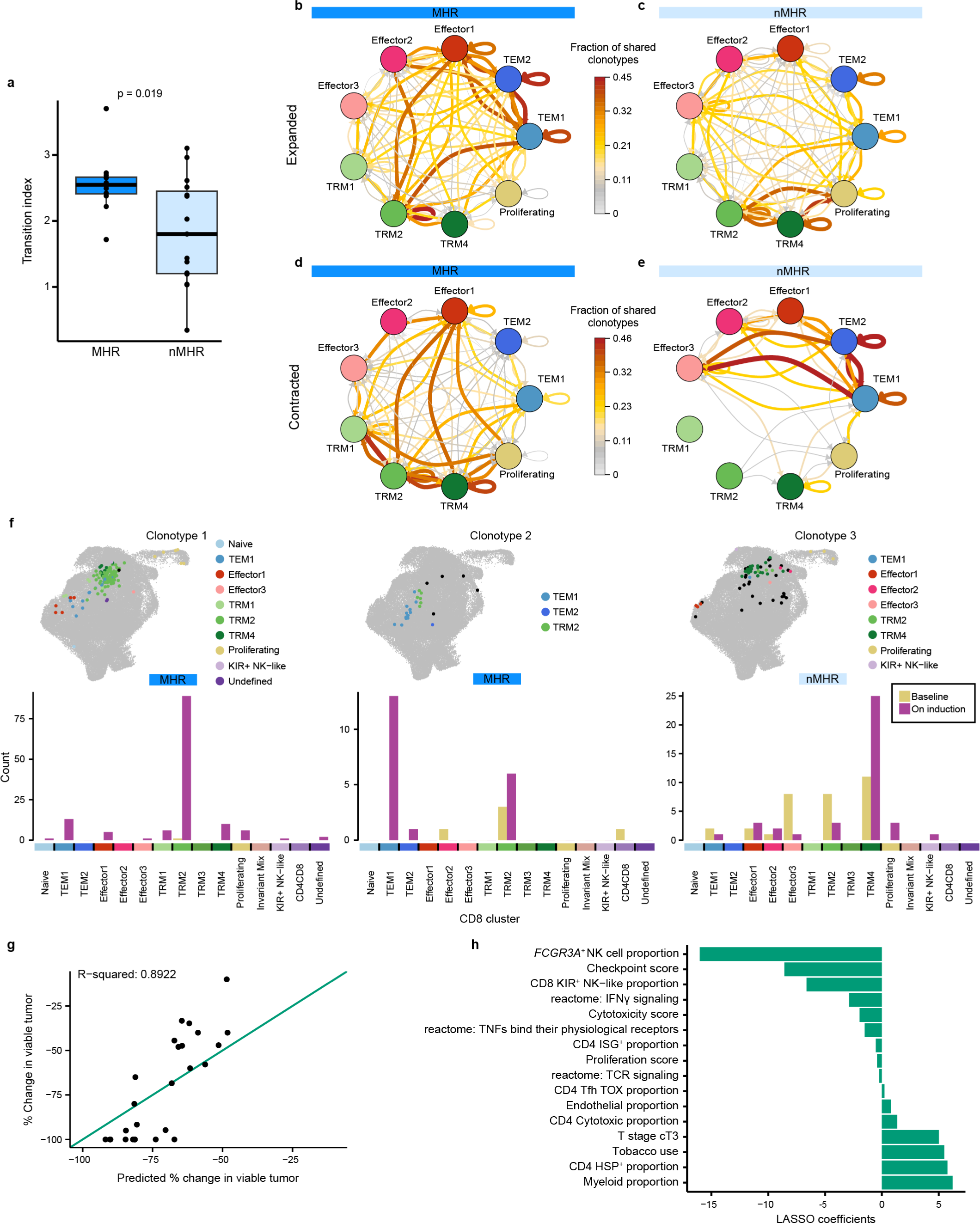
Transition of CD8^+^ T cell clonotypes from TRM to effector memory and *IFNG*-expressing effector cells associated with response. **a,** Boxplot comparing CD8^+^ T cell cell-state transition index in major histological responders (MHRs; n=12) and in non-MHRs (nMHRs; n=15; see **Methods**). **b-e**, Circos plots showing sharing among CD8^+^ T-cell clusters of expanded TCR clonotypes in MHRs (**b**; 47 clonotypes) and nMHRs (**c**; 31 clonotypes) and of contracted TCR clonotypes in MHRs (**d**; 44 clonotypes) and nMHRs (**e**; 13 clonotypes). Arrows indicate direction from baseline state to on-induction state. Color and thickness of arrows indicate the fraction of TCR clonotypes shared by the connected clusters among all expanded (**b,c**) or contracted (**d,e**) TCR clonotypes in MHRs or nMHRs, respectively. To improve network clarity, only TEM, effector, TRM (exclusive of TRM3), and proliferating clusters are shown. Data for all other cell states can be found in Source Data file. **f,** Distribution among CD8 clusters of experimentally validated HPV16-specific TCR clonotypes at baseline and on induction shown as a projection on the UMAP of all CD8^+^ T cells (top) or bar plot (bottom). Black dots, baseline distribution; color-coded dots, on-induction distribution. y-axis, number of cells per cluster; x-axis, CD8 cluster. **g,** Percent change in tumor viability per histological review (y-axis) vs percent change predicted by baseline LASSO regression model (x-axis; n=26). Diagonal, line of identity (i.e., hypothetical scenario where the predicted values perfectly match the actual values). **h,** Features (y-axis; n=16) selected by the baseline LASSO model and corresponding coefficients (x-axis; see **Methods**). Tobacco use, current/former smoker.

Collectively, these data suggested that the *FCGR3A^+^* NK cell-mediated eTreg depletion induced by CTLA-4 ICB facilitated significant clonal expansion and cell-state transitions for the TEM1(*CD44*), TEM2(*GZMK*), and Effector1(*IFNG*) clusters in MHRs. In contrast, cell-state transitions in nMHRs appear to be limited to the exhausted TRM clusters.

### Functional validation of HPV-specificity of expanded T-cell clonotypes

To determine whether expanded TCR clonotypes were HPV-specific, 30 were selected for experimental validation based on HPV16 type, high clonotype frequency on iICB, and a cell-state transition. TCRs were expressed in luciferase reporter Jurkat cells and then stimulated with peptides representing the entire HPV16 proteome (see **Methods**). Three TCR clonotypes demonstrated significant luciferase expression compared to controls in response to 3 distinct HPV16 peptides (**Supplementary Table 13**). Their cell-state distribution at baseline and on iICB were representative of overall differences between MHRs and nMHRs (**Fig. 6f**).

### LASSO regression models associated TIME features with reduction in tumor viability

Our analysis identified several differences in baseline and on-induction features of the TIME in MHRs and nMHRs, including proportions of CD4^+^ and CD8^+^ T-cell and NK-cell clusters, CD8^+^ T-cell GES and immune-related pathway scores, and TCR clonality. To determine which baseline TIME and clinical features (**Supplementary Table 14**) were most influential in predicting treatment response, we built a multivariate LASSO regression model with an outcome of percent reduction in tumor viability. The prediction of the model, which included 16 features, correlated strongly with the observed reduction in tumor viability (R-squared=0.8922, **Fig. 6g,h**). Consistent with our mechanistic hypothesis of eTreg depletion driving response, the most influential factor was the basal proportion of *FCGR3A^+^* NK cells among all NK cells. A separate LASSO model that evaluated change from baseline in TIME features identified percent increase in the Effector1(*IFNG*) CD8^+^ cluster, which correlated inversely with eTreg frequency (**Fig. 4k,i**), as the most influential factor for reduction in tumor viability (**Extended Data Fig. 10**).

## DISCUSSION

Patients with newly diagnosed HPV-positive OPC were responsive to induction CTLA-4 and PD-1 ICB, as demonstrated by a 46% MHR rate. High plasma cfHPV clearance rates (∼71%) in patients with MHR indicated that most became MRD negative during induction. Single-cell profiling of paired tumor biopsy samples in patients with and without MHR revealed a higher baseline frequency of CD8^+^ T cells with an antigen-experienced, TRM phenotype in MHRs. Our data suggest that eTreg depletion by highly cytotoxic *FCGR3A^+^* NK cells via ipilimumab-induced ADCC drives restoration of this antitumor immunity. eTreg depletion was associated with clonotype transition from TRM2 and TEM CD8^+^ T cells toward an Effector1(*IFNG*) phenotype. The preliminary associations we observed between low baseline *FCGR3A^+^* NK-cell frequency, high on-iICB eTreg frequency, and cancer progression support eTreg depletion as important for memory generation and long-term cancer control.

Elucidation of the biological mechanisms contributing to response to iICB will inform strategies to increase MHR rates. To date, the relative contribution of restoration of CD28 co-stimulation vs Treg depletion to the clinical efficacy of ipilimumab in humans has been unclear. Sharma et al reported that intratumoral CD8^+^ T-cell density increased in response to ipilimumab in several tumor types, whereas Treg density did not decline^36^. Potential weaknesses of their analysis include lack of stratification by response and use of FoxP3 alone to identify Tregs. In our analysis, several lines of evidence implicate eTreg depletion by innate effector *FCGR3A^+^* NK cells as critical for response to CTLA-4 and PD-1 iICB. Treg decline was observed only in MHRs, was specific to the eTreg cluster, and increased as *CTLA-4* expression increased. A decrease in 4-1BB^+^ Tregs was similarly associated with MPR to neoadjuvant CTLA-4 and PD-1 in HPV-negative HNC profiled by scRNA-seq^52^. The eTreg population, highly expressive of both *GITR* and *OX40*, may be the Tregs subset that restricts response to PD-1 in lung cancer^53^. Our data are consistent with murine models that showed that higher CTLA-4 expression on Tregs explained their preferential depletion by CTLA4 ICB^54^, and that tumor response was dependent upon Fc competence to mediate ADCC and Treg depletion^34,35,54,55^. Fc engineering of ipilimumab to increase affinity for CD16/FcγRIIIa resulted in more effective Treg depletion with higher expression of CTLA-4 than CD8^+^ T cells^34^. Reprogramming has been proposed to explain decreased eTreg frequency^56^. Treg reprogramming due to decreased TGF-β was implicated in response of HNC to induction bintrafusp alfa^46^, a bifunctional anti-PD-L1 and TGF-β trap. However, the low frequency (∼3%) of TCR clonotype sharing we observed between baseline eTregs and on-induction CD4^+^ T cell clusters was insufficient to explain the ∼60% decline in eTregs in MHRs (**Extended Data Fig. 4**). Decreased Treg frequency was not accompanied by decreased expression of *FOXP3, TGFB1,* or activated Treg markers (e.g., CD25 and 4-1BB) as reported in melanoma after combination therapy^57,58^. We conclude that eTreg depletion rather than reprogramming predominated in MHRs.

MHRs had a significantly higher baseline frequency of *FCGR3A^+^* NK cells capable of mediating ADCC, analogous to improved response to ipilimumab in melanomas with higher tumor-infiltrating *FCGR3A^+^* myeloid cells^37^. HPV-positive cancers have higher infiltration by *FCGR3A^+^* NK cells and higher *CTLA4* expression than HPV-negative HNC^4^. Our multivariable LASSO model indicated that baseline *FCGR3A^+^* NK-cell proportion was the most influential factor in tumor cell reduction. Preclinical models have implicated ADCC by NK cells as a mechanism of response to several antitumor antibodies, including trastuzumab^59^, cetuximab^60^ and obinotumzumab^61^ and shown antibody engagement of FcγRIIIa on NK cells enhances NK-cell degranulation, cytokine production, TNF death receptor expression, and tumor-cell killing^62^. Treg depletion may also enhance direct antitumor killing of tumor cells^63^. Collectively, our findings strongly suggest that the mechanism of action of ipilimumab in MHRs is mediated by depletion of immunosuppressive eTreg cells through NK-mediated ADCC. This novel insight underscores the potential of optimizing ADCC by antibodies targeting intratumoral eTregs, such as Fc-enhanced anti-CD25^64^, anti-CCR8^65^ mABs, and anti-TIGIT^66^.

In our analysis, decline in eTreg frequency correlated with increase in frequency of Effector1(*IFNG*) CD8^+^ T cells, which were most influential in on-iICB tumor viability reduction per LASSO regression. Expansion of Effector1(*IFNG*) and TEM clusters in MHRs is consistent with preclinical studies linking Treg depletion to an increase in T cells with an effector memory phenotype and long-lasting memory^65^. Preclinical models also support increased presence of intratumoral IFNγ^+^ effector T cells in response to combination CTLA-4 and PD-1 ICB vs PD-L1 alone^58^. Based on TCR clonotype sharing across timepoints, our data support a capacity of TRM2 cells to transition toward Effector1(*IFNG*) or TEM cells or towards terminal exhaustion, with Treg depletion (and likely increased IL2 in the TIME) being essential to the former.

Although our scTCR-seq data support cell-state transitions of clonal effector-memory and tissue-resident memory cells toward IFNγ-producing effectors, we cannot rule out the possibility of recruitment of clonotype-identical cells from the circulation. In agreement with prior reports^44,52^, we observed a decrease in TRM cells with a terminally exhausted phenotype (TRM4) in MHRs, suggesting contraction in the absence of antigen stimulation. We cannot exclude the possibility of ipilimumab-mediated clearance by ADCC given these cells highly express CTLA-4. Nevertheless, the increase of cells with a TEM phenotype in MHRs supports the formation of long-lasting anti-tumor memory. Expanded clonotypes that were confirmed to be HPV-specific *in vitro* had memory phenotypes in scRNA-seq data. We note that no MHRs experienced cancer progression.

Although not classified as MHRs, nMHRs nevertheless experienced a 10–80% reduction in tumor viability to iICB. In most neoadjuvant PD-1 ICB trials in HPV-negative HNC, these patients would be classified as responders^24,27,67^. In nMHRs, we observed significant on-induction expansion and TCR clonotype sharing among CD8^+^ T-cell clusters with high frequency and abundance of CTLA-4 and PD-1 expression (i.e., TRM2, TRM4), which also had high cytotoxic and checkpoint scores, consistent with reports that TRM cells are tumor reactive and responsive to PD-1 ICB^44,46^. Our findings concur with a previous scRNA-seq analyses that observed ITGAE^+^ CD8^+^ T-cell expansion in 6 HPV-negative tumors after neoadjuvant PD-1 +/− CTLA4 ICB^44^ but disagree with expansion of the ZNF683^+^ CD8 (i.e., TRM3) in 8 HPV-negative tumors after PD-1 ICB^68^. Given eTreg frequency was not affected by iICB in nMHRs, we conclude that reductions in tumor viability in nMHRs are largely attributable to restored proliferation and effector function of exhausted TRM cells. These transitions are analogous to the progenitor exhausted to exhausted and proliferative exhausted cell-state transitions among CD8^+^ T cells observed in response to PD-1 ICB alone^69^.

Several additional observations from this study should be briefly noted. As observed in melanoma, radiological response underestimated pathological response and should not be used to determine benefit from iICB^70^. Higher response rates reported here vs two neoadjuvant ICB trials with small subsets of HPV-positive OPC^28,29^ may be due to the use of PD-1 ICB alone in one trial and/or our restriction of eligibility to PD-L1 CPS-positive cancers. All progression events occurred in tobacco-exposed patients, consistent with decreased CD8 infiltration^71^ and poor disease control in smokers^1^. This highlights the need for further elucidation of the effects of smoking on the TIME. The 2-year PFS of 84% was like the cisplatin-based chemoradiation arm in RTOG1016^72^, but populations differed. INVAX excluded T4/N3 stage but had more tobacco-exposed patients. Although this study was designed as an alternative approach to therapy rather than as a de-escalation protocol, the XRT plan led to a marked reduction in dose to critical structures (e.g., salivary glands and larynx). Although the grade≥3 adverse event rate was higher during iICB than reported in R/M HNC (37% vs 28.2%)^14^, as expected in patients with intact immune systems^18^, the overall grade≥3 toxicity was less than for chemoradiation in RTOG1016 (66% vs ∼82%)^72^.

Our study has several limitations, including an analysis stratified by histopathology on a biopsy rather than a resection specimen. In melanoma, however, pathological response in an index lymph node is highly concordant with the entire resected specimen^73^ and histopathological features in early-on-treatment biopsies associate with survival^30^. MHR status was associated with cfHPV clearance, a biomarker of reduced risk of cancer progression in HPV-positive OPC^74^. In lung cancer, clearance of cfDNA during neoadjuvant therapy was associated with pathological complete response^75,76^. As with most studies in human subjects, our results are correlative and do not directly demonstrate mechanism. Our TCR validation was limited to HPV16 antigens, and the overall analysis was limited to lymphocytes. Ongoing experiments will assess the reactivity of TCRs to tumor neoantigens. Future analyses will examine the contributions of *FCGR3A*-expressing myeloid cells and germline *FCGR3A* polymorphisms^54,77^ to Treg depletion and investigate associations between non-lymphoid and malignant cell phenotypes with response.

In conclusion, our single-cell profiling of HPV-positive OPC before and on iICB implicates Treg depletion as critical to maximal restoration of effector function. While patients without Treg depletion still benefit from release of CTLA4 and PD-1 inhibition on TRM effector function, Treg depletion appears critical for reshaping of the TIME. Therefore, immunotherapy combinations designed to enhance baseline antitumor immunity (e.g., therapeutic HPV vaccine), NK-cell activation, and eTreg depletion in the TIME have considerable promise for the treatment of newly diagnosed HPV-positive OPC with immunotherapy alone, particularly if associated with less toxicity than observed here.

## Data Availability

All data produced in the present study are available upon reasonable request to the authors.

## ACKNOWLEDGMENTS

The clinical trial was supported by Bristol Myers Squibb. All correlative data were supported by grants to M.L.G. from CPRIT, the Oral Cancer Foundation, and The MD Anderson HPV-related Cancers Moonshot Program; and to X.J. from CPRIT (RP170593); and to K.C. from the NIH (U01CA247760). Dr. Gillison is a CPRIT Scholar in Cancer Research. We thank Cara Haymaker, PhD, Ayse Koksoy, MD, PhD, Khaled Sanber, MD, PhD, Miren Taberna Sanz, MD, Jagan Sastry, PhD, and David E Symer, MD, PhD for helpful discussions. We thank the members of the Translational Molecular Pathology Immune-Profiling Laboratory Moon Shot Platform at MD Anderson for their support. We thank Beatriz Sanchez-Espiridion and Celia Garcia-Prieto for their assistance with sample procurement and inventory and Mei Jiang, Renganayaki Krishna Pandurengan, Saxon Rodriguez, Carolina Sandoval, and Ou Shi for their technical assistance.

## Author contributions

Conceptualization, MLG; Study design, JJL, JP, MLG; Methodology, XJ, BJ, MW, JJ, WX, ERP, BS, JP, KC, MLG; Data Analysis, XJ, NPR, SY, SH, QL, JD, SE, KA, CHL, SL, DL, CHL, KC, MLG; Investigation, BJ, RF, JR, NG, ML, AG, CF, WX, MLG; Resources, MLG; Data Curation, XJ, NPR, KA; Writing-Original Draft, XJ, MLG; Writing-Review & Editing, XJ, NPR, JDD, MC, KC, MLG; Supervision, KC, MLG; Funding Acquisition, KC, MLG.

## Data Availability Statement

The raw sequencing data were deposited in the European Genome Archive (https://ega-archive.org/) with accession number (EGAC50000000088) under controlled access. Data requests will be reviewed by the HPV Virome Consortium within a week after the requester has provided a signed data use agreement form, which requires the requester to agree not to attempt to identify individual subject participants and not to redistribute the data). Requests for data access should be sent to either Keiko Akagi or Maura Gillison.

## Code availability

All code used for analysis is available from GitHub (https://github.com/KChen-lab/INVAX_manuscript).

## METHODS

### Clinical Trial Protocol

Eligible patients had untreated, pathologically-confirmed, AJCC 8^th^ edition Stage I-II (T1-T3N1-N2, T3N0M0)^78^ squamous cell carcinoma of the oropharynx; p16-positive (strong and diffuse nuclear and cytoplasmic staining in ≥70% of tumor cells)^79^ by immunohistochemistry (IHC); formalin-fixed, paraffin-embedded (FFPE) tumor positive for human papillomavirus (HPV) RNA by RNAScope HPV HR8^80^ or HPV DNA by in situ hybridization (ISH)^81^ or fine needle aspirate cytology positive by Roche Cobas^82^; PD-L1 (Clone 22C3, Dako PharmDx) combined positive score (CPS) ≥1 by IHC^83^; Eastern Cooperative Oncology Group performance status 0-1 (asymptomatic or symptomatic but ambulatory)^84^; age ≥ 18 years; measurable disease by RECIST 1.1^85^; and adequate bone marrow, hepatic and renal function. Patients status-post diagnostic tonsillectomy were eligible if RECIST measurable nodal disease was present. Lifetime tobacco exposure and pack-years of cigarette smoking exposure were determined at enrollment. Major exclusion criteria included active autoimmune disease and diagnosis of other invasive malignancy within the prior 3 years.

For eligibility testing, FFPE tumor specimens were tested for p16 expression by immunohistochemistry using a mouse monoclonal antibody (MTM Laboratories, Heidelberg, Germany) visualized with the Ventana XT autostainer using the 1-view secondary detection kit (Ventana). p16 expression was scored as positive if strong and diffuse nuclear and cytoplasmic staining were present in ≥70% of the tumor cells^79^. HPV E6/E7 mRNA expression in FFPE tumor specimens was detected by use of RNAscope ISH Probe HPV HR 8 assay (types 16, 18, 31, 33, 35, 45, 52, 58) and interpreted as described by Bishop et al ^80^. FFPE tumors were evaluated for HPV DNA using the *in situ* hybridization (ISH)-catalyzed signal amplification method for biotinylated probes (Dako GenPoint, Carpinteria, CA)^86^. Specific staining of tumor cell nuclei for HPV in either analysis defined a positive tumor.

Induction dual immune checkpoint blockade (ICB) consisted of intravenous nivolumab at a dose of 3 mg/kg on days 1, 15, and 29 and ipilimumab at 1 mg/kg on day 1 of a six-week cycle. A second six-week cycle was administered concurrently with intensity modulated radiotherapy (IMRT). Acute toxicity was evaluated weekly during therapy using Common Toxicity Criteria for Adverse Events, Version (CTCAE 4.0; ctep.cancer.gov/protocolDevelopment/electronic_applications/ctc.htm). Immunotherapy was delayed or permanently discontinued due to treatment-related adverse events (TRAE) per management algorithms included in the protocol. Immunotherapy was resumed if TRAE resolved to grade ≤1 within 6 (nivolumab) or 12 (ipilimumab) weeks and corticosteroids dose was prednisone ≤10 mg/day.

Computed tomography or magnetic resonance imaging of the neck was performed at baseline and after induction immunotherapy (i.e., cycle 1), and response was determined per RECIST 1.1^85^ by independent review by a study neuroradiologist masked to clinical data. The trial IMRT strategy consisted of both dose and volume reduction. The primary and nodal gross tumor volume (GTV) was based on post-induction volume as assessed by the treating radiation oncologist to allow for GTV volume reduction in response to induction ICB. Primary and nodal disease were assessed separately, based upon categorization as low (primary <3 cm, node <4 cm) or intermediate (primary ≥3 cm, node ≥4 cm) volume. All patients received an initial plan of 36-44 Gy in 20 fractions over 4 weeks (36 Gy to subclinical disease and 40-44 Gy to gross disease) followed by a boost of 5-10 fractions. Primary/nodal disease with complete response received a 10Gy boost in 5 fractions (total dose 50Gy). Low volume primary/nodal disease received 40Gy followed by a 20Gy boost at 2.0Gy per fraction (total dose 60Gy). Intermediate volume disease received 42-44Gy followed by a boost of 21-22Gy at 2.1-2.2Gy per fraction (total 63-66Gy). All patients received dose- and volume-reduced IMRT to at-risk nodal regions to minimize potential effects on T-cell priming. The dose to the elective treatment volume was reduced to 36 Gy in 20 fractions at 1.8Gy per fraction and at-risk 1^st^ echelon nodal stations.

Fluorodeoxyglucose positron emission tomography (FDG-PET) was performed at baseline and 12 weeks after end of IMRT, and metabolic response was evaluated on a five-point scale per The Hopkins Criteria^87^ as complete (1–2), indeterminant (3), or positive (4–5, i.e., concerning for possible residual tumor). Patients with a score of 4–5 underwent further evaluation, including biopsy and surgical resection as appropriate. Patients with an indeterminant score or without histopathological confirmation of residual tumor had a repeat FDG-PET/CT at 6 months after end of IMRT. 6-month complete metabolic response rate is defined as the proportion of patients who had a score of 1-2 at 3 or 6 months.

To assess tumor status, physical exam and imaging studies were performed every three months for two years, every six months through year five, and then annually.

The protocol was registered with the National Cancer Institute (NCT03799445) and approved by the institutional review board (UT MD Anderson Cancer Center). All patients provided written informed consent. Identification numbers assigned to enrolled patients are known only by the research team; neither the patients themselves nor hospital staff outside of the research team know specific number assignments.

### Clinical trial design and statistical analysis

The study design included a safety evaluation of the treatment regimen in the first 8 enrolled patients. This safety lead-in had a principal outcome of dose limiting toxicity (DLT), defined as: any ≥grade 3 CTCAE V4 TRAE that did not resolve to ≤grade 1 within 28 days; radiotherapy delay of >1 week due to TRAE; radiotherapy discontinuation due to TRAE; or ≥grade 3 radiation mucositis that did not resolve to <grade 1 within 28 days of end of radiotherapy. The period of observation for DLT was 28 days from end of radiotherapy. The study would proceed to phase 2 in the event of ≤2 DLTs or be discontinued in the event of >2 DLTs. With a cohort of 8 patients, the probability of the combination immunotherapy being judged to be too toxic when the true toxicity rate is 45% or higher is at least 78%. If the true toxicity rate is 20% or lower, the probability that the therapy will be safe is 80%.

The primary endpoints of the phase 2 study were to estimate the 6-month complete metabolic response rate by PET and the two-year progression-free survival (PFS) rate. The original non-inferiority design had a sample size of 170 and 82% power to establish non-inferiority of the 6-month complete response rate when compared to that expected from cisplatin-based chemoradiotherapy (0.87), but funding was discontinued by the sponsor at a planned interim analysis after 35 patients were enrolled. Based on the recommendation of the UT MD Anderson Investigational New Drug Office, which served as the data safety monitoring board, two enrolled patients, one with a major protocol deviation (proton therapy) and one who was found upon review to be ineligible (presence of metastasis at enrollment), were replaced. Therefore, the co-primary endpoints are reported here for 35 evaluable patients out of 37 enrolled. Progression-free survival (PFS) was defined as time from start of protocol therapy to death or first documented relapse, categorized as local-regional (primary site or regional nodes) failure (LRF) or distant metastases (DM). OS was defined as time from start of protocol therapy to death. OS and PFS rates were estimated by Kaplan-Meier method. The 95% pointwise CI was obtained by applying the logarithmic transformation to the Kaplan-Meier estimate of the progression-free survival function and the variance of the Kaplan-Meier estimate was calculated using the Greenwood’s formula.

Secondary endpoints included acute and chronic toxicity per CTCAE. Descriptive statistics were used to summarize patient characteristics and toxicity events. Fisher’s exact test or Wilcoxon sign-rank test were used for comparisons between groups for dichotomous and continuous variables, respectively. Spearman’s correlation coefficient was used to evaluate relationships between two continuous variables. Analyses were performed in STATA 13.0, SAS 9.4, or R 4.1.2.

### Biospecimen collection and analysis

Hematoxylin and eosin-stained slides from FFPE at baseline and on induction were evaluated by immune-related pathological response criteria (irPRC)^30,88^ by a head and neck pathologist masked to patient response data. Percent residual viable tumor was calculated as the viable tumor area/total tumor bed area (in mm^2^), whereby the total tumor bed was the sum of the regression bed (characterized by lymphoid infiltrate, proliferative fibrosis, and neovascularization), residual viable tumor, and necrosis. Major histological response was defined as <10% residual viable tumor on induction. Percent reduction in tumor viability was calculated as the baseline tumor viability minus on induction viability/baseline tumor viability.

Whole blood was collected at baseline, days 1, 15, and 29 of cycles 1 and 2, and at 8 and 12 weeks after end of IMRT. Plasma was separated by standard operating procedure and stored at −80°C. Missing samples for tumor or blood-based biomarkers are attributable to the COVID emergency.

HPV type in tumors was detected by ddPCR in DNA purified from FFPE by proteinase K digestion, phenol-chloroform extraction, and ethanol precipitation. cfDNA was purified from 4 ml plasma by use of the QIAsymphony DSP Circulating DNA Kit (Qiagen, Hilden, Germany) on the QiAsymphony SP with the circDNA_2000_DSP or circDNA_4000_DSP program, eluted in 60 µl of AVE buffer, and stored at −20°C. cfHPV DNA was quantified in copies per ml plasma by use of a digital droplet PCR assay with a lower limit of detection of 5 copies and lower limit of quantification of 16 copies per ml of HPV 16, 18, 31, 33, 35, 39, 45, 51, 52, 56, 58, 59, 68 and ERV3 as positive control (Michael T Wotman, Weihong Xiao, Robyn R Du, Bo Jiang, Suyu Liu, Maura L Gillison. Development and validation of an assay to quantify plasma cell-free human papillomavirus DNA for 13 high-risk types that cause 98% of HPV-positive cancers. Submitted). cfHPV clearance was defined as time from cycle 1 day 1 to two consecutive negative tests (i.e., <5 copies/ml plasma) and was visualized by Kaplan-Meier method.

The primary tumor or an involved lymph node underwent core needle biopsy (2 mm X 4) at baseline and on induction (cycle 1, day 15–22 or 36–42). Two cores were submitted for FFPE and two were immersed in ice-cold RPMI 1640 with 10% FBS and transferred to the laboratory for immediate dissociation^89^. After mincing to <1 mm^3^, tissue was placed in RPMI 1640 with 1 mg/ml collagenase A and 0.4 mg/ml hyaluronidase and incubated at 37°C for 1–2 hours. After trypsin digestion, the suspension was filtered through a 70-μm strainer and centrifuged at 450xg for 5 min. Red blood cells were removed when present by ammonium chloride solution. After resuspension in PBS with 0.04% BSA and passage through a 40-μm cell strainer, cells were counted and resuspended at 700–1200 cells/µl to ensure >70% viability. Baseline and on-induction tumor samples from two patients treated on protocol but subsequently excluded from clinical trial analysis (and replaced) due to protocol violations (indeterminant baseline pulmonary nodules subsequently confirmed by biopsy as metastases or receipt of proton therapy) were included in the single-cell data for analyses comparing patients with and without major histological response.

### Single-cell library generation and sequencing

Single-cell capture, barcoding, and preparation of gene expression and T-cell receptor (TCR) libraries were performed as recommended using a Chromium Next GEM Single Cell 5’ library preparation kit (PN-1000006/PN-1000165) and a Chromium Single Cell V(D)J Reagent Kit (PN-1000005), respectively, with V1/V1.1 chemistry reagents (10x Genomics). We loaded approximately 8000 single cells per sample onto the 10x controller. The final libraries containing barcoded single-cell transcriptomes and TCRs were pooled together at 10:1 ratio and sequenced at 150-bp paired-end sequencing on the Novoseq 6000 system (Illumina) aiming for 100–200 million read pairs per sample. A minimum of 5,000 read pairs/cell was sequenced for TCR libraries, and a minimum of 20,000 read pairs/cell for gene expression libraries. We performed additional sequencing run(s) for gene expression libraries that did not initially reach the desired sequencing coverage of >20,000 read pairs/cell.

### Single-cell RNA-seq data processing and clustering

Quality control metrics indicated that single cell transcriptome libraries were sequenced at ∼272 million reads per library and 43,000 reads per cell, at 70% sequencing saturation on average. Raw sequence reads were aligned to a custom reference genome, and gene expression levels were quantified using cellranger-multi (version 6.0.0) with default parameters. The custom reference genome included both human genome reference (GRCh38 primary genome) and 13 high-risk HPV genomes (PaVE database, https://pave.niaid.nih.gov). Gencode human gene annotation database (release v32) was used for transcript quantification. Each single-cell V(D)J library was sequenced at greater than 5 million read pairs.

In addition to the basic filtering performed by Cell Ranger, cells with low quality data were filtered from analysis if the number of detected genes was <250 and number of reads detected was <350. The upper limits of these two matrices were sample dependent (**Supplementary Fig. 1**). Cells were further filtered if the proportion of mitochondrial gene counts was higher than 15%. Genes including TCR variable genes, immunoglobulin variable genes, ribosomal protein coding genes, high abundance lincRNA genes (e.g., MALAT1) were excluded from downstream analysis (hereafter, blacklist genes).

### Single-cell RNA-seq data clustering and cell type determination

The cell by gene count matrices were further processed using Seurat package (v4.3.0)^90^. The standard protocol in Seurat was used to normalize the expression level of genes in each cell and to identify the 2,000 most highly variable genes using ‘*NormalizeData*’ and ‘*FindVariableFeatures*’ function. The blacklist genes were not included in the 2,000 most highly variable genes. After scaling and centering of the gene expression matrices using ‘*ScaleData*’ function, we performed PCA based on the 2,000 most variable genes and dimension reduction using ‘*RunUMAP*’. To perform clustering, the shared nearest neighbor (SNN) graph was first constructed with ‘*FindNeighbors*’ and then clusters were determined using Louvain algorithm by ‘*FindClusters*’ with resolution at 0.6 and projected on the UMAP plot. We used canonical marker genes, e.g., *PTPRC, CD3E, CD8A, CD4, FOXP3, MS4A1, GNLY, MZB1, LYZ, KIT, COL1A1,* and *PLVAP* to label major cell types in each cluster, including T, B, plasma, myeloid, fibroblast, and endothelial cells (**Supplementary Fig. 2d**). To confirm the accuracy of cell-type labeling, we ran SingleR package for each sample using the CHETAH reference dataset, which contains four tumor types: colorectal, breast, melanoma, and head and neck^91^. Cell-type annotations from these methods largely agreed for most cell types, while rare discrepancies were reconciled by further inspection of marker genes in the clusters. A single cluster, which expressed markers for multiple cell types consistent with the presence of doublets, was excluded, resulting in a total of 367,451 cells for further analysis. Except for tumor cells, all clusters shown on the UMAP plot were cell-type specific and not patient specific, indicating minimal-to-no batch effects at this level of clustering analysis.

### Identification of malignant cells

We combined epithelial score, HPV gene expression, and inferred copy number aberration (CNA) to identify malignant cells in each sample. Because malignant cells in HPV-positive oropharyngeal cancer have an epithelial lineage, we first calculated an epithelial score using the mean expression level of epithelial genes *KRT5*, *KRT6A*, *KRT6B*, *KRT13*, *KRT14*, *KRT15*, *KRT16*, *KRT17*, *KRT18*, and *KRT19*, selected using the method described by Puram, et al^92^. HPV gene expression was readily available from the scRNA-Seq profile. To infer gene copy number from gene expression profiles, we performed CopyKAT (v1.0.8)^89^ with the default parameters for 10x data. CopyKAT output included a normal/aneuploid label for each cell along with the copy number profile. Because sparsity of a scRNA-Seq profile often leads to drop-out of epithelial markers and HPV genes and may introduce noise in the inferred CNA, we labeled cells at the cluster level rather than directly labeling individual cells. For each sample, fine resolution clustering was first performed using ‘*FindClusters*’ with resolution set at 1.0. Cells in a cluster were then classified as malignant if the cluster met two out of three criteria: 1) >50% cells expressed HPV genes; 2) >50% of cells were aneuploid; or 3) >50% cells had positive epithelial scores.

### T-cell clustering analysis

We extracted CD8^+^ T and CD4^+^ T cells, including regulatory T (Treg) cells from the full dataset for further analysis. Silhouette score^93^ was used to evaluate batch effects in the integrated dataset without correction. Alternative results were obtained by applying several batch correction methods including reciprocal principal-component analysis and Harmony^94^. Silhouette score results indicated little difference across various batch correction methods. To avoid over-correction, we used the integrated dataset without correction for further analysis^95^. Clustering analysis was then performed for CD8^+^ T cells and CD4^+^ T cells separately as described above using the first 20 principal components and clustering resolution of 0.6. This analysis yielded 15 and 14 clusters for the CD8^+^ and CD4^+^ T-cell compartments, respectively. The distributions of CD8 and CD4 clusters in each sample indicated no batch effects in the two datasets (**Supplementary Figs. 4b,c and 8c,d**).

### Phenotyping of T-cell clusters

Marker genes for each cluster were identified from a list of differentially expressed genes found by ‘*FindAllMarkers,*’ which uses a two-sided Wilcoxon rank-sum test (**Supplementary Figs. 4d and 8e, Supplementary Tables 8 and 12**). We created bubble plots for each cluster showing expression of canonical immune markers^96,97^. In the end, we integrated all information to define the functional states for each T-cell cluster in collaboration with immunologists on the study team.

### Analysis of the change in frequency of cells with *CTLA4* expression level greater than different expression percentiles

We modeled the response variable *y* (percent reduction in the fraction of effector Treg cells expressing *CTLA4* above certain levels in x-axis) as nonlinear functions of *x* (0 to 99^th^ percentile of *CTLA4* expression level among all effector Treg cells) using a generalized additive model (GAM)

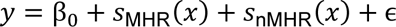

with Gaussian error ε ∼ *N*(0, σ^2^). The cubic smoothing spline for each patient group *p* ∈ {MHR,nMHR} can be represented as a linear combination of *K* cubic basis functions,

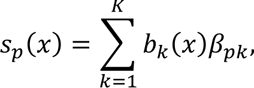

where the cubic basis functions *b*_*k*_(*x*) are enforced to be the same for all patient groups. This enables one to test differential patterns between patient groups by comparing the smoothing spline parameters β_*pk*_ using a Wald test.

The model was fitted using R package mgcv, and the 95% pointwise confidence bands for the fitted model were estimated as 2 standard errors above and below the fitted model^98^.

To test differential patterns between patient groups, we constructed a hypothesis test based on the work of Van den Berge at al^99^. We tested the null hypothesis that all smoothing spline coefficients within the same knot are equal, i.e., *H*_0_: β_MHR,*k*_ = β_nMHR,*k*_ for all *k* ∈ {1, …, *K*}. This null hypothesis is represented as *H*_0_: ***C***^*T*^**β** = 0, where ***C*** is a contrast matrix of size 2 × *K* whose column corresponds to a contrast between β_MHR,*k*_ and β_nMHR,*k*_. We test the null hypothesis using the following Wald test statistics,

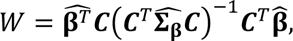

where 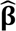 denotes the estimator of **β**, 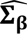 is an estimator of the covariance matrix **Σ_β_** of **β**. The *p*-value is calculated based on the asymptotic distribution of the Wald test statistics *W*, i.e., the chi-squared distribution with degrees of freedom equal to the column rank of ***C***.

### Quantification of gene set activity scores

The GSDensity algorithm was used to generate gene expression signature (GES) scores for gene sets for each cell using transcriptomic data. GSDensity is a cluster-free method tailored to quantify the collective activity level of a given gene signature/set from sparse single-cell or spatial molecular count data^100^. The immune checkpoint GES score included *PDCD1, CTLA4, TIM3 (HAVCR2), LAG3*, and *TOX* expression. The tissue-resident memory GES score included *ITGAE (CD103), ZNF683 (HOBIT), ITGA1 (CD49a), CD69, CXCR6*, and *CXCL13* expression^101^. *PDCD1* was excluded to avoid overlap with the immune checkpoint gene set. The CD8^+^ T-cell cytotoxic GES score included *PRF1, GZMA, GZMB, GZMH,* and *GNLY* expression, which were also in the NK-cell cytotoxic GES score, along with *GZMM, FASLG,* and *NKG7*^41^. GES scores for inhibitory receptors (*KIR2DL3, KIR2DL4, KLRC1, TIGIT,* and *CD96)* and for stimulatory receptors (*FCGR3A, NCR3, KLRF1, KLRK1,* and *KLRC2)* were also determined for NK cells. Alternatively, GES scores were calculated using the AUCell algorithm^102^.

### Single-cell TCR-seq data analysis

Filtered contigs annotated each T cell contig with high confidence and were mapped to scRNA-seq data using shared cell barcodes. A total of 155,187 cells with V(D)J information was detected among all 65 samples. Among them, 84.21% of cells contained sequence data for both TCR alpha and beta chains, 14.16% contained only TCR beta sequence data, and 1.63%, only TCR alpha chain data. In total, 131,954 cells (85.03%) with TCR sequence data were mapped to T cells identified by scRNA-seq. TCR clonotype was determined for 80.75% of CD8^+^ T cells and 89.45% of CD4^+^ T cells. We used the amino acid sequences of both the alpha- and beta-chain CDR3 regions for clonotype identification in downstream analysis.

### Analysis of TCR clonotype expansion

To identify clonotypes with a significant change in frequency from baseline to on-induction timepoints, we applied Fisher’s exact test. *P*-values were corrected for multiple testing using false discovery rates for each patient-specific paired sample, after exclusion of clonotypes detected only once. Clonotypes with adjusted *p*-value <0.05 were considered statistically significant. Clonotypes were further stratified into expanded clonotypes (those with significant increase on induction), contracted clonotypes (those with significant decrease on induction), and novel clonotypes, which were not detected at baseline.

### Evaluation of T-cell state transition

To measure the degree of phenotypic transition of a TCR clonotype, we first calculated a transition index by adopting the Kullback–Leibler (KL) divergence. We defined the transition index *Trans_t_* for clonotype *t* as:

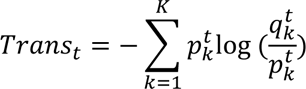

where 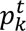 is the ratio of the number of cells with TCR clonotype *t* in cluster *k* over the total number of cells with TCR clonotype *t* at baseline, while 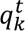 is the ratio of the number of cells with TCR clonotype *t* in cluster *k* over the total number of cells with TCR clonotype *t* on induction. *K* is the total number of cell clusters. The metric is similar to the STARTRAC method^103^, although rather than using Shannon entropy, KL divergence was used to measure the relative entropy between two probability distributions to reflect the distributional difference or distance.

We then aggregated transition indices across all clonotypes to quantify T-cell state transition at the patient level:

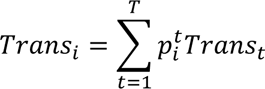

where 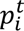 is the ratio of the number of cells within clonotype *t* over the total number of cells for patient *i* at baseline. *T* is the total number of clonotypes.

To evaluate whether the cluster distributions of a TCR clonotype at two timepoints were significantly distinct, we applied Fisher’s exact test for each TCR clonotype. A contingency table (2 rows, 15 columns) representing the number of cells for a TCR clonotype across CD8^+^ T-cell clusters at baseline and on induction was generated for the test.

### Multivariate regression model

To identify features of the tumor immune microenvironment (TIME) associated with percent reduction in tumor viability on induction, we fitted a LASSO (Regression Shrinkage and Selection via the Lasso) regression model using the R “glmnet” package^104^, where the optimal value of the regularization parameter, lambda, was determined via a ten-fold cross-validation. After exclusion of one patient with less than 100 T cells in the baseline sample, LASSO regression model fitting was limited to the 26 remaining patients with paired baseline-on induction data. For the baseline model, regression covariates (p=95; **Supplementary Table 14**) included patient clinical characteristics (e.g., age, gender, tobacco smoking status [never, former/current], and tumor and nodal stage) and features curated from scRNA-seq and scTCR-seq data collected before treatment. Tumor stage features contain N and T stages, which were converted to dummy variables via one-hot encoding. To assess treatment effects, we also developed another LASSO model (herein referred to as “delta model”) that included both baseline and on-induction features. For the delta model, regression covariates (p=87) included the difference of each TIME molecular feature between on-induction and baseline (p=85) and two additional features: the number of expanded and number of contracted clones. All regression covariates were standardized to have a mean of 0 and a variance of 1 prior to fitting the LASSO regression model to avoid coefficient scaling issues. Features that had non-zero coefficients were deemed to have linear associations with percent reduction in tumor viability from baseline. LASSO regression identified 13 baseline (baseline model) and 18 changed (delta model) features that were independently associated with reduction in tumor viability.

### Multiplexed barcode fluorescent imaging

Multiplexed barcoding staining was performed on 4-µm thick FFPE sections using a PhenoCycler (Akoya Biosciences) with antibodies against pan-cytokeratin (CK; clone AE1/AE-3; Akoya Biosciences), CD45 (clone D9M81; Akoya Biosciences), CD3e (clone EP449E; Akoya Biosciences), CD8 (clone C8/144B; Akoya Biosciences), CD45RO (clone UCHL1; Akoya Biosciences), and PD-1 (clone D4W2J; Akoya Biosciences) and with DAPI (4′,6-diamidino-2-phenylindole) as previously described^105^. Entire tissue sections were imaged at 20x magnification using an inverted fluorescence microscope (Keyence BZX800). All images were processed using an Akoya Processer computer and QPTIFF image storage. A pathologist selected the entire tumor area using the QuPath 4.04 image analysis software to capture various elements of tissue heterogeneity. All the data codes were process in R studio 4.1.2 and consolidated using phenoptr 0.3.2 and phenoptrReports 0.3.3 packet (Akoya Biosciences).

### Quantification of T cell and T-cell subtype proportions in fluorescent images

Major types and subtypes were annotated based on marker co-expression, reported with pathologists’ manual supervision. For major cell types, CD45 and CD3e identified T cells, and CK identified malignant cells. The combination of CD45/CD3e/CD8/PD1/CD45RO was used to identify antigen-experienced effector memory T cells. For each sample section, cells with low detection fidelity (cellular area <10 or >1000 um^2^, nuclear area <5 or >80 um^2^, or detection probability less than 0.75) or with unidentifiable type (e.g., not positive for any markers) were excluded from quantification. All analysis were performed using R 4.2.1.

### Identification of HPV16 epitopes recognized by expanded TCR clonotypes

The full-length paired TCR alpha/beta sequences for a selection of CD8 TCR clonotypes that expanded significantly from baseline were determined from the 5’ V(D)J single sequencing data derived from tumors of patients with HPV-positive OPC. Specifically, the data output in the AIRR file format was used. The human constant region of the TCR was replaced with the mouse counterpart^106^, then synthesized and cloned into the pMSGV3 retroviral backbone (Genscript). Expression of the transgenic TCRs in CD8^+^ Jurkat cells without an endogenous TCR and modified to express luciferase upon TCR engagement was performed via electroporation. Briefly, ∼6 million Jurkat (TCRαβ-KO) CD8^+^ cells (Promega) with cell viability > 90% were transfected with program CL-120 and 10 μg of TCR-pMSGV3 in a 100 µl single Nucleocuvette™ using SE Cell Line 4D-Nucleofector Kit and LONZA 4D-Nucleofector™ System. TCR expression was confirmed by flow cytometry after 24 hours. Then, Jurkat cells with TCR expression were co-cultured with partially or fully HLA-matched CD3-depleted peripheral blood mononuclear cells, obtained via negative selection using magnetic beads (Miltenyi), and either CEFX or HPV16 peptide pools. The CEFX peptide pool comprises MHC class I-restricted peptide epitopes from common viruses (cytomegalovirus, Epstein-Barr virus, and influenza virus); thus, most people should have T cells that react to this pool. The HPV16 peptide pools was designed in house. Briefly, a set of peptides covering the entire HPV16 proteome was generated using peptides 15 amino acids (aa) in length that overlapped by 11 aa. The peptides were assigned to pools in a 96-well plate per the following criteria: 1) two peptides may be pooled together only once; 2) a peptide may not be pooled with another peptide if it has already been pooled with either neighboring peptide; and 3) each peptide should be present three times. After 6 hours co-culture, cells were processed for luciferase assay using a T Cell Activation Bioassay (TCRαβ-KO) kit (Promega) per maufacturer’s protocol, and the plates were read on a Synergy H1 microplate reader (Biotek). For HPV16, the designed peptide pool matrix allows for direct demultiplexing and identification of the 15mer that gave rise to a response. Furthermore, in some cases, it allows for direct identification of the 11mer containing the minimal epitope, if two neighboring 15mers both give a positive signal. For CEFX, no demultiplexing is needed.

### Additional statistical analysis

All the statistical analyses were performed using R. To compare the abundance of T cell clusters across timepoints or response groups, Fisher’s exact test was used to calculate p values. For cell fraction comparison at patient level, Wilcoxon rank-sum was used to compare across response groups, Wilcoxon signed-rank was used to compare paired samples. Spearman’s correlation coefficient was used to evaluate relationships between two continuous variables. For cell level GES score comparison, Wilcoxon rank-sum test was used considering the number of cells. Unless specified, all statistical testing was two-sided and significance level cutoff was 0.05. An FDR correction was applied for all comparisons with multiple statistical tests.

### Data oversight

Study design and implementation, data collection, analysis, and interpretation, and manuscript preparation were performed by the authors. The authors had complete access to all data. The first (XJ) and last two authors (KC and MLG) serve as guarantors of all analyses and manuscript content. The clinical trial sponsor (BMS) did not participate in data analysis or manuscript preparation.

---

**Extended Data Fig. 1.**
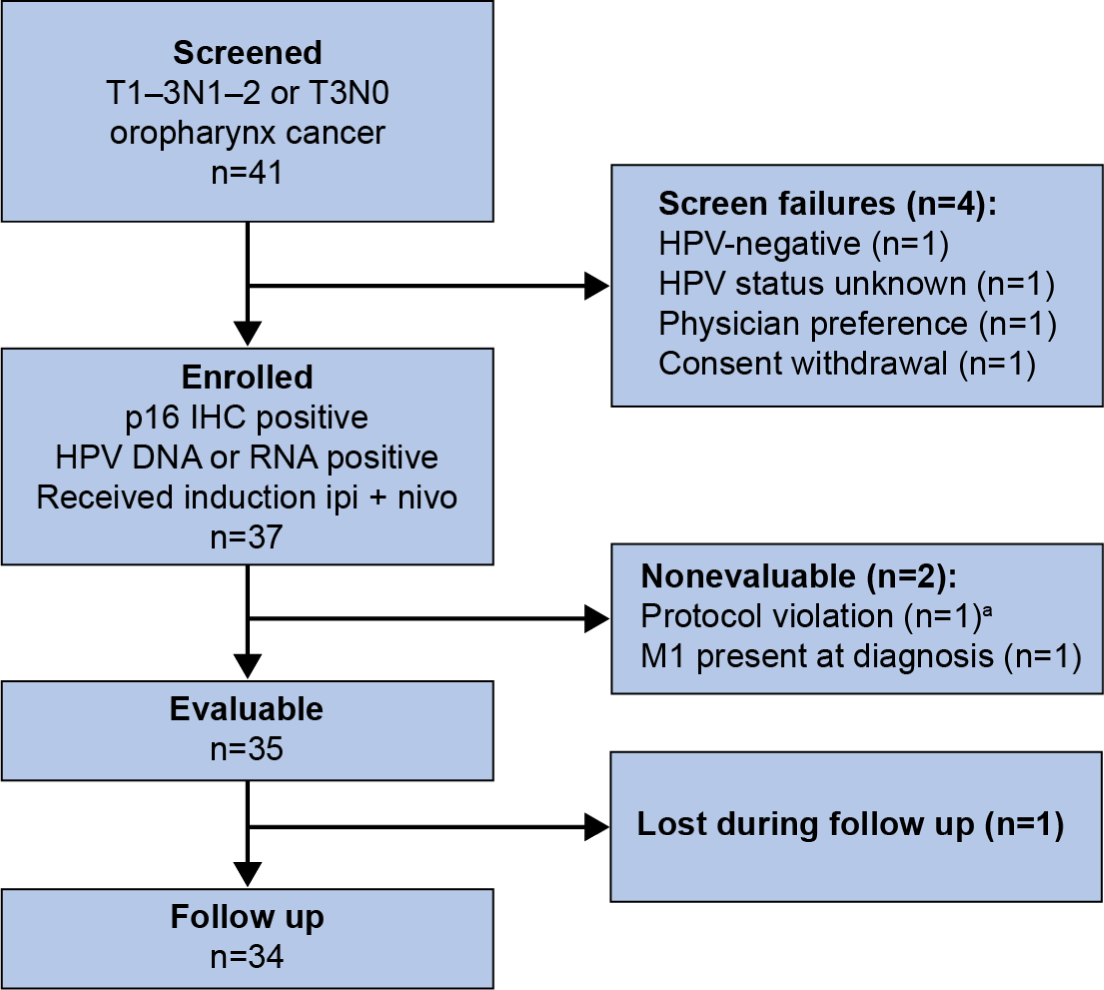
CONSORT flow diagram. Flow diagram shows the disposition of patients throughout the study phases. Reasons for screen failures or nonevaluable status are indicated. ^a^Patient received proton radiotherapy instead of intensity modulated radiotherapy.

**Extended Data Fig. 2.**
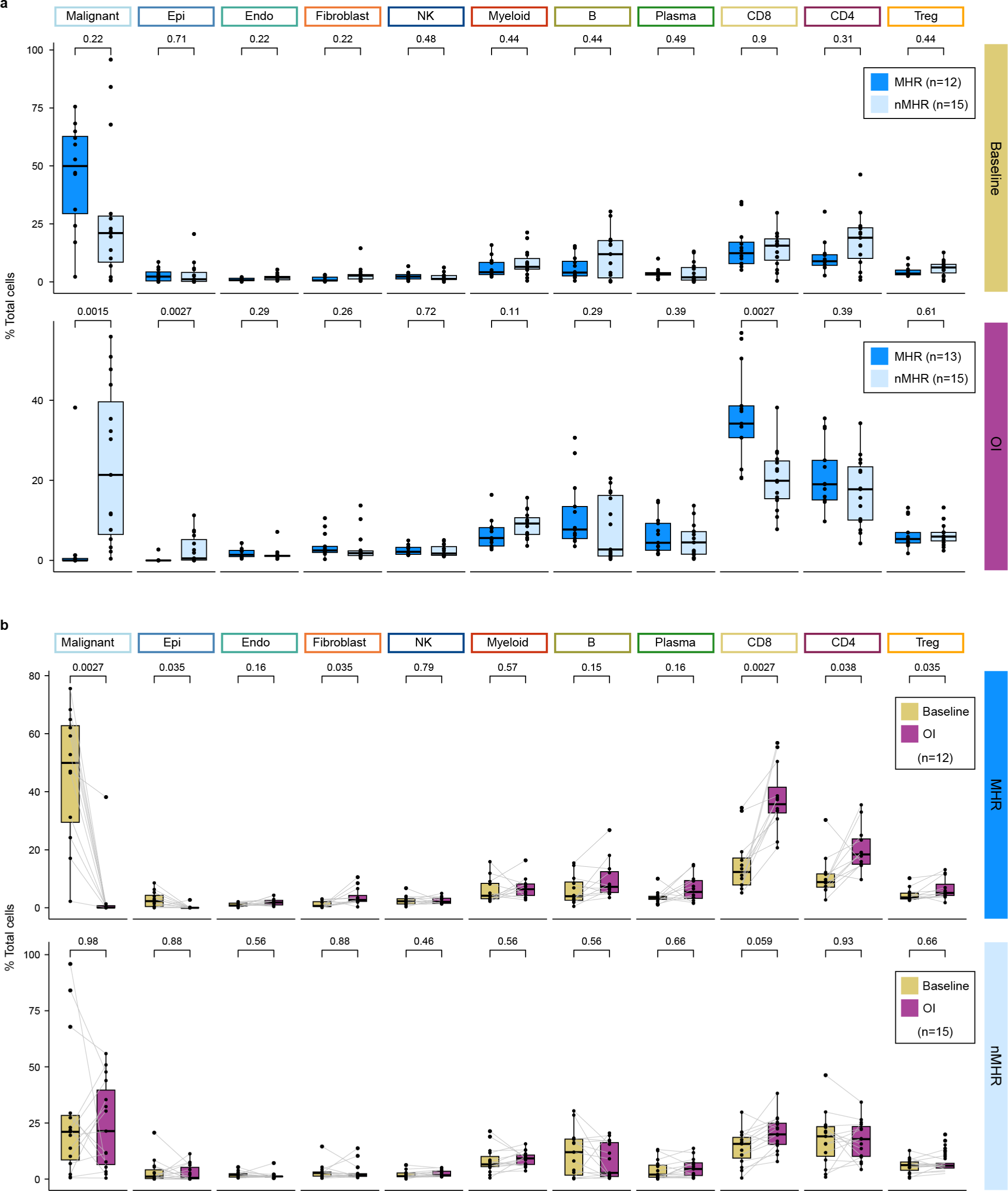
Proportions of cell types in the tumor immune microenvironment stratified by histological response and time. **a,** Proportion of indicated cell type in major histological responders (MHRs) and in non-MHRs (nMHRs) compared at baseline (MHRs, n=12; nMHRs, n=15) and on induction (OI) CTLA-4 and PD-1 immune checkpoint blockade (MHRs, n=13; nMHRs, n=15). Wilcoxon rank-sum test was used. **b,** Proportion of indicated cell type at baseline and OI compared in MHRs (n=12) and in nMHRs (n=15). Wilcoxon signed rank test was used. **a,b,** Dots, individual patient values; midline, median; box limits, interquartile range; whiskers, 1.5x interquartile range; grey lines, matched baseline-OI sample pairs. *P*-values were false discovery rate adjusted.

**Extended Data Fig. 3.**
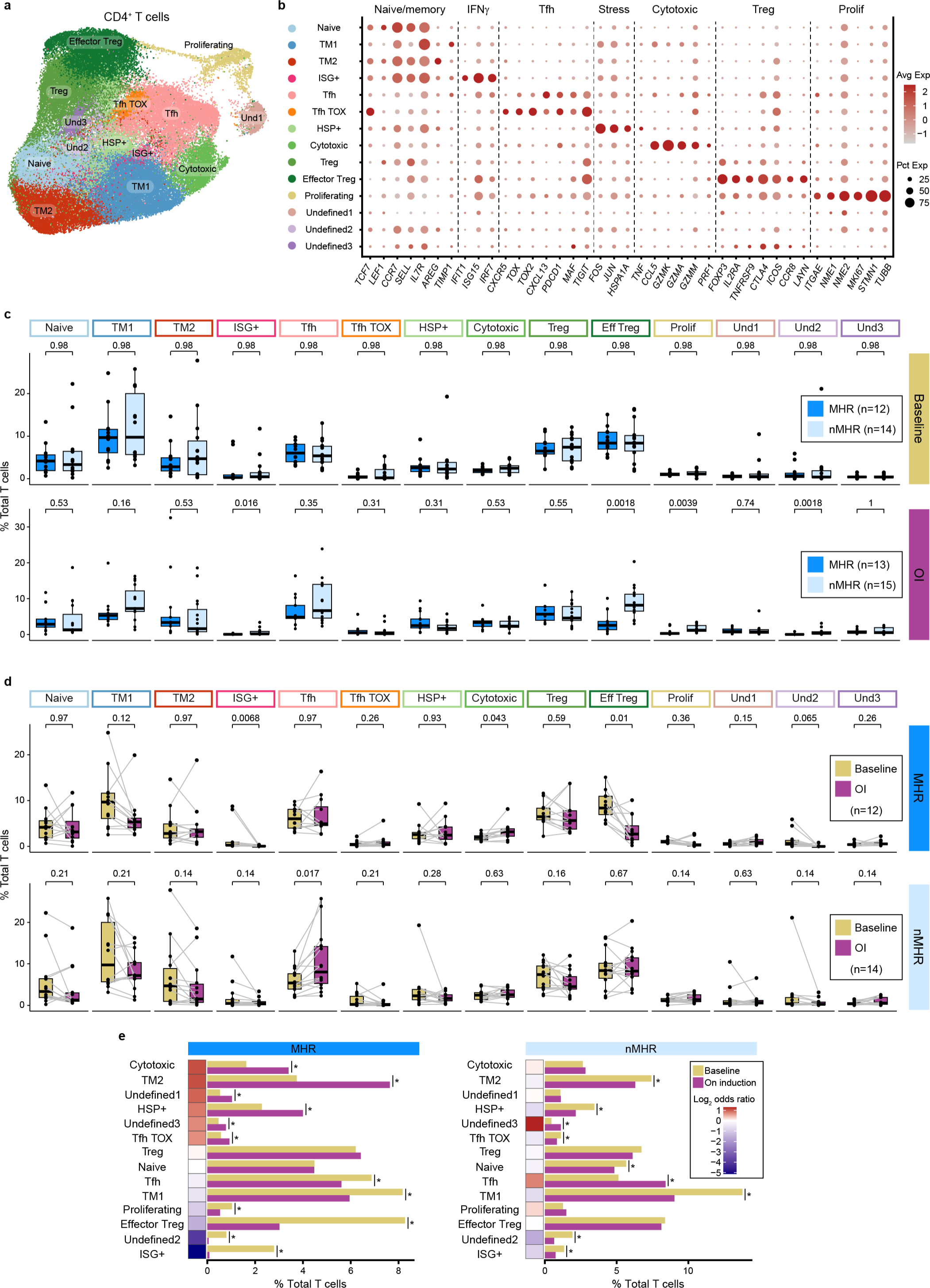
Characterization of CD4^+^ T cells identified in the HPV-positive OPC tumor immune microenvironment at baseline and on induction dual immune checkpoint blockade. **a**, UMAP of total CD4^+^ T cells (n=78,622) color-coded by cluster. TM, T memory; ISG+, interferon-stimulated genes positive; Tfh, follicular helper T cell; Tfh TOX, *TOX*-positive Tfh; HSP+; heat shock protein positive; Treg, regulatory T cell; Und, undefined. **b**, Bubble plot showing expression of naïve/memory T cell, interferon gamma signaling (IFNγ), Tfh, stress, cytotoxic, Treg, and proliferating (prolif) genes in each cluster. Avg Exp, average expression level of gene in cluster; Pct Exp, percent of cells in cluster that express gene. **c**, Proportion of each CD4 cluster in major histological responders (MHRs) and in non-MHRs (nMHRs) compared at baseline (MHRs, n=12; nMHRs, n=14) and on induction (OI) CTLA-4 and PD-1 immune checkpoint blockade (MHRs, n=13; nMHRs, n=15). Wilcoxon rank sum test was used. **d**, Proportion of each CD4 cluster at baseline and OI compared in MHRs (n=12) and in nMHRs (n=14), respectively. Wilcoxon signed-rank test was used. **c,d,** Dots, individual patient values; midline, median; box limits, interquartile range; whiskers, 1.5x interquartile range; grey lines, matched baseline-OI sample pairs. *P*-values were false discovery rate (FDR) adjusted. **e**, Proportion of total T cells in each CD4 cluster at baseline and OI in the MHR group and in the nMHR group. Fisher’s exact test was used to test the change of abundance upon treatment. *, FDR-adjusted *p*-value < 0.001; heatmap, log_2_ of OI:baseline odds ratio.

**Extended Data Fig. 4.**
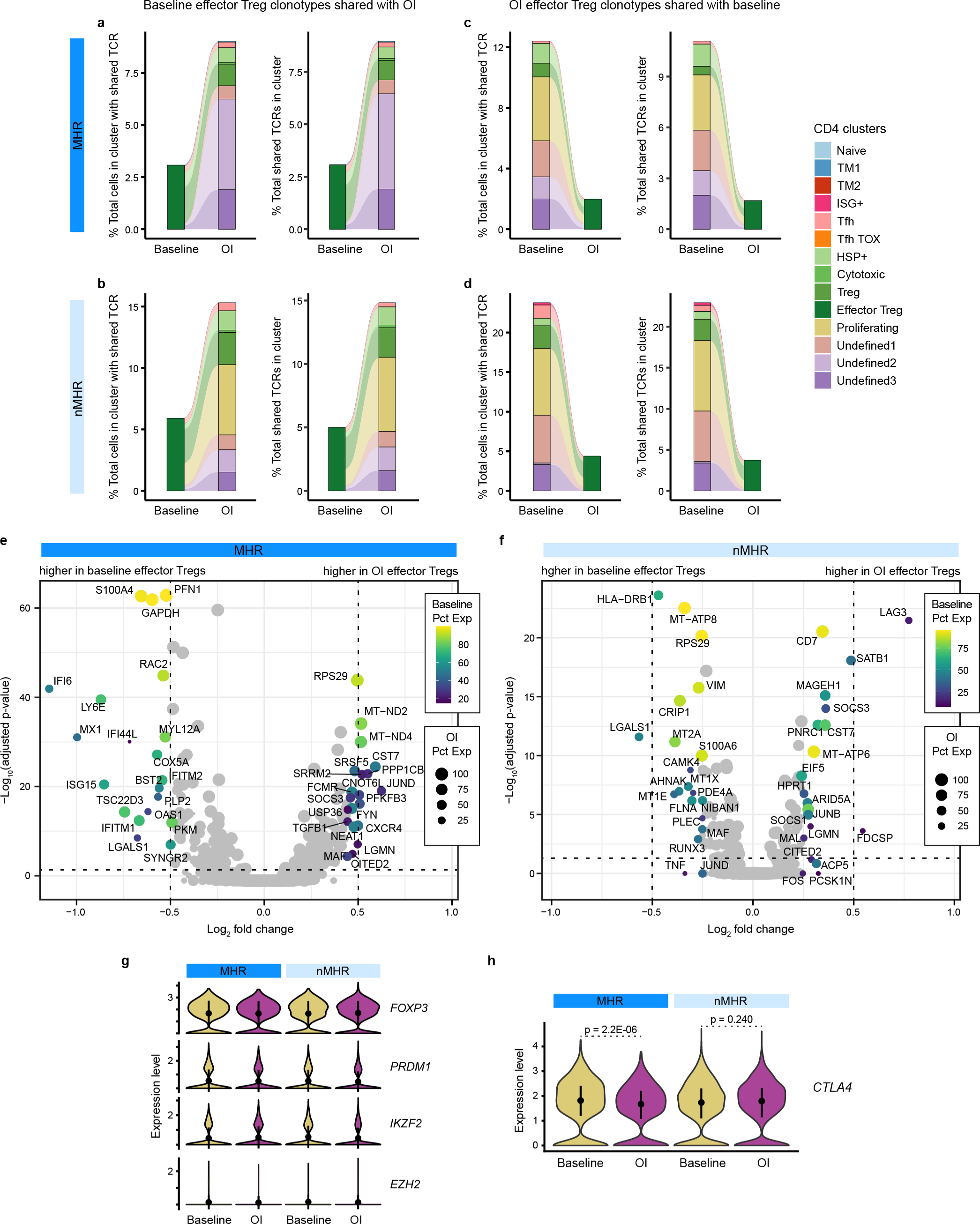
Minimal sharing of TCR clonotypes contradicts reprogramming as the main mechanism of decreased effector Treg frequency. Integration of scTCR-seq and scRNA-seq data for all samples enabled tracking of TCR clonotypes across timepoints and CD4 clusters. **a,b,** Alluvial plots of TCR clonotypes shared between the effector Treg cluster at baseline and CD4 clusters exclusive of effector Tregs OI in MHRs (**a**) and nMHRs (**b**). **c,d**, Alluvial plots of TCR clonotypes shared between CD4 clusters exclusive of effector Tregs at baseline and the effector Treg cluster OI in MHRs (**c**) and nMHRs (**d**). **a-d,** Left plot, proportion of cells (bar height) in a CD4 cluster (bar color) from baseline/OI samples that share a TCR clonotype with the connected clusters from the other timepoint, expressed as percent of the total number of cells in the cluster that share a TCR clonotype with cells from the other timepoint, including cells from the same cluster, which for clarity, are not shown. Right plot, proportion of TCR clonotypes (bar height) in a CD4 cluster (bar color) from baseline/OI samples that are shared with the connected clusters from the other timepoint, expressed as percent of the total number of TCR clonotypes shared with the other timepoint, including clonotypes shared with the same cluster, which for clarity, are not shown. Links, sharing between baseline and OI CD4 clusters; link color, cluster sharing with effector Treg cluster; link thickness, percent of cells with shared clonotype or of shared clonotypes in a cluster. **e,f,** Volcano plot of genes differentially expressed between baseline and OI effector Treg cells in MHRs (**e**) and nMHRs (**f**). y-axis, −log_10_ Bonferroni-adjusted *p*-value from Wilcoxon rank-sum test; horizontal dashed line, p = 0.05; x-axis, log_2_ fold change in expression; color scale, percent of baseline effector Treg cells expressing gene; dot size, percent of OI effector Treg cells expressing gene. **g,h,** Violin plots comparing expression of transcription factors that regulate effector Treg phenotype (**g**) and of *CTLA4* (**h**) at baseline and OI in MHRs and in nMHRs. Dot, mean value; line, +/− one standard deviation from the mean. *P*-values were determined by Wilcoxon rank-sum test.

**Extended Data Fig. 5.**
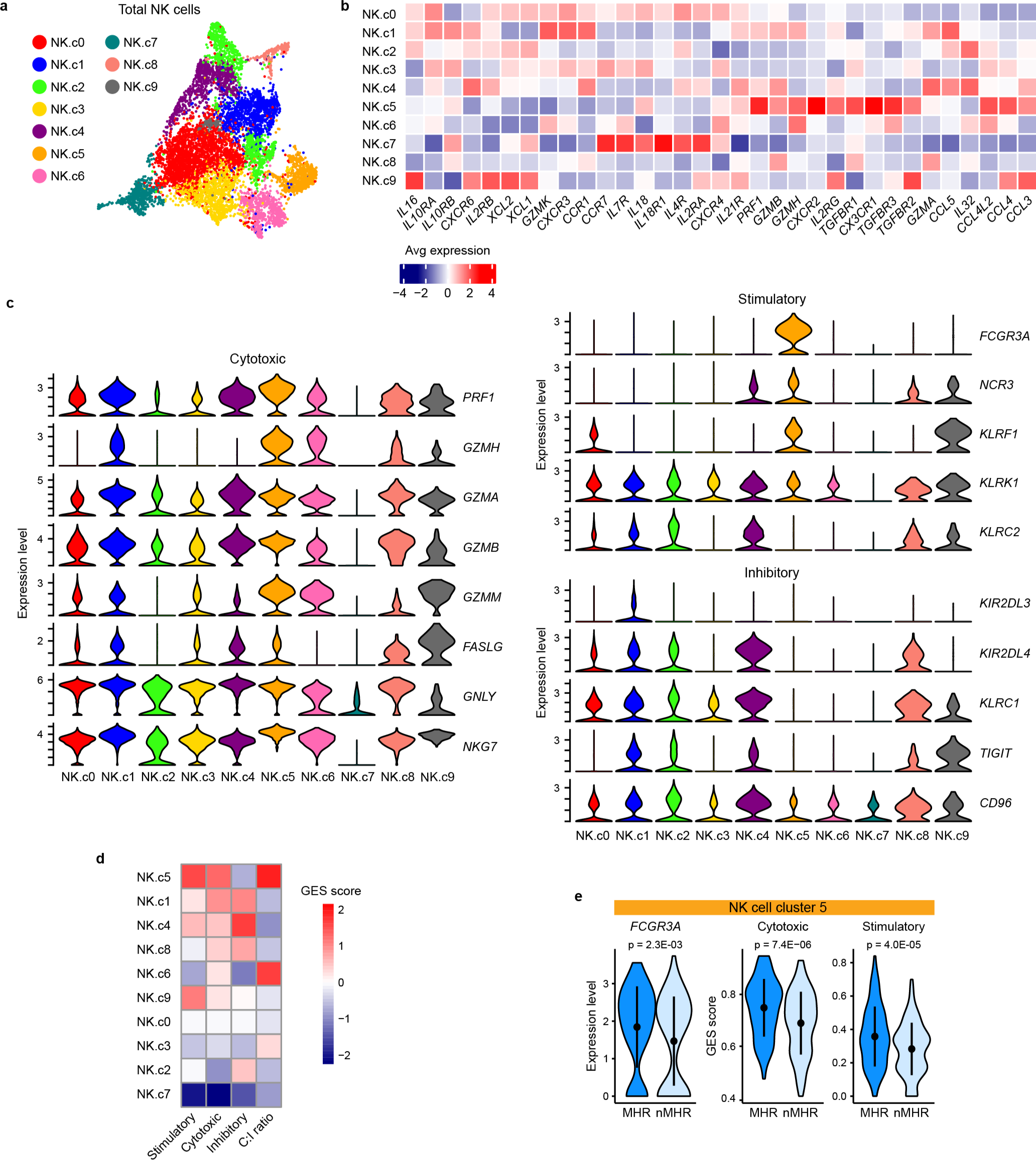
Characterization of NK cells identified in the HPV-positive OPC tumor immune microenvironment at baseline and on induction dual immune checkpoint blockade. **a**, UMAP of total intratumoral NK cells (n=8,000) color-coded by cluster. **b,** Heatmap of average expression of NK functional genes described by Tang et al^107^ in each NK-cell cluster. **c,** Violin plots showing expression of cytotoxic (left), stimulatory (upper right), and inhibitory (lower right) signature gene sets in each cluster. **d,** Heatmap showing the cytotoxic, stimulatory, and inhibitory gene expression signature (GES) score for each cluster and the ratio of cytotoxic score to inhibitory score (C:I ratio). **e,** Comparison of baseline Fc gamma Receptor IIIa (*FCGR3A)* expression and cytotoxic and stimulatory GES scores in cluster 5 NK cells from major histological responder (MHR) and non-MHR (nMHR) groups. Dot, mean value; line, +/− one standard deviation from the mean. *P*-values were determined by Wilcoxon rank-sum test.

**Extended Data Fig. 6.**
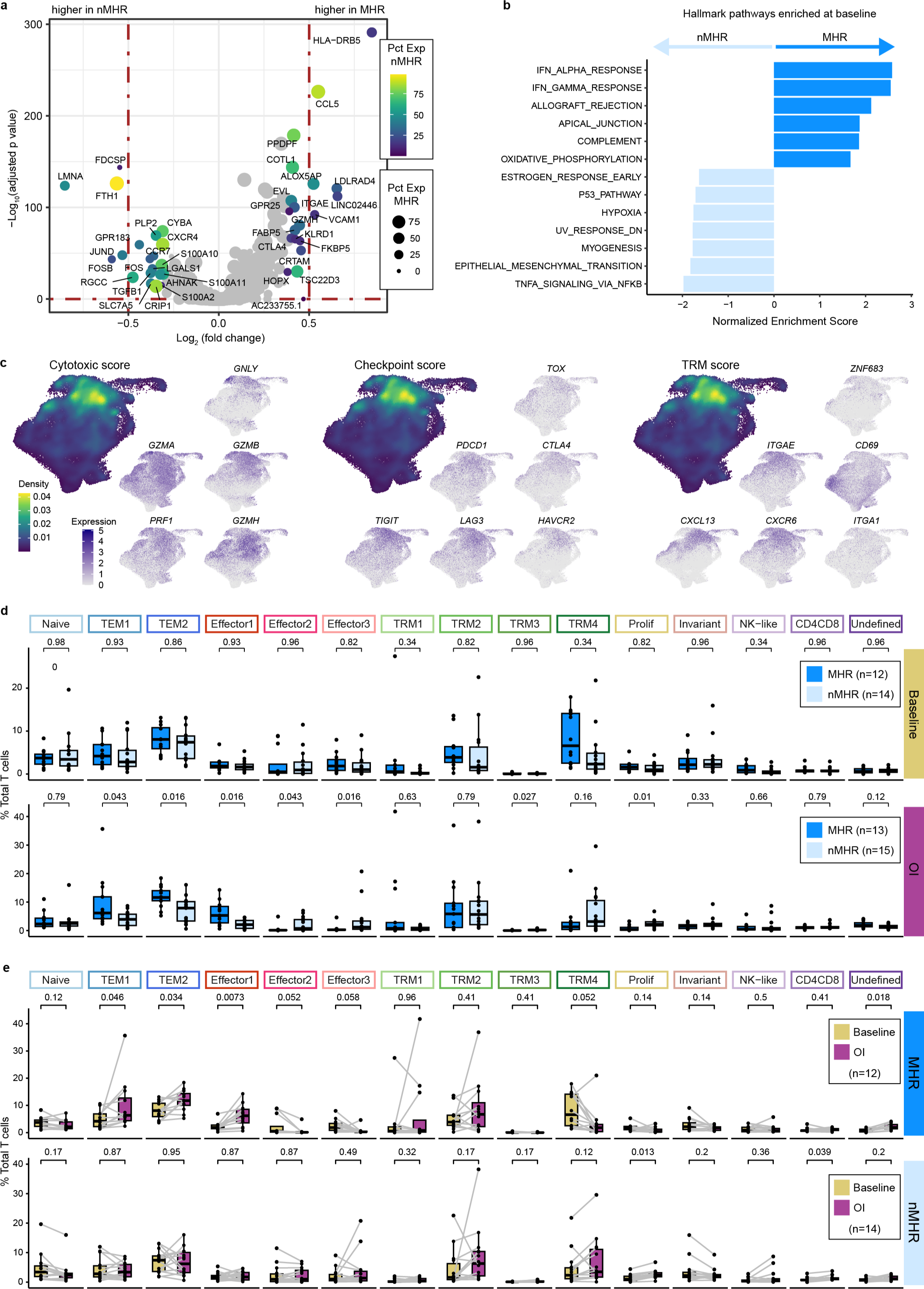
Characterization of CD8^+^ T cells identified in the HPV-positive OPC tumor immune microenvironment at baseline and on induction dual immune checkpoint blockade. **a,** Volcano plot of genes differentially expressed between baseline CD8^+^ T cells from major histological responders (MHRs) and those from non-MHRs (nMHRs). y-axis, −log_10_ Bonferroni adjusted *p*-value from Wilcoxon rank-sum test; x-axis, log_2_ fold change in expression; color scale, percent of CD8^+^ T cells expressing gene in nMHRs; dot size, percent of CD8^+^ T cells expressing gene in MHRs. **b,** Hallmark gene pathways differentially enriched between total baseline CD8^+^ T cells in MHRs and those in nMHRs. IFN, interferon; DN, down. **c,** Projection of cytotoxic (*PRF1, GZMA, GZMB, GZMH,* and *GNLY*), immune checkpoint (*PDCD1, CTLA4, TIM3* [*HAVCR2*]*, LAG3*, and *TOX*), and tissue-resident memory (TRM; *ITGAE* [*CD103*]*, ZNF683* [*HOBIT*]*, ITGA1* [*CD49a*]*, CD69, CXCR6*, and *CXCL13*) gene expression signature (GES) score and of expression level of genes within each GES on the UMAP of all CD8^+^ T cells. Density plots were used to show GES score distribution. **d**, Proportion of each CD8 cluster in MHRs and in nMHRs compared at baseline (MHRs, n=12; nMHRs, n=14) and on induction (OI) CTLA-4 and PD-1 immune checkpoint blockade (MHRs, n=13; nMHRs, n=15). Wilcoxon rank-sum test was used. **e**, Proportion of each CD8 cluster at baseline and OI compared in MHRs (n=12) and in nMHRs (n=14), respectively. Wilcoxon signed-rank test was used. **d,e,** Dots, individual patient values; midline, median; box limits, interquartile range; whiskers, 1.5x interquartile range; grey lines, matched baseline-OI sample pairs. All *p*-values were false discovery rate adjusted.

**Extended Data Fig. 7.**
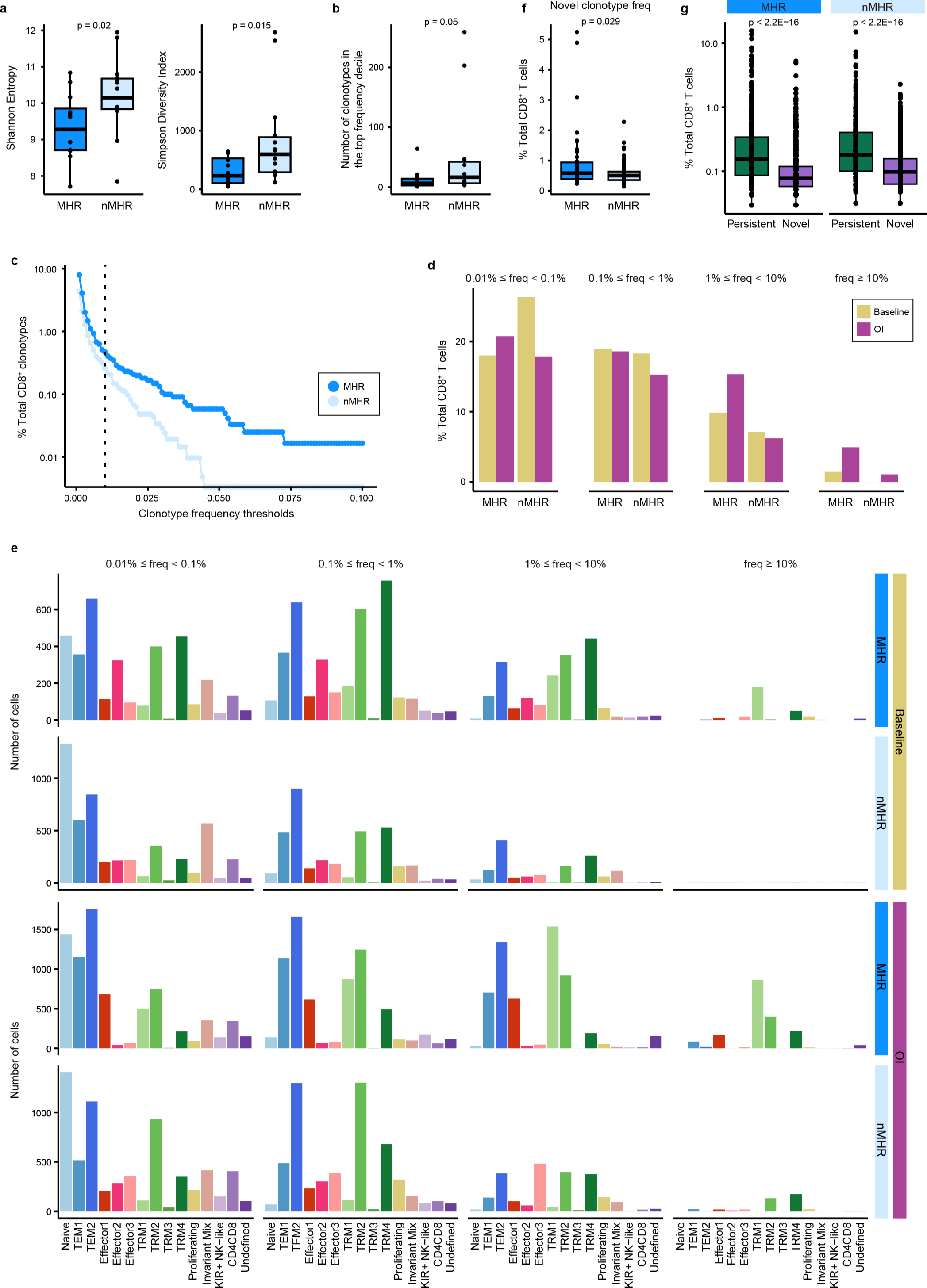
CD8^+^ T-cell TCR repertoire clonality and dynamics. Integration of scTCR-seq and scRNA-seq data for all samples enabled tracking of TCR clonotypes across timepoints and CD8 clusters. **a,** Shannon entropy (left) and Simpson diversity index (right) of baseline TCR repertoires from major histological responders (MHRs) and non-MHRs (nMHRs). Smaller values indicate greater clonality. **b,** Number of unique TCR clonotypes comprising the top frequency decile of baseline CD8^+^ T cells in MHRs and nMHRs. **c,** Fraction of baseline TCR clonotypes (y-axis) in MHRs and nMHRs with frequency greater than the indicated thresholds (x-axis). The fraction was calculated as the number of baseline CD8 clonotypes exceeding threshold frequency in a response group divided by total number of baseline CD8 clonotypes in the response group. **d,** Percent of total CD8^+^ T cells with indicated clonotype frequency at baseline and OI compared in MHRs and in nMHRs. **e,** Distribution among CD8 clusters of TCR clonotypes with indicated frequency in MHRs and nMHRs, at baseline and OI. y-axis, number of cells; x-axis, CD8 cluster. **f,** Frequency of novel clonotypes (i.e., detected only OI) in MHRs and nMHRs. **g,** Frequency of persistent (i.e., detected at baseline and OI) and novel TCR clonotypes compared in MHRs and nMHRs. **a,b,f,g,** Dots, individual values; midline, median; box limits, interquartile range; whiskers, 1.5x interquartile range. *P*-values were determined by Wilcoxon rank-sum test.

**Extended Data Fig. 8.**
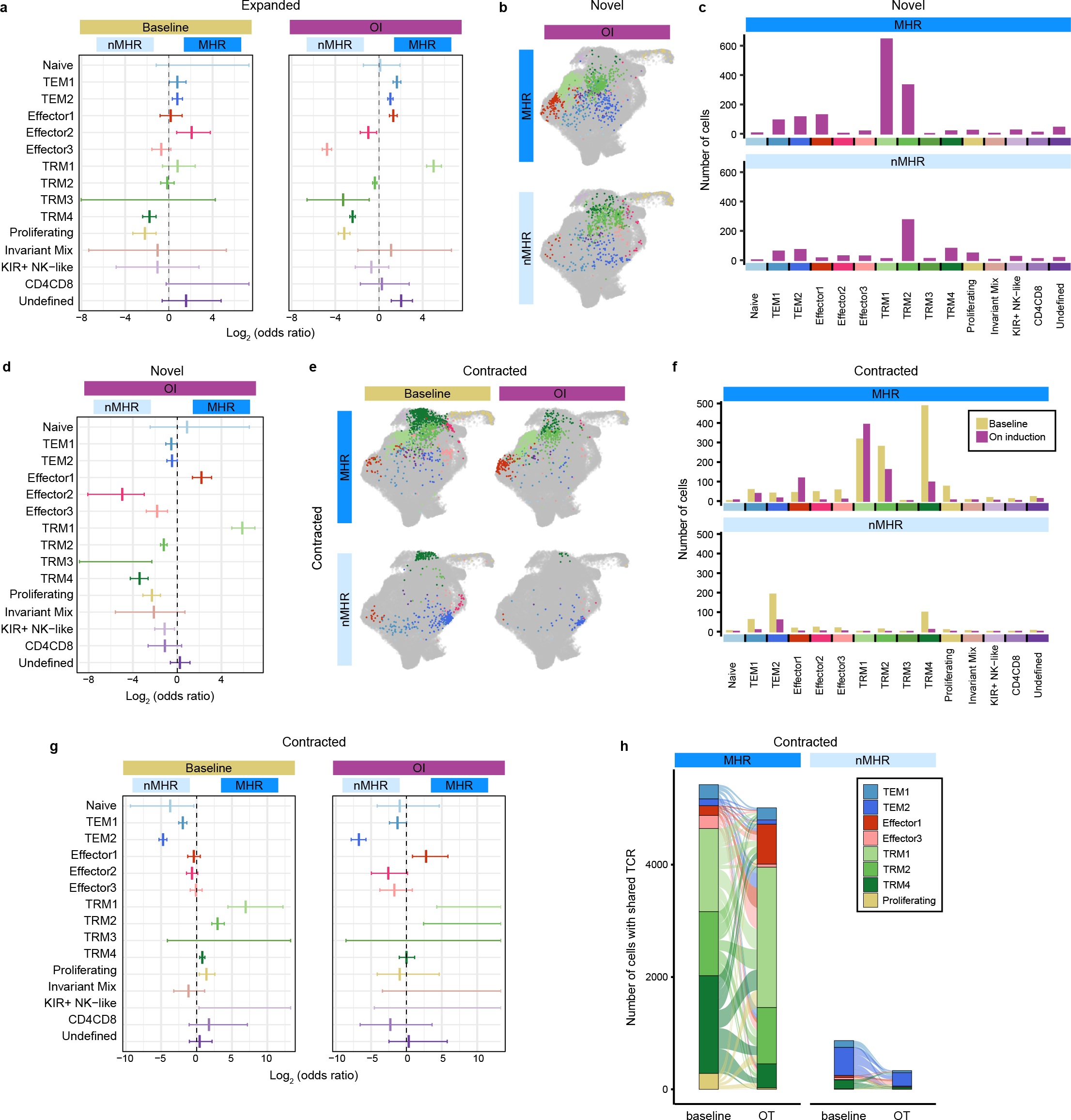
Phenotypic distribution of CD8^+^ T-cell clonotypes with altered frequency upon induction dual immune checkpoint blockade. Integration of scTCR-seq and scRNA-seq data for all samples enabled tracking of TCR clonotypes across timepoints and CD8 clusters. Clonotype frequencies at baseline and on induction (OI) CTLA-4 and PD-1 immune checkpoint blockade were used to identify TCR clonotypes that significantly expanded or contracted or were only detected (i.e., novel clones) in response to treatment. **a,** MHR:nMHR odds ratio of clonotype expansion in each cluster calculated at baseline (clonotypes that will expand) and OI (clonotypes that have expanded). **b,c,** Distribution of cells with novel TCR clonotypes among CD8 clusters at baseline and OI in MHRs and nMHRs, visualized as projections on the UMAP of total CD8^+^ T cells (**b**) and bar plots (**c**). **d,** MHR:nMHR odds ratio of novel clonotype detection in each CD8 cluster. **e,f,** Distribution of cells with contracted TCR clonotypes among CD8 clusters in MHRs and nMHRs, visualized as projections on the UMAP of total CD8^+^ T cells (**e**) and bar plots (**f**). **g,** MHR:nMHR odds ratio of clonotype contraction in each cluster calculated at baseline (clonotypes that will contract) and OI (clonotypes that have contracted). **h**, Alluvial plots showing the number (bar height) and phenotypic distribution (bar color) of baseline and OI cells that share a contracted TCR clonotype in MHRs or in nMHRs. Links, sharing between clusters; link color, baseline phenotype of shared clonotypes; link thickness, number of cells with shared clonotypes in each connected cluster. **a,d,g,** y-axis, CD8 cluster; x-axis, log_2_ odds ratio; midline, odds ratio (no midline, odds ratio not calculatable); horizontal line, 95% confidence interval.

**Extended Data Fig. 9.**
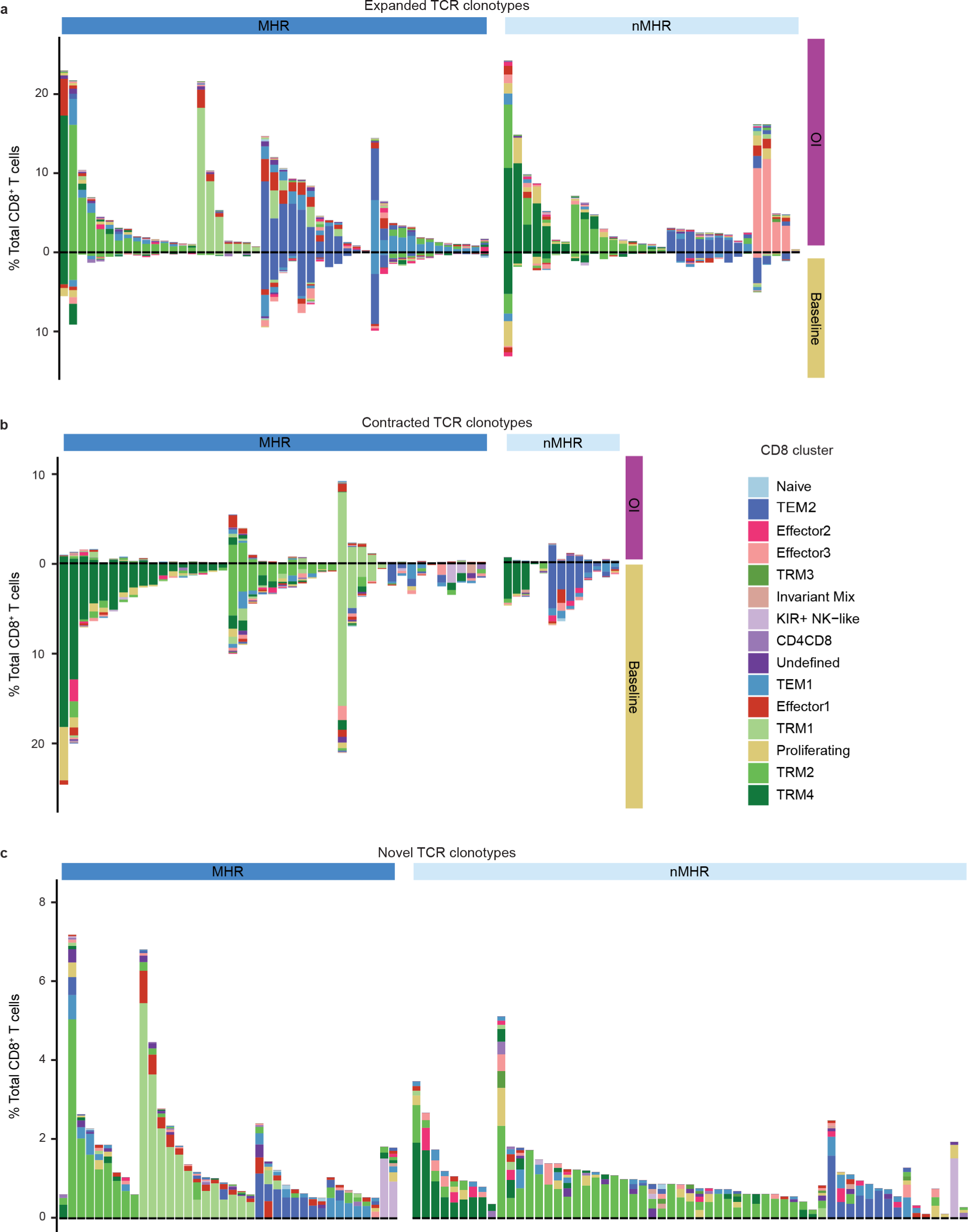
Predominant phenotypes of CD8^+^ T-cell clonotypes with altered frequency upon induction dual immune checkpoint blockade. The distribution among CD8 clusters of expanded, contracted, and novel TCR clonotypes from either major histological responders (MHRs) or non-MHRs was determined at baseline and on induction (OI) CTLA-4 and PD-1 immune checkpoint blockade. **a,** Distribution of expanded clonotypes (n=78), grouped by histological response and then by predominant OI phenotype. **b,** Distribution of contracted clonotypes (n=57), grouped by histological response and then by predominant baseline phenotype. **c,** Distribution of novel clonotypes (n=97), grouped by histological response and then by predominant phenotype. y-axis, clonotype frequency; x-axis, individual clonotypes; bar color, CD8 cluster. Clonotype frequency was calculated as the number of CD8^+^ T cells with a specific clonotype divided by the total number of CD8^+^ T cells in the same patient sample at each timepoint.

**Extended Data Fig. 10.**
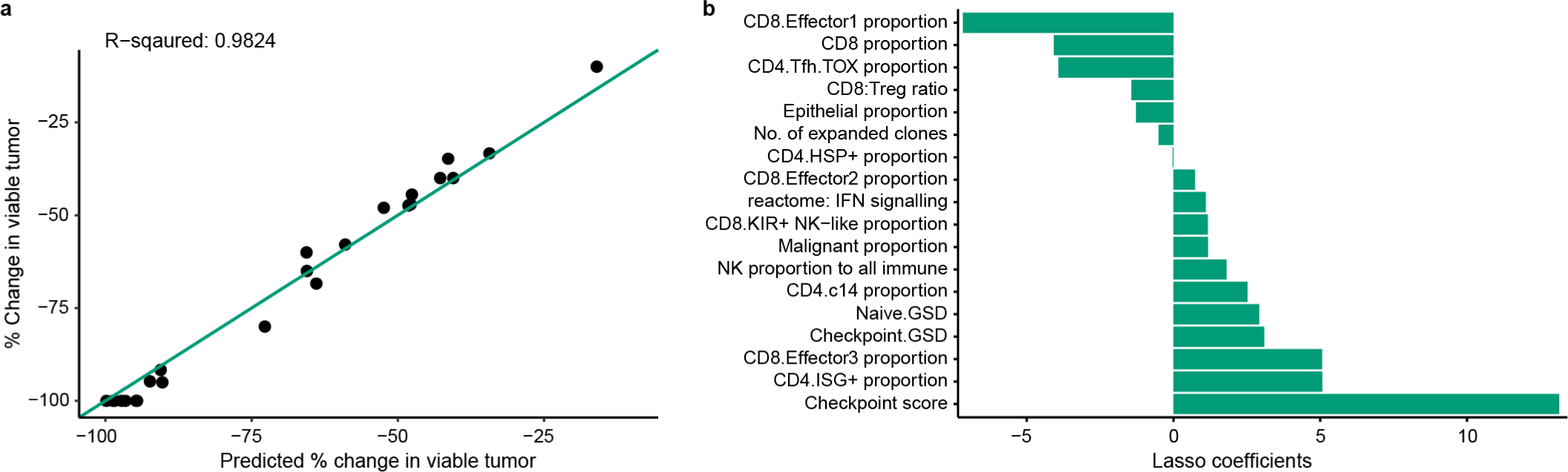
Delta LASSO regression model incorporates TIME features that change from baseline. A “delta” LASSO model incorporating both baseline and on-induction features of the tumor immune microenvironment (TIME) was developed to include treatment-induced changes that associated with percent reduction in tumor viability. **a,** Percent change in tumor viability per histological review (y-axis) vs percent change predicted by delta LASSO regression model (x-axis; n=26). Diagonal, line of identity (i.e., hypothetical scenario where the predicted values perfectly match the actual values). **b,** Features (y-axis; n=18) selected by the delta model and corresponding coefficients (x-axis).

