## Supplementary Material for "Depletion of effector regulatory T cells drives major response to induction dual immune checkpoint blockade"

X. Jiang et al.

**SUPPLEMENTARY MATERIAL**

Supplementary Figures 1–11 ..... 2
Supplementary Tables 1–5 ..... 21
Supplementary Tables 6–14 ..... separate file

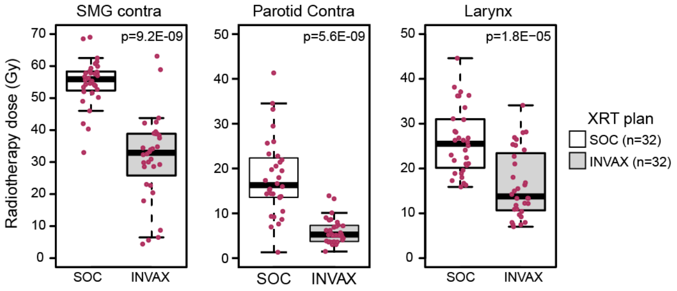

**Supplementary Fig. 1. Radiotherapy dose delivered to the contralateral submandibular gland and parotid and the larynx.** Patients on the INVAX trial received dose and volume response-adapted radiotherapy (XRT). INVAX XRT plans were evaluable for 32 patients. Boxplots show radiation dose (y-axis) delivered to the contralateral submandibular gland (SMG contra), contralateral parotid (parotid contra), and larynx according to standard-of-care (SOC) plan and according to trial protocol (INVAX) plan (x-axis). Dots, individual patient dose; midline, median; box, interquartile range; whiskers, 1.5x interquartile range. Wilcoxon rank-sum test used for comparison. *P*-values were false discovery rate adjusted. Mean doses delivered per SOC vs per INVAX were 36.9 vs 14.7 Gy to SMG contra, 18.1 vs 6.3 Gy to parotid contra, and 24.6 vs 13.7 Gy to larynx, respectively.

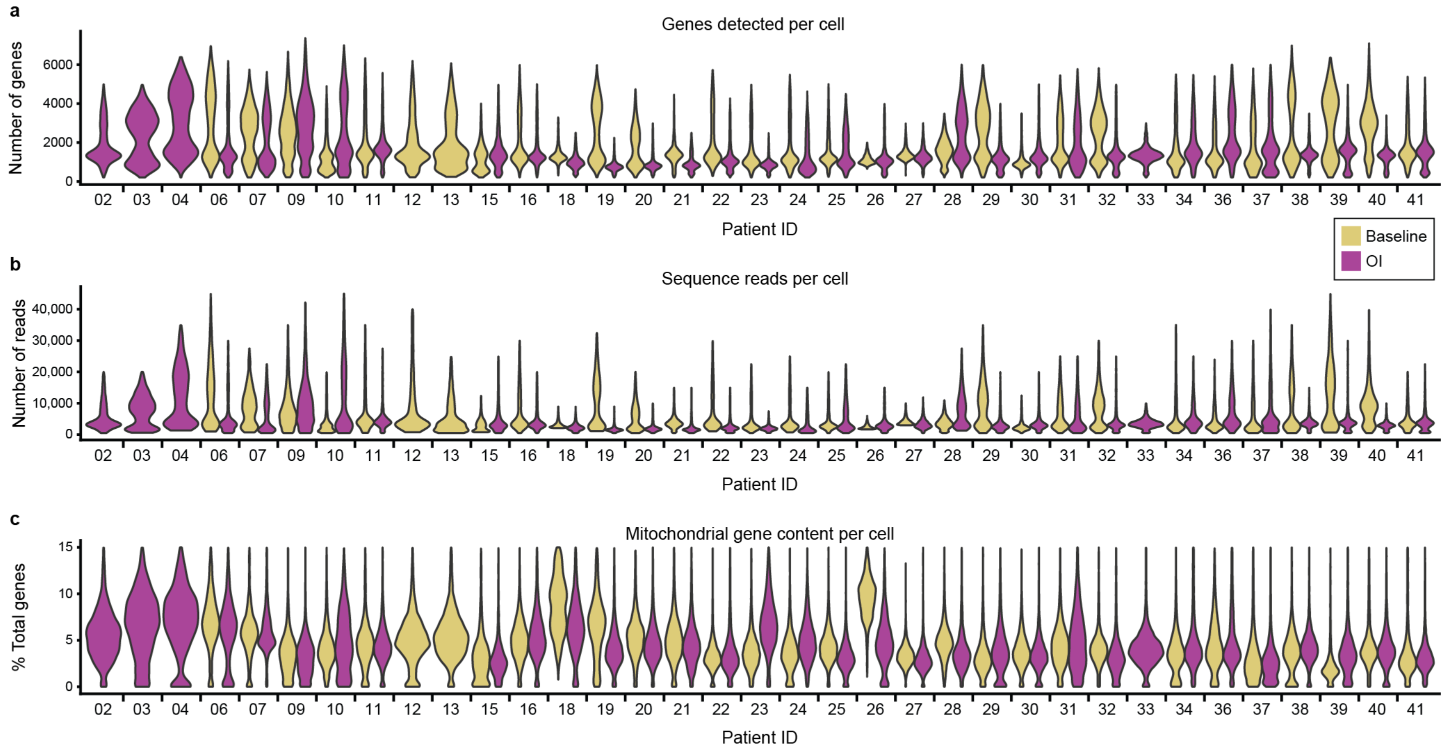

**Supplementary Fig. 2. Quality control metrics for scRNA-seq data.** Fresh tumor biopsies were collected at baseline and/or on induction (OI) CTLA-4 and PD-1 immune checkpoint blockade from 35 out of 37 enrolled HPV-positive oropharyngeal cancer patients. A total of 64 samples, including matched baseline-OI pairs from 29 patients, were analyzed by single-cell RNA sequencing (scRNA-seq) and single-cell T-cell receptor sequencing. **a-c**, Violin plots of the number of genes detected per cell (**a**), number of reads sequenced per cell, (**b**) or the percentage of mitochondrial genes among total genes detected per cell (**c**) in baseline and OI samples. x-axis, patient identifiers in order of enrollment (left to right).

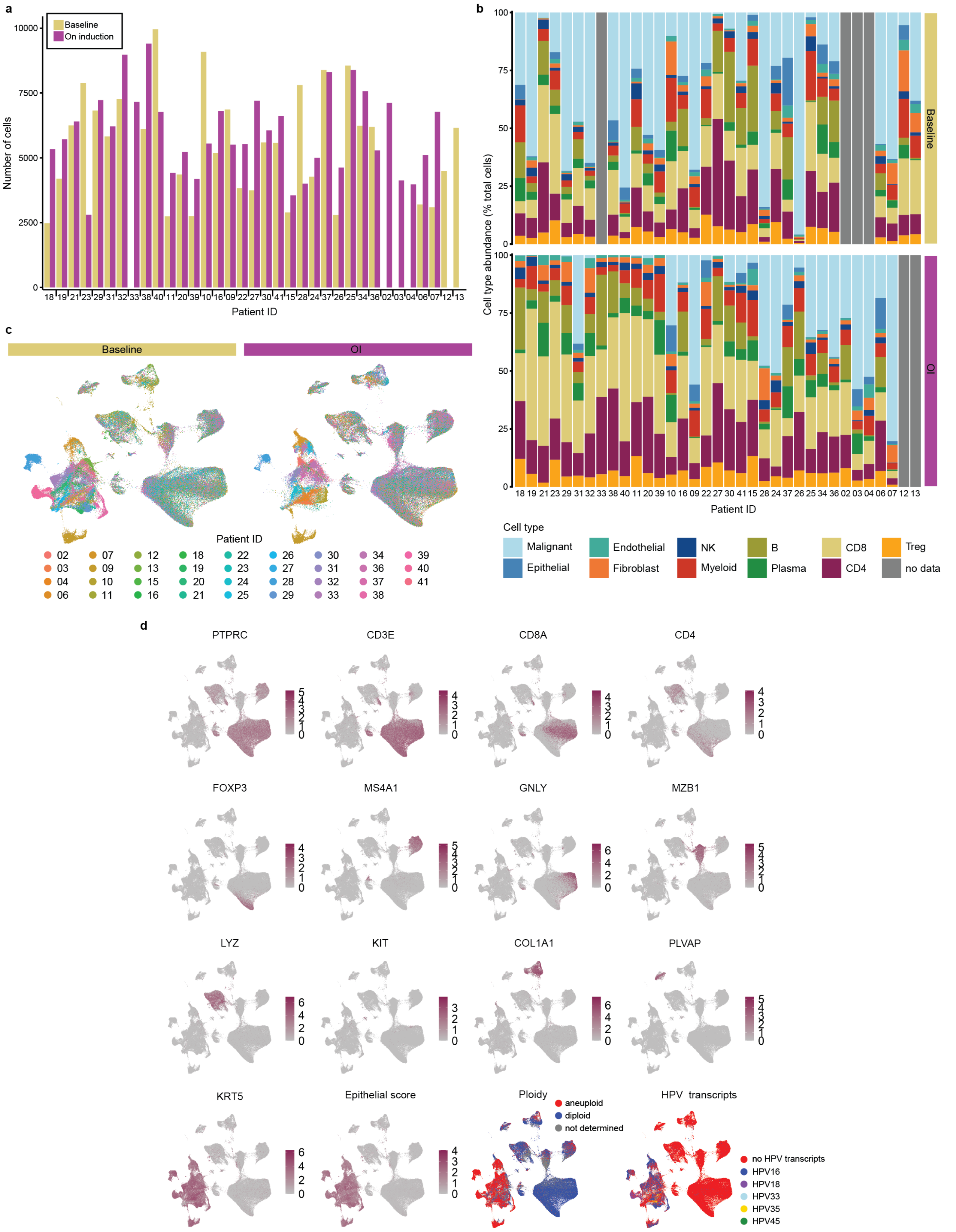
a
Baseline
On induction
Number of cells
10000
7500
5000
2500
0
18 19 21 23 29 31 32 33 38 40 11 20 39 10 16 09 22 27 30 41 15 28 24 37 26 25 34 36 02 03 04 06 07 12 13
Patient ID
b
Cell type abundance (% total cells)
100
75
50
25
0
Baseline
OI
18 19 21 23 29 31 32 33 38 40 11 20 39 10 16 09 22 27 30 41 15 28 24 37 26 25 34 36 02 03 04 06 07 12 13
Patient ID
Cell type
Malignant
Epithelial
Endothelial
Fibroblast
NK
Myeloid
B
Plasma
CD8
CD4
Treg
no data
c
Baseline
OI
Patient ID
02 03 04 06 07 09 10 11 12 13 15 16 18 19 20 21 22 23 24 25 26 27 28 29 30 31 32 33 34 36 37 38 39 40 41
d
PTPRC
CD3E
CD8A
CD4
FOXP3
MS4A1
GNLY
MZB1
LYZ
KIT
COL1A1
PLVAP
KRT5
Epithelial score
Ploidy
aneuploid
diploid
not determined
HPV transcripts
no HPV transcripts
HPV16
HPV18
HPV33
HPV35
HPV45

**Supplementary Fig. 3. Major cell types identified in the HPV-positive OPC tumor immune microenvironment at baseline and on induction dual immune checkpoint blockade.** Fresh tumor biopsies were collected at baseline and on induction (OI) dual CTLA-4 and PD-1 immune checkpoint blockade. **a,** Total number of tumor immune microenvironment (TIME) cells identified per sample at baseline and OI. **b,** Relative abundance of cell types in each sample. **a,b,** x-axis, patient identifiers arranged in descending order of the reduction in viable tumor (left to right) per histological review if sample was available or, if not, in order of enrollment (2, 3, 4, 6, 7, 12, and 13). **c,** Projection of patient identification on the UMAP of all TIME cells. **d,** Projections of cell-type marker expression on the UMAP. *PTPRC*: lymphocyte/myeloid cells, *CD3E*: T cells, *CD8A*: $CD8^+$ T cells, *CD4*: $CD4^+$ T cells, *FOXP3*: regulatory $CD4^+$ T cells, *MS4A1*: B cells, *GNLY*: NK cells, *MZB1*: plasma cells, *LYZ*: myeloid cells, *KIT*: monocytes, *COL1A1*: fibroblasts, *PLVAP*: endothelial cells, *KRT5*: epithelial cells. Ploidy and expression of HPV transcripts were used to identify malignant epithelial cells. HPV transcripts were also used to determine the HPV type (color code).

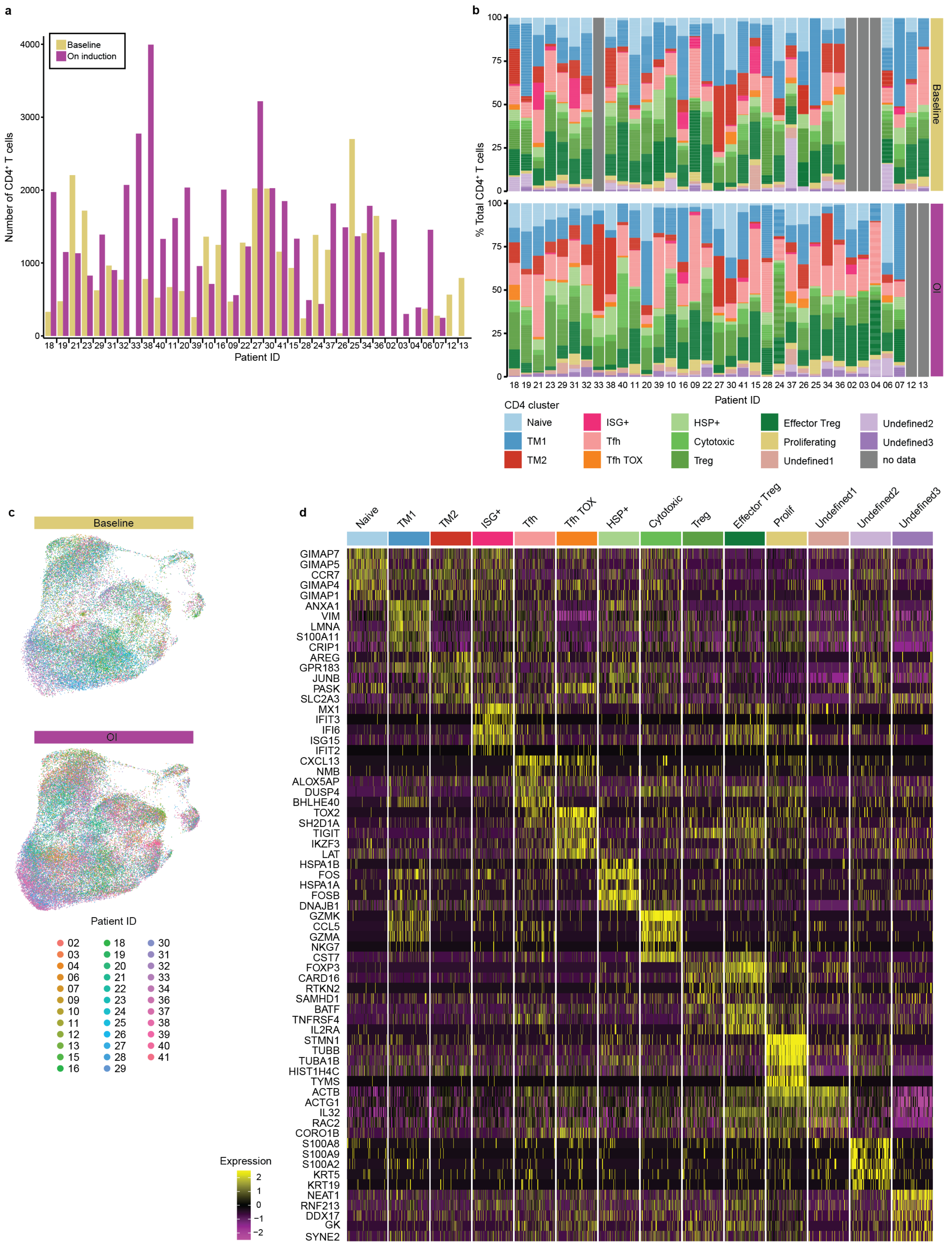
a
Baseline
On induction
Number of CD4+ T cells
4000
3000
2000
1000
0
18 19 21 23 29 31 32 33 38 40 11 20 39 10 16 09 22 27 30 41 15 28 24 37 26 25 34 36 02 03 04 06 07 12 13
Patient ID
b
% Total CD4+ T cells
100
75
50
25
0
Baseline
OI
18 19 21 23 29 31 32 33 38 40 11 20 39 10 16 09 22 27 30 41 15 28 24 37 26 25 34 36 02 03 04 06 07 12 13
Patient ID
CD4 cluster
Naive
TM1
TM2
ISG+
Tfh
Tfh TOX
HSP+
Cytotoxic
Treg
Effector Treg
Proliferating
Undefined1
Undefined2
Undefined3
no data
c
Baseline
OI
Patient ID
02 03 04 06 07 09 10 11 12 13 15 16
18 19 20 21 22 23 24 25 26 27 28 29
30 31 32 33 34 36 37 38 39 40 41
d
Naive
TM1
TM2
ISG+
Tfh
Tfh TOX
HSP+
Cytotoxic
Treg
Effector Treg
Prolif
Undefined1
Undefined2
Undefined3
GIMAP7
GIMAP5
CCR7
GIMAP4
GIMAP1
ANXA1
VIM
LMNA
S100A11
CRIP1
AREG
GPR183
JUNB
PASK
SLC2A3
MX1
IFIT3
IFI6
ISG15
IFIT2
CXCL13
NMB
ALOX5AP
DUSP4
BHLHE40
TOX2
SH2D1A
TIGIT
IKZF3
LAT
HSPA1B
FOS
HSPA1A
FOSB
DNAJB1
GZMK
CCL5
GZMA
NKG7
CST7
FOXP3
CARD16
RTKN2
SAMHD1
BATF
TNFRSF4
IL2RA
STMN1
TUBB
TUBA1B
HIST1H4C
TYMS
ACTB
ACTG1
IL32
RAC2
CORO1B
S100A8
S100A9
S100A2
KRT5
KRT19
NEAT1
RNF213
DDX17
GK
SYNE2
Expression
2
1
0
−1
−2

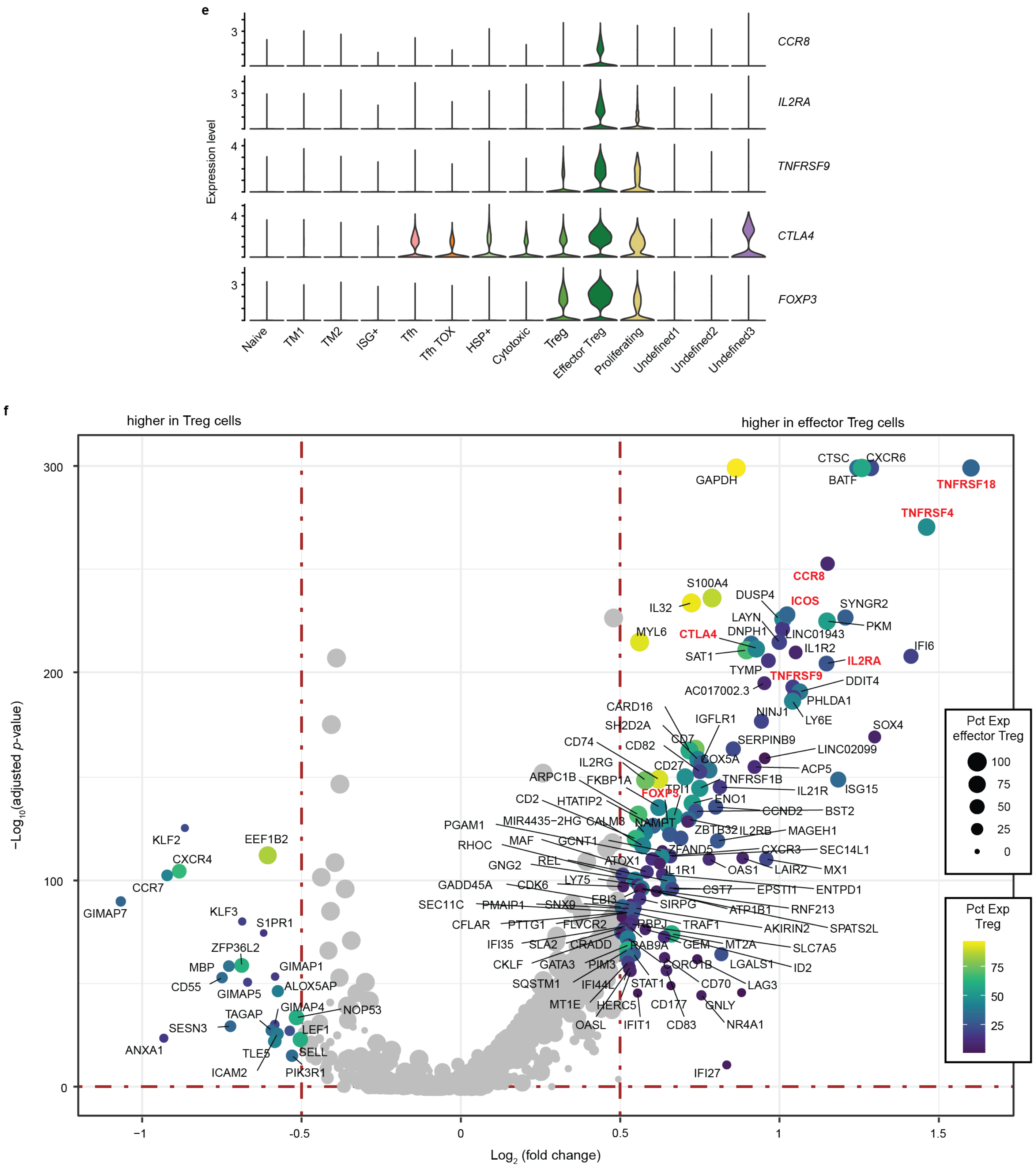

**Supplementary Fig. 4. CD4⁺ T cells identified in the HPV-positive OPC tumor immune microenvironment at baseline and on induction dual immune checkpoint blockade.** Fresh tumor biopsies were collected at baseline and on induction (OI) CTLA-4 and PD-1 immune checkpoint blockade. **a,** Total number of CD4⁺ T cells identified per sample at baseline and OI. **b,** Relative abundance of CD4⁺ T-cell clusters in each sample. **a,b,** x-axis, patient identifiers arranged in descending order of the reduction in viable tumor (left to right) per histological review if sample was available or, if not, in order of enrollment (2, 3, 4, 6, 7, 12, and 13). **c,** Projection of patient identification on the UMAP of all $CD4^+$ T cells. **d,** Heatmap of top 5 differentially expressed genes of each cluster. **e,** Violin plots of cluster-level expression of effector regulatory T (effector Treg) cell markers. **f,** Volcano plot of genes differentially expressed between effector Treg and Treg cells. y-axis, $-\log_{10}$ Bonferroni-adjusted *p*-value from Wilcoxon rank-sum test; x-axis, $\log_2$ fold change in expression; color scale, percent of cells in Treg cluster expressing gene; dot size, percent of cells in effector Treg cluster expressing gene; bold red text, genes of particular interest.

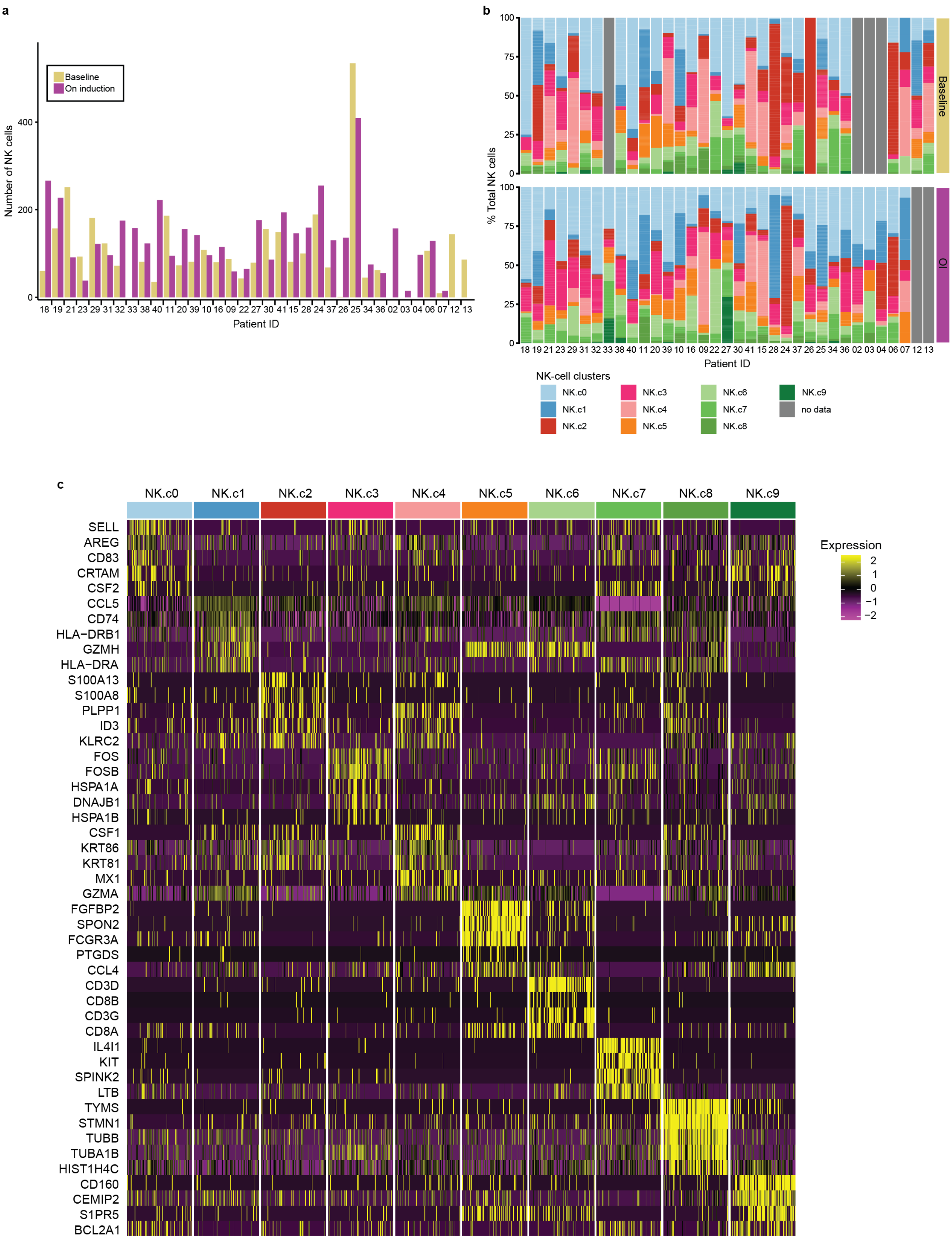
a
Baseline
On induction
Number of NK cells
400
200
0
18 19 21 23 29 31 32 33 38 40 11 20 39 10 16 09 22 27 30 41 15 28 24 37 26 25 34 36 02 03 04 06 07 12 13
Patient ID
b
100
75
50
25
0
% Total NK cells
Baseline
OI
18 19 21 23 29 31 32 33 38 40 11 20 39 10 16 09 22 27 30 41 15 28 24 37 26 25 34 36 02 03 04 06 07 12 13
Patient ID
NK-cell clusters
NK.c0
NK.c1
NK.c2
NK.c3
NK.c4
NK.c5
NK.c6
NK.c7
NK.c8
NK.c9
no data
c
NK.c0
NK.c1
NK.c2
NK.c3
NK.c4
NK.c5
NK.c6
NK.c7
NK.c8
NK.c9
SELL
AREG
CD83
CRTAM
CSF2
CCL5
CD74
HLA−DRB1
GZMH
HLA−DRA
S100A13
S100A8
PLPP1
ID3
KLRC2
FOS
FOSB
HSPA1A
DNAJB1
HSPA1B
CSF1
KRT86
KRT81
MX1
GZMA
FGFBP2
SPON2
FCGR3A
PTGDS
CCL4
CD3D
CD8B
CD3G
CD8A
IL4I1
KIT
SPINK2
LTB
TYMS
STMN1
TUBB
TUBA1B
HIST1H4C
CD160
CEMIP2
S1PR5
BCL2A1
Expression
2
1
0
−1
−2

**Supplementary Fig. 5. NK cells identified in the HPV-positive OPC tumor immune microenvironment at baseline and on induction dual immune checkpoint blockade.** Fresh tumor biopsies were collected at baseline and on induction (OI) dual CTLA-4 and PD-1 immune checkpoint blockade. **a,** Total number of NK cells identified per sample at baseline and OI. **b,** Relative abundance of NK-cell clusters in each sample. **a,b,** x-axis, patient identifiers arranged in descending order of the reduction in viable tumor (left to right) per histological review if sample was available or, if not, in order of enrollment (2, 3, 4, 6, 7, 12, and 13). **c,** Heatmap of top 5 differentially expressed genes of each cluster.

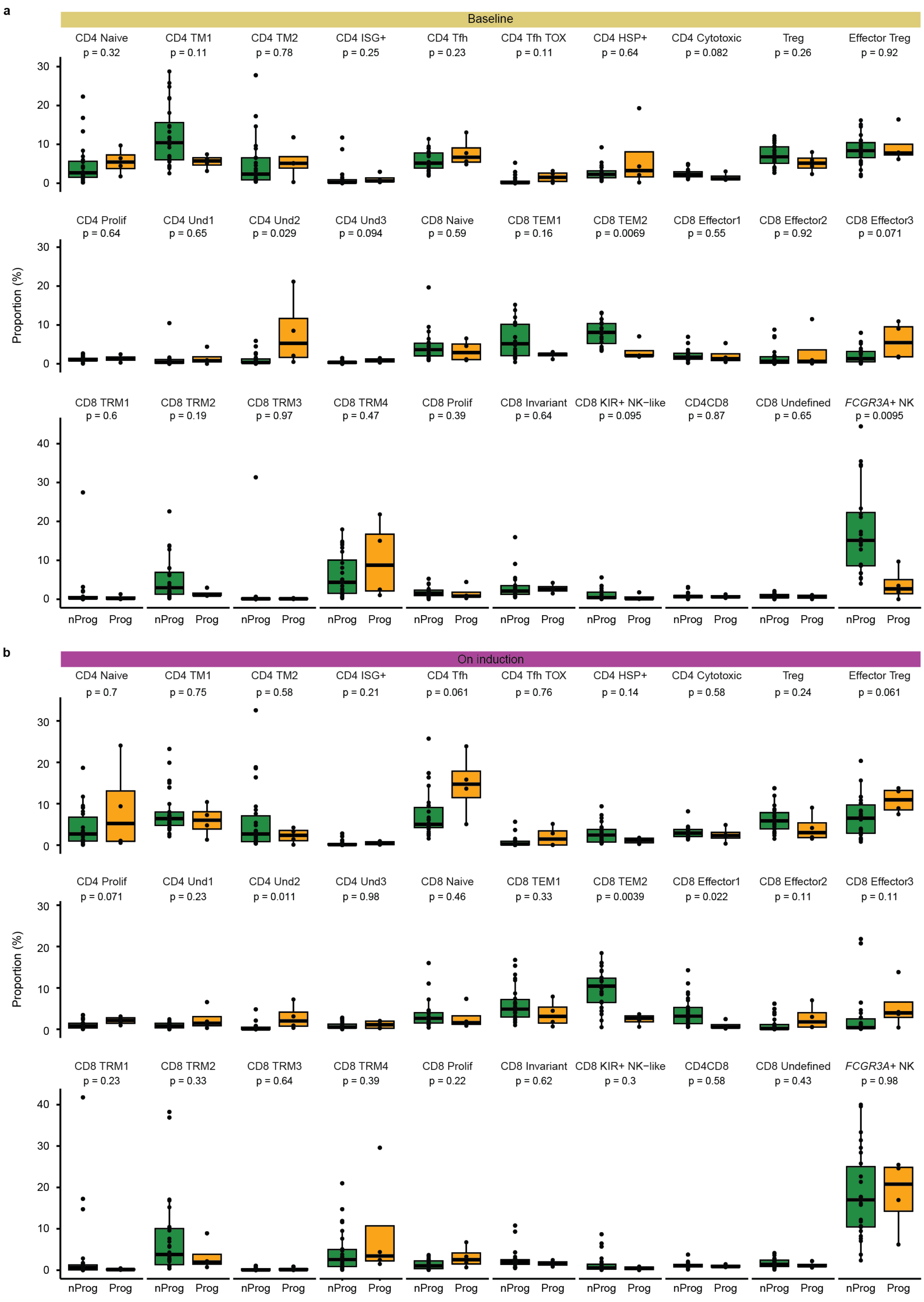
a
Baseline
CD4 Naive p = 0.32
CD4 TM1 p = 0.11
CD4 TM2 p = 0.78
CD4 ISG+ p = 0.25
CD4 Tfh p = 0.23
CD4 Tfh TOX p = 0.11
CD4 HSP+ p = 0.64
CD4 Cytotoxic p = 0.082
Treg p = 0.26
Effector Treg p = 0.92
CD4 Prolif p = 0.64
CD4 Und1 p = 0.65
CD4 Und2 p = 0.029
CD4 Und3 p = 0.094
CD8 Naive p = 0.59
CD8 TEM1 p = 0.16
CD8 TEM2 p = 0.0069
CD8 Effector1 p = 0.55
CD8 Effector2 p = 0.92
CD8 Effector3 p = 0.071
CD8 TRM1 p = 0.6
CD8 TRM2 p = 0.19
CD8 TRM3 p = 0.97
CD8 TRM4 p = 0.47
CD8 Prolif p = 0.39
CD8 Invariant p = 0.64
CD8 KIR+ NK−like p = 0.095
CD4CD8 p = 0.87
CD8 Undefined p = 0.65
*FCGR3A*+ NK p = 0.0095
Proportion (%)
0
10
20
30
40
nProg
Prog
b
On induction
CD4 Naive p = 0.7
CD4 TM1 p = 0.75
CD4 TM2 p = 0.58
CD4 ISG+ p = 0.21
CD4 Tfh p = 0.061
CD4 Tfh TOX p = 0.76
CD4 HSP+ p = 0.14
CD4 Cytotoxic p = 0.58
Treg p = 0.24
Effector Treg p = 0.061
CD4 Prolif p = 0.071
CD4 Und1 p = 0.23
CD4 Und2 p = 0.011
CD4 Und3 p = 0.98
CD8 Naive p = 0.46
CD8 TEM1 p = 0.33
CD8 TEM2 p = 0.0039
CD8 Effector1 p = 0.022
CD8 Effector2 p = 0.11
CD8 Effector3 p = 0.11
CD8 TRM1 p = 0.23
CD8 TRM2 p = 0.33
CD8 TRM3 p = 0.64
CD8 TRM4 p = 0.39
CD8 Prolif p = 0.22
CD8 Invariant p = 0.62
CD8 KIR+ NK−like p = 0.3
CD4CD8 p = 0.58
CD8 Undefined p = 0.43
*FCGR3A*+ NK p = 0.98
Proportion (%)
0
10
20
30
40
nProg
Prog

**Supplementary Fig. 6. Lymphocyte proportions in the tumor immune microenvironment of patients with and without disease progression.** Of 37 enrolled INVAX trial patients, 32 were evaluable for progression and had scRNA-seq data from either matched baseline and on induction (OI) samples (n=26), only baseline (n=2), or only OI (n=4). Of the 32 patients, 4 progressed, all of which had matched scRNA-seq data. **a,** Baseline proportions of $CD4^+$ T-cell clusters, $CD8^+$ T-cell clusters, and *FCGR3A*$^+$ NK cells in nonprogressors (nProg; n=24) and progressors (Prog; n=4). **b,** OI proportions of $CD4^+$ T-cell clusters, $CD8^+$ T-cell clusters, and *FCGR3A*$^+$ NK cells in nProgs (n=26) and Progs (n=4). Dots, individual patient values; midline, median; box, interquartile range; whiskers, 1.5x interquartile range. *P*-values were determined by Wilcoxon rank-sum test.

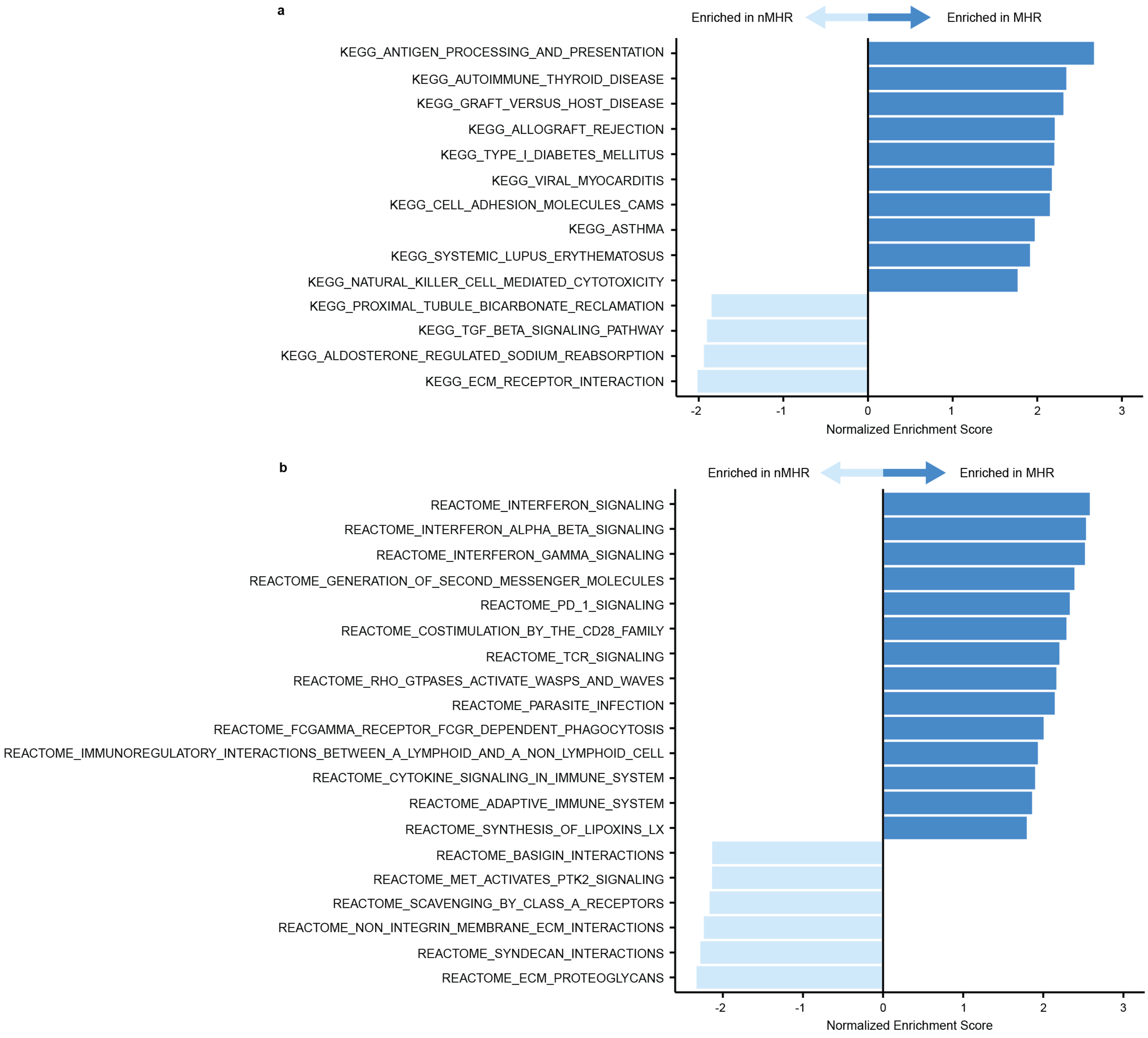
a
Enriched in nMHR
Enriched in MHR
KEGG_ANTIGEN_PROCESSING_AND_PRESENTATION
KEGG_AUTOIMMUNE_THYROID_DISEASE
KEGG_GRAFT_VERSUS_HOST_DISEASE
KEGG_ALLOGRAFT_REJECTION
KEGG_TYPE_I_DIABETES_MELLITUS
KEGG_VIRAL_MYOCARDITIS
KEGG_CELL_ADHESION_MOLECULES_CAMS
KEGG_ASTHMA
KEGG_SYSTEMIC_LUPUS_ERYTHEMATOSUS
KEGG_NATURAL_KILLER_CELL_MEDIATED_CYTOTOXICITY
KEGG_PROXIMAL_TUBULE_BICARBONATE_RECLAMATION
KEGG_TGF_BETA_SIGNALING_PATHWAY
KEGG_ALDOSTERONE_REGULATED_SODIUM_REABSORPTION
KEGG_ECM_RECEPTOR_INTERACTION
-2
-1
0
1
2
3
Normalized Enrichment Score
b
Enriched in nMHR
Enriched in MHR
REACTOME_INTERFERON_SIGNALING
REACTOME_INTERFERON_ALPHA_BETA_SIGNALING
REACTOME_INTERFERON_GAMMA_SIGNALING
REACTOME_GENERATION_OF_SECOND_MESSENGER_MOLECULES
REACTOME_PD_1_SIGNALING
REACTOME_COSTIMULATION_BY_THE_CD28_FAMILY
REACTOME_TCR_SIGNALING
REACTOME_RHO_GTPASES_ACTIVATE_WASPS_AND_WAVES
REACTOME_PARASITE_INFECTION
REACTOME_FCGAMMA_RECEPTOR_FCGR_DEPENDENT_PHAGOCYTOSIS
REACTOME_IMMUNOREGULATORY_INTERACTIONS_BETWEEN_A_LYMPHOID_AND_A_NON_LYMPHOID_CELL
REACTOME_CYTOKINE_SIGNALING_IN_IMMUNE_SYSTEM
REACTOME_ADAPTIVE_IMMUNE_SYSTEM
REACTOME_SYNTHESIS_OF_LIPOXINS_LX
REACTOME_BASIGIN_INTERACTIONS
REACTOME_MET_ACTIVATES_PTK2_SIGNALING
REACTOME_SCAVENGING_BY_CLASS_A_RECEPTORS
REACTOME_NON_INTEGRIN_MEMBRANE_ECM_INTERACTIONS
REACTOME_SYNDECAN_INTERACTIONS
REACTOME_ECM_PROTEOGLYCANS
-2
-1
0
1
2
3
Normalized Enrichment Score

**Supplementary Fig. 7. Gene set enrichment analysis comparing total MHR and nMHR CD8$^+$ T cells at baseline.** Fresh tumor biopsies were collected from newly diagnosed HPV-positive OPC patients at baseline, and CD8$^+$ T cells were identified by scRNA-seq. **a,b,** Barplots of KEGG (**a**) or reactome (**b**) pathways differentially enriched between total baseline CD8$^+$ T cells from major histological responders (MHRs) and non-MHRs (nMHRs). The normalized enrichment score was calculated using gene set enrichment analysis by R package (fgsea), and *p*-values were false discovery rate adjusted. Pathways with adjusted *p*-values <0.05 are shown.

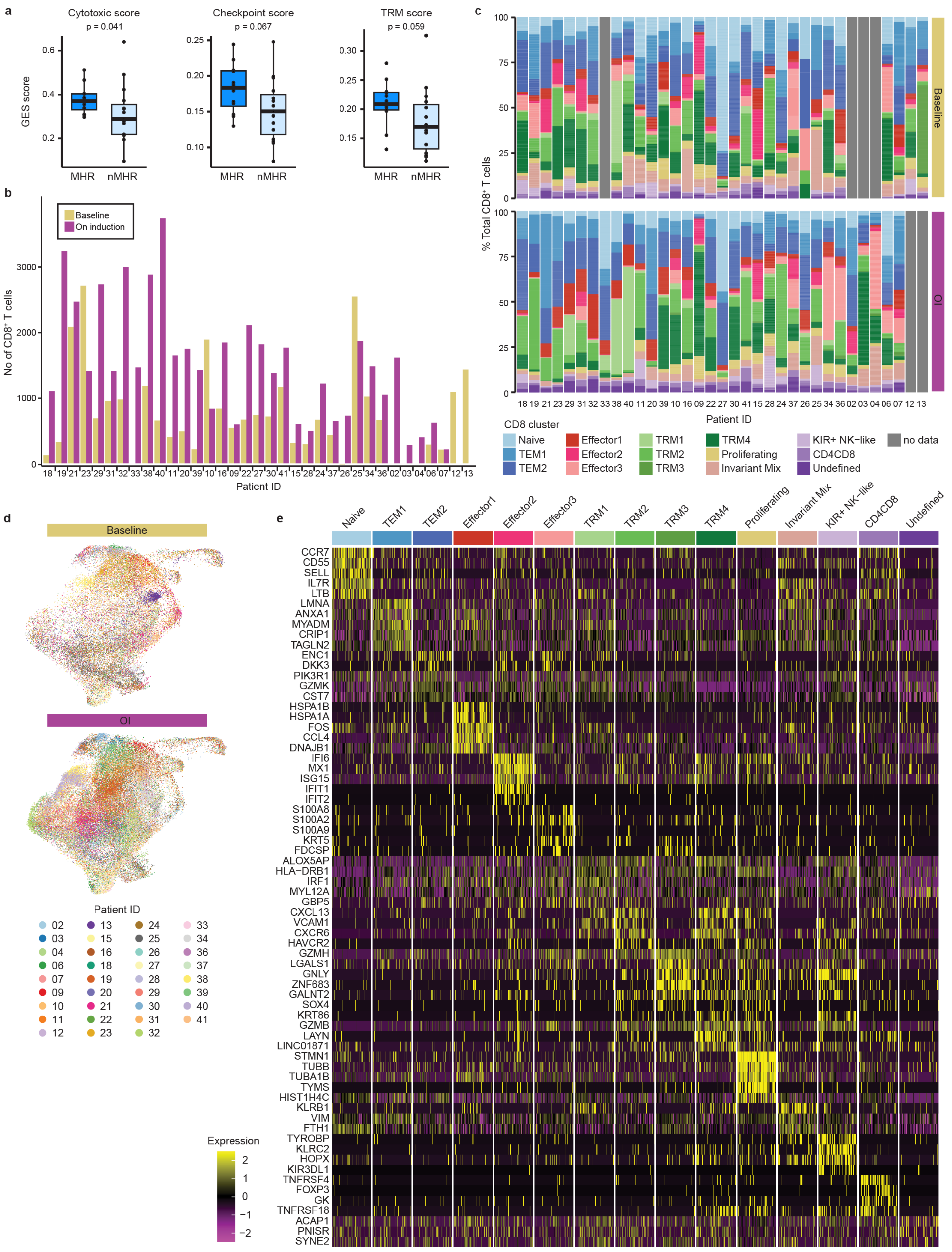

a
Cytotoxic score
p = 0.041
Checkpoint score
p = 0.067
TRM score
p = 0.059
GES score
MHR
nMHR
b
Baseline
On induction
No of CD8+ T cells
Patient ID
c
% Total CD8+ T cells
Baseline
OI
Patient ID
CD8 cluster
Naive
TEM1
TEM2
Effector1
Effector2
Effector3
TRM1
TRM2
TRM3
TRM4
Proliferating
Invariant Mix
KIR+ NK−like
CD4CD8
Undefined
no data
d
Baseline
OI
Patient ID
e
Naive
TEM1
TEM2
Effector1
Effector2
Effector3
TRM1
TRM2
TRM3
TRM4
Proliferating
Invariant Mix
KIR+ NK−like
CD4CD8
Undefined
CCR7
CD55
SELL
IL7R
LTB
LMNA
ANXA1
MYADM
CRIP1
TAGLN2
ENC1
DKK3
PIK3R1
GZMK
CST7
HSPA1B
HSPA1A
FOS
CCL4
DNAJB1
IFI6
MX1
ISG15
IFIT1
IFIT2
S100A8
S100A2
S100A9
KRT5
FDCSP
ALOX5AP
HLA−DRB1
IRF1
MYL12A
GBP5
CXCL13
VCAM1
CXCR6
HAVCR2
GZMH
LGALS1
GNLY
ZNF683
GALNT2
SOX4
KRT86
GZMB
LAYN
LINC01871
STMN1
TUBB
TUBA1B
TYMS
HIST1H4C
KLRB1
VIM
FTH1
TYROBP
KLRC2
HOPX
KIR3DL1
TNFRSF4
FOXP3
GK
TNFRSF18
ACAP1
PNISR
SYNE2
Expression

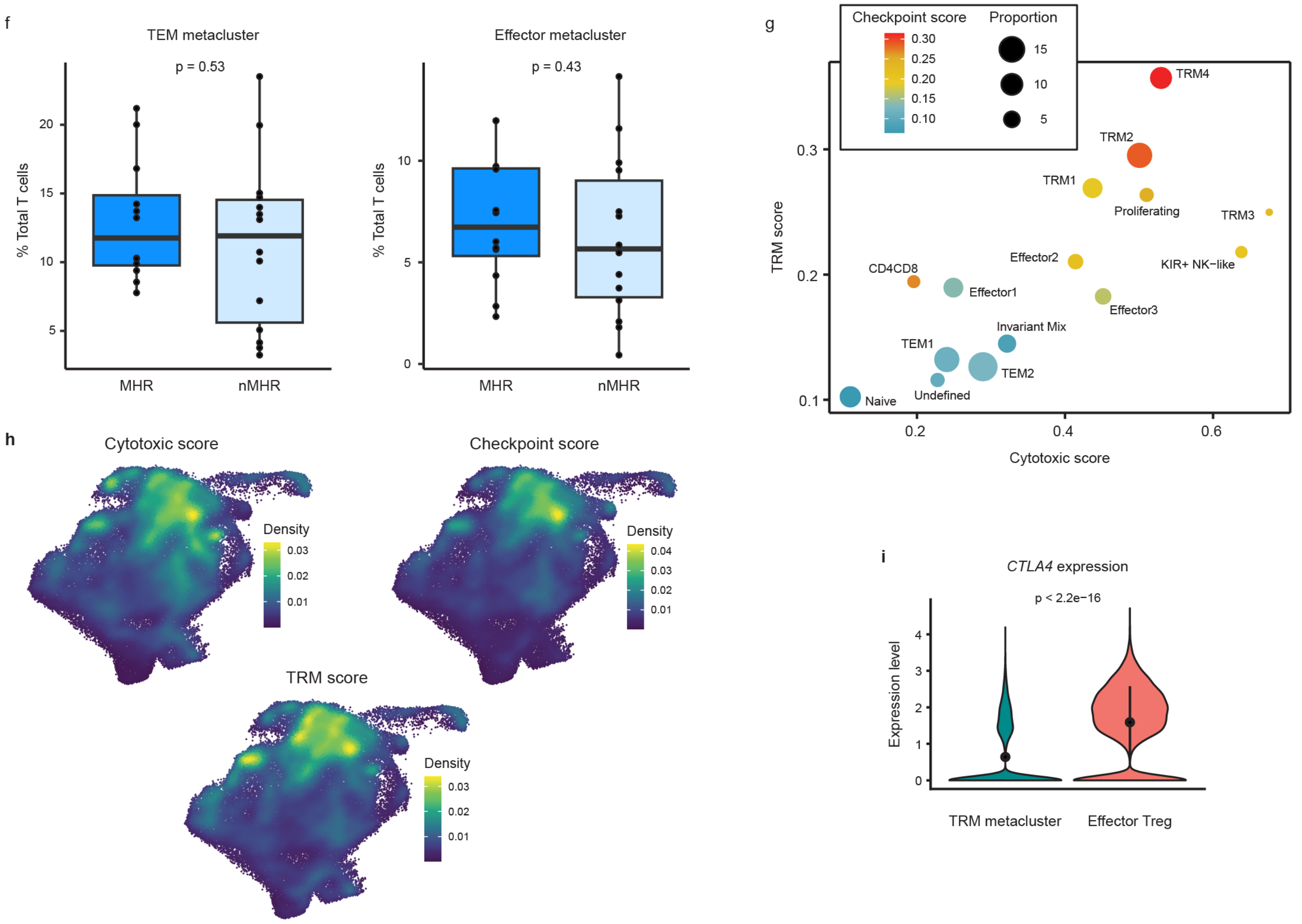

**Supplementary Fig. 8. CD8⁺ T cells identified in the HPV-positive OPC tumor immune microenvironment at baseline and after induction dual immune checkpoint blockade.** Fresh tumor biopsies were collected from newly diagnosed HPV-positive OPC patients at baseline and on induction (OI) CTLA-4 and PD-1 immune checkpoint blockade. **a,** Boxplots comparing cytotoxic (*PRF1, GZMA, GZMB, GZMH,* and *GNLY*), immune checkpoint (*PDCD1, CTLA4, TIM3* [*HAVCR2*], *LAG3*, and *TOX*) and tissue-resident memory (TRM; *ITGAE* [*CD103*], *ZNF683* [*HOBIT*], *ITGA1* [*CD49a*], *CD69, CXCR6*, and *CXCL13*) gene expression signature (GES) scores for CD8⁺ T cells from major histological responders (MHRs; n=12) and non-MHRs (nMHRs; n=15) at baseline. **b,** Total number of CD8⁺ T cells identified in each sample. **c,** Relative abundance of each CD8⁺ T-cell cluster in each sample. **b,c,** x-axis, patient identifiers arranged in descending order of the reduction in viable tumor (left to right) per histological review if sample was available or, if not, in order of enrollment (2, 3, 4, 6, 7, 12, and 13). **d,** Projection of patient information on the UMAP of all CD8⁺ T cells. **e,** Heatmap of top 5 differentially expressed genes of each cluster. **f,** Boxplots comparing proportion of total CD8⁺ T cells in the effector memory (TEM) and effector metaclusters in MHRs (n=12) vs nMHRs (n=14) at baseline. **g,** Scatter plot of the average TRM (y-axis) and cytotoxic (x-axis) GES scores for each cluster. Color scale, checkpoint GES score; circle size, cluster proportion (percent of total CD8⁺ T cells). **h,** Density plots of indicated GES scores projected on the total CD8⁺ T-cell UMAP. **a,f,** Dots, individual patient values; midline, median value; box, interquartile range; whiskers, 1.5x interquartile range. *P*-values were determined by Wilcoxon rank-sum test. **i,** Violin plot comparing *CTLA4* expression in the TRM metacluster and effector Treg cluster. Dot, mean value; line, +/- standard deviation from the mean. *P*-value was determined by Wilcoxon rank-sum test. **a, g, h,** GES scores calculated by AUCell.

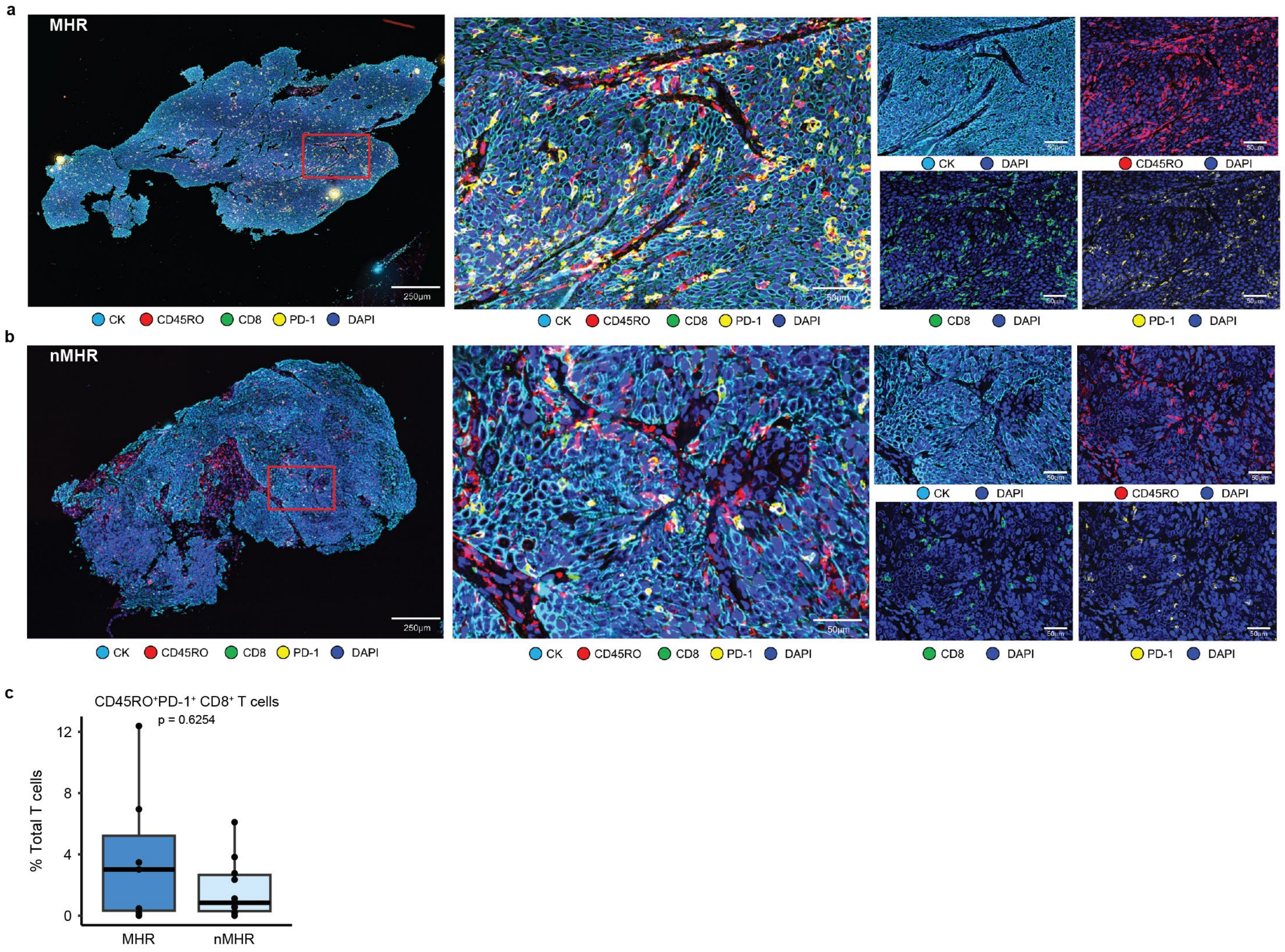

**Supplementary Fig. 9. Multiplexed fluorescent imaging of antigen-experienced, memory CD8$^+$ T cells in HPV-positive OPC tumors at baseline.** Multiplex barcoded fluorescent imaging was performed on baseline FFPE tumor tissue from HPV-positive OPC to assess tumor infiltration by CD45RO$^+$PD-1$^+$ CD8$^+$ T cells in major histological responders (MHRs) and non-MHRs (nMHRs). **a,b,** Representative images of a MHR (**a**) and a nMHR (**b**). *Left*, panoramic view (5× magnification; scale bar, 125 μm) of tissue sample, including cytokeratin (CK; *cyan*), CD45RO (*red*), CD8 (*green*), PD-1 (*yellow*), and 4′,6-diamidino-2-phenylindole (DAPI; *blue*) signals; *middle*, 20× magnification image (scale bar, 50 μm) of area outlined by red rectangle in the panoramic image; *right*, individual channels in combination with DAPI. **c,** Percent of CD45RO$^+$PD-1$^+$ CD8$^+$ T cells among total T cells (defined as CD45$^+$CD3$^+$ cells; images not shown) was quantified in MHRs (n=7) and nMHRs (n=10; see **Methods**). *P*-value determined by Wilcoxon rank-sum test.

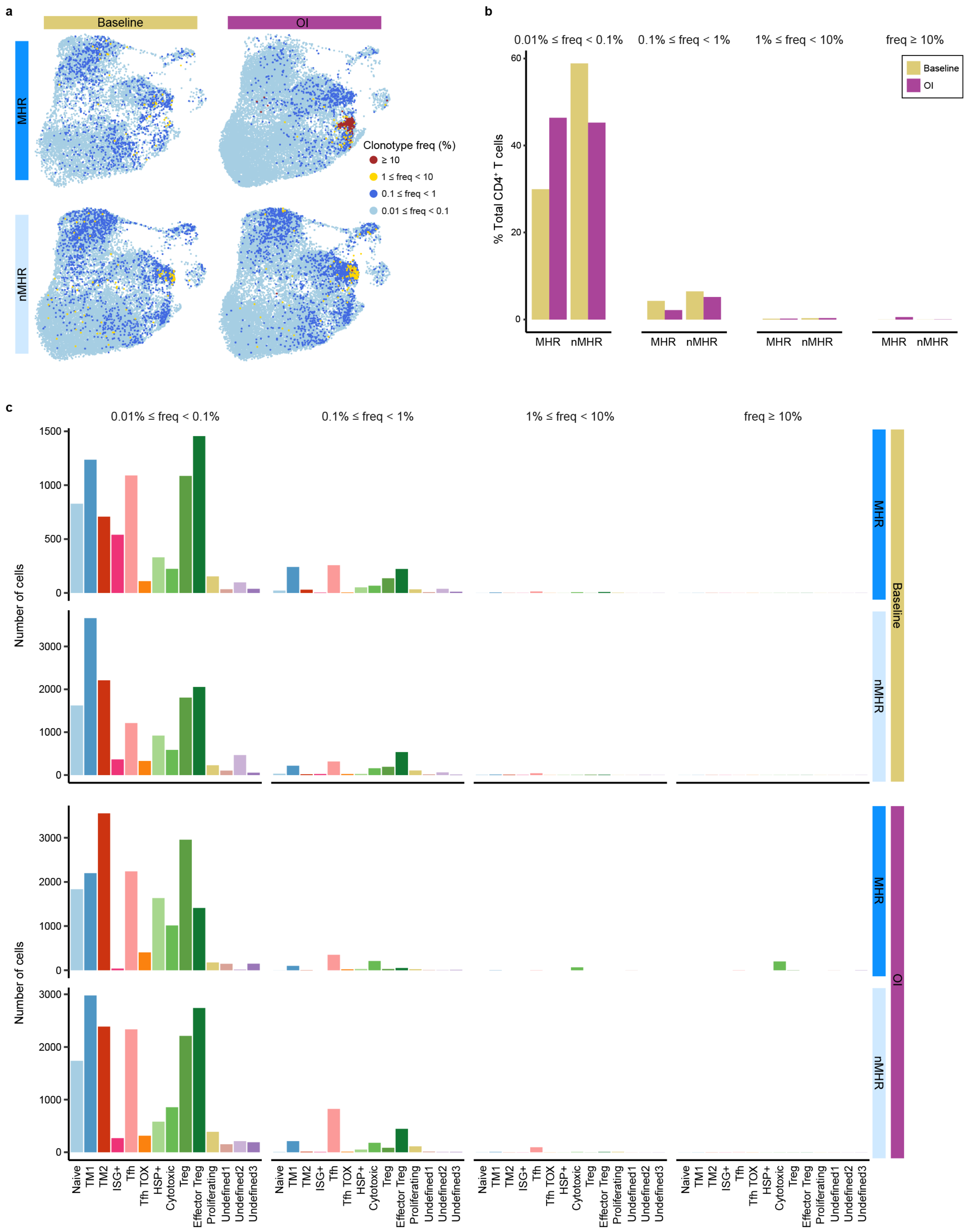

**Supplementary Fig. 10. Landscape of CD4$^+$ T-cell TCR clonotypes.** Integration of scTCR-seq and scRNA-seq data for all samples enabled tracking of TCR clonotypes across timepoints and CD4 clusters. **a,** Distribution of TCR clonotypes with indicated frequency at baseline and on induction (OI) CTLA-4 and PD-1 immune checkpoint blockade projected on the UMAP of total $CD4^+$ T cells. **b,** Percent of total $CD4^+$ T cells with indicated clonotype frequency at baseline and OI compared in MHRs and in nMHRs. **c,** Distribution among CD4 clusters of TCR clonotypes with indicated frequency in MHRs and nMHRs, at baseline and OI. y-axis, number of cells; x-axis, CD4 cluster.

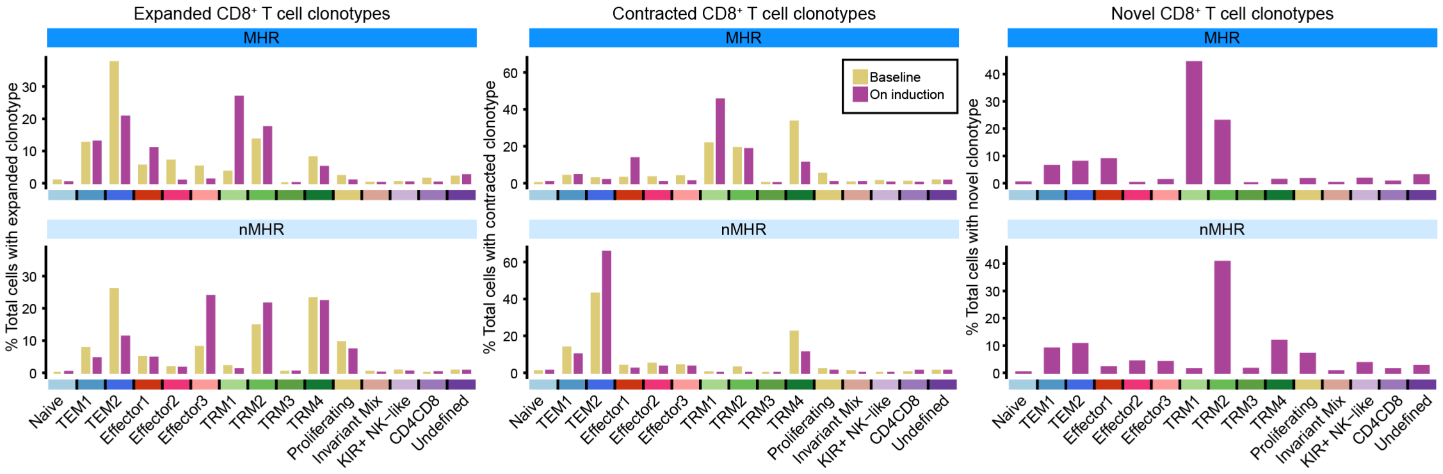

**Supplementary Fig. 11. Phenotypic distribution of CD8$^+$ T-cell TCR clonotypes responsive to induction dual immune checkpoint blockade.** Combined scTCR-seq and scRNA-seq data were used determine the distribution of expanded, contracted, and novel TCR clonotypes among CD8$^+$ T-cell clusters at baseline (clonotypes that will expand/contract) and on induction (OI; clonotypes that have expanded/contracted and novel clonotypes) in major histological responder (MHR) and non-MHR (nMHR) groups. y-axis, percent of total cells with expanded, contracted, or novel TCR clonotypes, respectively; x-axis, CD8$^+$ T-cell cluster.

**Supplementary Table 1. Patient and tumor characteristics.**

| | Total (n=35) |
|---|---|
| Age (years) | |
| ≤ 65 | 23 (66%) |
| > 65 | 12 (34%) |
| Mean (standard deviation) | 63.2 (6.9) |
| Median | 63.9 |
| Min - max | 48–78 |
| Gender | |
| Male | 34 (97%) |
| Female | 1 (3%) |
| Race | |
| White | 33 (94%) |
| Black | 1 (3%) |
| Other | 1 (3%) |
| Ethnicity | |
| Hispanic NOS | 4 (11%) |
| Other non-Hispanic | 31 (89%) |
| Smoking history | |
| Never | 15 (43%) |
| Former | 11 (31%) |
| Current | 9 (26%) |
| ≥ 10 pack-years | 7 (20%) |
| T stage (AJCC 8th Edition) | |
| T1 | 14 (37%) |
| T2 | 15 (43%) |
| T3 | 6 (20%) |
| N stage (AJCC 8th Edition) | |
| N0 | 0 (0%) |
| N1 | 33 (94%) |
| N2 | 2 (6%) |
| Overall stage (AJCC 8th Edition) | |
| I | 33 (94%) |
| II | 2 (6%) |
| Risk group per RTOG 0129 | |
| Low-risk | 30 (86%) |
| Intermediate-risk | 5 (14%) |
| PD-L1 CPS | |
| 1–19 | 19 (54%) |
| ≥ 20 | 16 (46%) |

**Supplementary Table 2. Clinical and histopathological characteristics of individual patients.**

| Patient ID | Age | Gender | Tobacco use | HPV type | N stage | T stage | Base tumor viability (%) | OI tumor viability (%) | Histo response | RECIST response | Progression at follow up | Base sc data | OI sc data |
|---|---|---|---|---|---|---|---|---|---|---|---|---|---|
| 1 | 61-65 | male | former | 16 | cN2 | cT2 | 95 | N.D. | N.A. | stable | Y | N | N |
| 2 | 61-65 | male | never | 16 | cN1 | cT1 | 100 | N.D. | N.A. | stable | N | N | Y |
| 3 | 51-55 | female | never | 35 | cN1 | cT2 | 95 | N.D. | N.A. | N/E | N/E | N | Y |
| 4 | 61-65 | male | current | 16 | cN1 | cT3 | 95 | N.D. | N.A. | stable | N | N | Y |
| 5 | 61-65 | male | never | 16 | cN1 | cT1 | 95 | N.D. | N.A. | stable | N | N | N |
| 6 | 56-60 | male | current | 16 | cN1 | cT2 | 95 | N.D. | N.A. | stable | Y | Y | Y |
| 7 | 71-75 | male | current | 16 | cN1 | cT1 | 100 | N.D. | N.A. | stable | N | Y | Y |
| 9 | 56-60 | male | former | 16 | cN1 | cT3 | 100 | 35 | nMHR | stable | Y | Y | Y |
| 10 | 66-70 | male | former | 16 | cN1 | cT1 | 100 | 20 | nMHR | stable | N | Y | Y |
| 11 | 66-70 | male | former | 16 | N1 | cT2 | 100 | 5 | MHR | PR | N | Y | Y |
| 12 | 76-80 | male | current | 16 | cN1 | cT1 | 95 | N.D. | N.A. | stable | N | Y | N |
| 13 | 76-80 | male | never | 16 | cN1 | cT2 | N.D. | N.D. | N.A. | stable | N | Y | N |
| 15 | 51-55 | female | never | 16 | cN1 | cT2 | 85 | 45 | nMHR | stable | N | Y | Y |
| 16 | 46-50 | male | current | 16 | cN1 | cT2 | 95 | 30 | nMHR | N/E | N/E | Y | Y |
| 18 | 46-50 | male | current | 16 | cN1 | cT1 | 98 | 0 | MHR | stable | N | Y | Y |
| 19 | 66-70 | male | never | 16 | cN1 | cT2 | 95 | 0 | MHR | PR | N | Y | Y |
| 20 | 66-70 | male | former | 33 | cN1 | cT1 | 95 | 5 | MHR | stable | N | Y | Y |
| 21 | 61-65 | male | never | 16 | cN1 | cT1 | 95 | 0 | MHR | PR | N.D. | Y | Y |
| 22 | 56-60 | male | former | 16 | cN1 | cT2 | 100 | 40 | nMHR | progression | N | Y | Y |
| 23 | 51-55 | male | never | 16 | cN1 | cT2 | 95 | 0 | MHR | PR | N | Y | Y |
| 24 | 61-65 | male | never | 33 | cN1 | cT1 | 100 | 60 | nMHR | stable | N | Y | Y |
| 25 | 66-70 | male | never | 16 | cN1 | cT2 | 92 | 60 | nMHR | stable | N | Y | Y |
| 26 | 66-70 | male | never | 16 | cN1 | cT1 | 95 | 60 | nMHR | stable | N | Y | Y |
| 27 | 56-60 | male | never | 45 | pN1 | pT1 | 95 | 40 | nMHR | stable | N | Y | Y |
| 28 | 56-60 | male | former | 16 | cN1 | cT1 | 90 | 50 | nMHR | stable | N | Y | Y |
| 29 | 51-55 | male | former | 16 | cN1 | cT1 | 95 | 0 | MHR | stable | N | Y | Y |
| 30 | 51-55 | male | former | 16 | cN1 | cT3 | 100 | 52 | nMHR | stable | N | Y | Y |

| | | | | | | | | | | | | | |
|---|---|---|---|---|---|---|---|---|---|---|---|---|---|
| 31 | 66-70 | male | current | 16 | cN1 | cT2 | 100 | 0 | MHR | stable | N | Y | Y |
| 32 | 61-65 | male | never | 16 | cN1 | cT2 | 95 | 0 | MHR | stable | N | Y | Y |
| 33 | 56-60 | male | never | 16 | cN0 | cT2 | 90 | 0 | MHR | stable | N | N | Y |
| 34 | 61-65 | male | current | 16 | cN1 | cT3 | 90 | 60 | nMHR | stable | N | Y | Y |
| 36 | 66-70 | male | current | 16 | cN1 | cT1 | 100 | 90 | nMHR | stable | Y | Y | Y |
| 37 | 56-60 | male | former | 16 | cN1 | cT2 | 100 | 60 | nMHR | stable | Y | Y | Y |
| 38 | 61-65 | male | current | 16 | cN1 | cT3 | 99 | 0 | MHR | PR | N | Y | Y |
| 39 | 61-65 | male | never | 16 | cN1 | cT3 | 60 | 5 | MHR | stable | N | Y | Y |
| 40 | 66-70 | male | never | 18 | cN1 | cT1 | 95 | 0 | MHR | stable | N | Y | Y |
| 41 | 61-65 | male | current | 33 | cN2 | cT2 | 95 | 50 | nMHR | stable | N | Y | Y |

Age, age at time of consent; base, baseline; OI, on induction dual immune checkpoint blockade; Histo, histological; MHR, major histological responder; nMHR, non-MHR; sc, single-cell; PR, partial response; N.D., no data; N.A., not applicable; N/E, nonevaluable.

**Supplementary Table 3. Treatment related adverse events that occurred after first dose.**

| AE | All grade | | Grade 1 | | Grade 2 | | Grade 3 | | Grade 4 | |
|---|---|---|---|---|---|---|---|---|---|---|
| | Count | % | Count | % | Count | % | Count | % | Count | % |
| Mucositis (tongue, throat, mouth) | 31 | 89 | 8 | 23 | 18 | 51 | 5 | 14 | | |
| Dysgeusia | 29 | 83 | 14 | 40 | 15 | 43 | | | | |
| Fatigue | 29 | 83 | 19 | 54 | 7 | 20 | 3 | 9 | | |
| Pain (tongue, mouth, throat) | 26 | 74 | 13 | 37 | 10 | 29 | 3 | 9 | | |
| Dry mouth | 24 | 69 | 17 | 49 | 7 | 20 | | | | |
| Dysphagia/Odynophagia | 24 | 69 | 7 | 20 | 11 | 31 | 6 | 17 | | |
| Dermatitis radiation | 22 | 63 | 7 | 20 | 15 | 43 | | | | |
| Weight loss | 19 | 54 | 8 | 23 | 7 | 20 | 4 | 11 | | |
| Serum amylase increased | 17 | 49 | 8 | 23 | 5 | 14 | 4 | 11 | | |
| Hypothyroidism | 16 | 46 | 5 | 14 | 11 | 31 | | | | |
| Nausea | 15 | 43 | 13 | 37 | 2 | 6 | | | | |
| Alanine aminotransferase increased | 13 | 37 | 9 | 26 | 3 | 9 | 1 | 3 | | |
| Lymphocyte count decreased | 13 | 37 | 6 | 17 | 3 | 9 | 4 | 11 | | |
| Pruritus | 11 | 31 | 10 | 29 | 1 | 3 | | | | |
| Sore throat | 11 | 31 | 10 | 29 | 1 | 3 | | | | |
| Lipase increased | 10 | 29 | 5 | 14 | 3 | 9 | 1 | 3 | 1 | 3 |
| Aspartate aminotransferase increased | 9 | 26 | 9 | 26 | | | | | | |
| Constipation | 9 | 26 | 6 | 17 | 3 | 9 | | | | |
| Creatinine increased | 9 | 26 | 9 | 26 | | | | | | |
| Dehydration | 9 | 26 | 6 | 17 | 3 | 9 | | | | |
| Hyponatremia | 8 | 23 | 7 | 20 | 1 | 3 | | | | |
| Lymphedema | 8 | 23 | 7 | 20 | 1 | 3 | | | | |
| Rash maculo-papular | 8 | 23 | 5 | 14 | 2 | 6 | 1 | 3 | | |
| Anemia | 5 | 14 | 5 | 14 | | | | | | |
| Fever | 5 | 14 | 2 | 6 | 3 | 9 | | | | |
| Headache | 5 | 14 | 5 | 14 | | | | | | |
| White blood cell decreased | 5 | 14 | 4 | 11 | 1 | 3 | | | | |
| Aspiration pneumonia | 4 | 11 | | | | | 4 | 11 | | |
| Hyperthyroidism | 4 | 11 | 4 | 11 | | | | | | |
| Anorexia | 3 | 9 | 1 | 3 | 1 | 3 | 1 | 3 | | |
| Cough | 3 | 9 | 1 | 3 | 2 | 6 | | | | |
| Diarrhea | 3 | 9 | 3 | 9 | | | | | | |
| Mouth sore | 3 | 9 | 3 | 9 | | | | | | |
| Neck pain | 3 | 9 | 2 | 6 | 1 | 3 | | | | |
| Oral Mucositis | 3 | 9 | 1 | 3 | 2 | 6 | | | | |
| Pain of skin | 3 | 9 | 3 | 9 | | | | | | |
| TSH decreased | 3 | 9 | 3 | 9 | | | | | | |
| Vomiting | 3 | 9 | 3 | 9 | | | | | | |
| Adrenal insufficiency | 2 | 6 | 1 | 3 | | | 1 | 3 | | |
| Anxiety | 2 | 6 | 1 | 3 | 1 | 3 | | | | |

| | | | | | | | | |
|---|---|---|---|---|---|---|---|---|
| BUN increased | 2 | 6 | 2 | 6 | | | | |
| Blurred vision | 2 | 6 | 1 | 3 | 1 | 3 | | |
| Conjunctivitis | 2 | 6 | 2 | 6 | | | | |
| Dry skin | 2 | 6 | 2 | 6 | | | | |
| Insomnia | 2 | 6 | 1 | 3 | 1 | 3 | | |
| Myalgia | 2 | 6 | 2 | 6 | | | | |
| T4 decreased | 2 | 6 | 1 | 3 | 1 | 3 | | |
| Thick mucous secretions | 2 | 6 | 2 | 6 | | | | |
| Trismus | 2 | 6 | | | 2 | 6 | | |
| Abdominal discomfort | 1 | 3 | 1 | 3 | | | | |
| Acute kidney injury | 1 | 3 | 1 | 3 | | | | |
| Adjustment disorder (anxiety mixed with depression) | 1 | 3 | | | 1 | 3 | | |
| Aguesia (altered/no taste) | 1 | 3 | 1 | 3 | | | | |
| Alkaline phosphatase increased | 1 | 3 | 1 | 3 | | | | |
| Altered smell | 1 | 3 | 1 | 3 | | | | |
| Amylase | 1 | 3 | 1 | 3 | | | | |
| Amylase decreased | 1 | 3 | 1 | 3 | | | | |
| Appetite change | 1 | 3 | 1 | 3 | | | | |
| Arthralgic stiffness (Both shoulders) | 1 | 3 | 1 | 3 | | | | |
| Bleeding mouth ulcers | 1 | 3 | 1 | 3 | | | | |
| Bloating | 1 | 3 | | | 1 | 3 | | |
| Blood bilirubin increased | 1 | 3 | 1 | 3 | | | | |
| Blood/mucus in stool | 1 | 3 | 1 | 3 | | | | |
| Colitis | 1 | 3 | 1 | 3 | | | | |
| Decreased appetite | 1 | 3 | 1 | 3 | | | | |
| Decreased smell | 1 | 3 | 1 | 3 | | | | |
| Diabetic ketoacidosis | 1 | 3 | | | | | 1 | 3 |
| Dyspnea | 1 | 3 | 1 | 3 | | | | |
| Erythema multiforme | 1 | 3 | 1 | 3 | | | | |
| Erythematous papules, forehead | 1 | 3 | 1 | 3 | | | | |
| Eyelid function disorder | 1 | 3 | | | 1 | 3 | | |
| Free T3 increased | 1 | 3 | 1 | 3 | | | | |
| Free T4 decreased | 1 | 3 | 1 | 3 | | | | |
| Hypercalcemia | 1 | 3 | 1 | 3 | | | | |
| Hyperglycemia | 1 | 3 | | | | | 1 | 3 |
| Hyperkalemia | 1 | 3 | 1 | 3 | | | | |
| Hypermagnesemia | 1 | 3 | 1 | 3 | | | | |
| Hypernatremia | 1 | 3 | 1 | 3 | | | | |
| Hypoalbuminemia | 1 | 3 | 1 | 3 | | | | |
| Hypokalemia | 1 | 3 | 1 | 3 | | | | |
| Hypophosphatemia | 1 | 3 | 1 | 3 | | | | |
| Infusion related reaction | 1 | 3 | | | 1 | 3 | | |

| | | | | | | | | |
|---|---|---|---|---|---|---|---|---|
| Itchiness on back of head and lower back | 1 | 3 | | | 1 | 3 | | |
| Joint pain | 1 | 3 | | | 1 | 3 | | |
| Laryngeal inflammation | 1 | 3 | 1 | 3 | | | | |
| Light headedness | 1 | 3 | 1 | 3 | | | | |
| Loose stool | 1 | 3 | 1 | 3 | | | | |
| Mild Thrush | 1 | 3 | | | 1 | 3 | | |
| Myositis | 1 | 3 | | | | | 1 | 3 |
| Nasal congestion | 1 | 3 | | | 1 | 3 | | |
| Oral dysesthesia | 1 | 3 | | | 1 | 3 | | |
| Oral thrush | 1 | 3 | | | 1 | 3 | | |
| Oropharyngeal Candidiasis | 1 | 3 | 1 | 3 | | | | |
| Palmar-Plantar | 1 | 3 | 1 | 3 | | | | |
| Pancreatitis | 1 | 3 | | | | | 1 | 3 |
| Proteinuria | 1 | 3 | 1 | 3 | | | | |
| Rash (cheek & posterior neck) | 1 | 3 | 1 | 3 | | | | |
| Rash acneiform | 1 | 3 | | | 1 | 3 | | |
| Rash palmar-plantar | 1 | 3 | | | 1 | 3 | | |
| Red eyes with occasional itching | 1 | 3 | 1 | 3 | | | | |
| Redness, neck area | 1 | 3 | 1 | 3 | | | | |
| Stomach pain | 1 | 3 | 1 | 3 | | | | |
| Swallow pain | 1 | 3 | | | 1 | 3 | | |
| Swelling right cervical nodes | 1 | 3 | 1 | 3 | | | | |
| Swelling right nodes | 1 | 3 | 1 | 3 | | | | |
| Telangiectasia | 1 | 3 | 1 | 3 | | | | |
| Thrush | 1 | 3 | | | 1 | 3 | | |
| Thyroiditis | 1 | 3 | 1 | 3 | | | | |
| Ulcer on left lateral tongue | 1 | 3 | 1 | 3 | | | | |
| Ulceration on right side of tongue | 1 | 3 | | | 1 | 3 | | |
| Upper Airway inflammation | 1 | 3 | 1 | 3 | | | | |
| Urticaria | 1 | 3 | | | 1 | 3 | | |
| WBC increased | 1 | 3 | 1 | 3 | | | | |
| Watering eyes | 1 | 3 | 1 | 3 | | | | |

Table summarizes the treatment related adverse events (possibly, probably, or definitely attributed to immunotherapy or radiation) experienced by number of patients after first dose. Each patient with any adverse event is counted only once in this table at the highest (maximum) grade experienced.

**Supplementary Table 4. Adverse events related to immune checkpoint blockade.**

| AE | Grade 3 | | Grade 4 | |
|---|---|---|---|---|
| | Count | % | Count | % |
| Dysphagia/Odynophagia | 6 | 17 | | |
| Mucositis (tongue, throat, mouth) | 5 | 14 | | |
| Aspiration pneumonia | 4 | 11 | | |
| Lymphocyte count decreased | 4 | 11 | | |
| Serum amylase increased | 4 | 11 | | |
| Weight loss | 4 | 11 | | |
| Fatigue | 3 | 9 | | |
| Pain (tongue, mouth, throat) | 3 | 9 | | |
| Adrenal insufficiency | 1 | 3 | | |
| Alanine aminotransferase increased | 1 | 3 | | |
| Anorexia | 1 | 3 | | |
| Diabetic ketoacidosis | 1 | 3 | | |
| Hyperglycemia | 1 | 3 | | |
| Lipase increased | 1 | 3 | 1 | 3 |
| Myositis | 1 | 3 | | |
| Pancreatitis | 1 | 3 | | |
| Rash maculo-papular | 1 | 3 | | |

**Supplementary Table 5. Serious adverse events.**

| Accession | Adverse events | Attribution | Grade |
|---|---|---|---|
| 1 | Dehydration | 1 | 3 |
| 9 | Hyponatremia | 1 | 3 |
| 13 | Myositis | 5 | 3 |
| 23 | Aspiration | 5 | 3 |
| 24 | Aspiration pneumonia | 5 | 3 |
| 26 | Dysphagia | 5 | 3 |
| 26 | Neck pain | 5 | 2 |
| 26 | Anorexia | 5 | 3 |
| 26 | Suicidal ideation | 2 | 3 |
| 29 | Diabetic ketoacidosis | 3 | 3 |
| 36 | Dysphagia | 5 | 3 |
| 38 | Aspiration pneumonia | 2 | 3 |
| 38 | Encephalopathy | 2 | 2 |
| 38 | Adrenal insufficiency | 3 | 3 |
